# COVID-19 Prognostic Modeling Using CT Radiomic Features and Machine Learning Algorithms: Analysis of a Multi-Institutional Dataset of 14,339 Patients

**DOI:** 10.1101/2021.12.07.21267364

**Authors:** Isaac Shiri, Yazdan Salimi, Masoumeh Pakbin, Ghasem Hajianfar, Atlas Haddadi Avval, Amirhossein Sanaat, Shayan Mostafaei, Azadeh Akhavanallaf, Abdollah Saberi, Zahra Mansouri, Dariush Askari, Mohammadreza Ghasemian, Ehsan Sharifipour, Saleh Sandoughdaran, Ahmad Sohrabi, Elham Sadati, Somayeh Livani, Pooya Iranpour, Shahriar Kolahi, Maziar Khateri, Salar Bijari, Mohammad Reza Atashzar, Sajad P. Shayesteh, Bardia Khosravi, Mohammad Reza Babaei, Elnaz Jenabi, Mohammad Hasanian, Alireza Shahhamzeh, Seyed Yaser Foroghi Gholami, Abolfazl Mozafari, Arash Teimouri, Fatemeh Movaseghi, Azin Ahmari, Neda Goharpey, Rama Bozorgmehr, Hesamaddin Shirzad-Aski, Rozbeh Mortazavi, Jalal Karimi, Nazanin Mortazavi, Sima Besharat, Mandana Afsharpad, Hamid Abdollahi, Parham Geramifar, Amir Reza Radmard, Hossein Arabi, Kiara Rezaei-Kalantari, Mehrdad Oveisi, Arman Rahmim, Habib Zaidi

**Author notes:** Corresponding author: Habib Zaidi, Ph.D Geneva University Hospital, Division of Nuclear Medicine and Molecular Imaging CH-1211 Geneva, Switzerland. **First Author: Isaac Shiri**, MSc Geneva University Hospital, Division of Nuclear Medicine and Molecular Imaging, CH-1211 Geneva, Switzerland.

## Abstract

**Objective:** In this large multi-institutional study, we aimed to analyze the prognostic power of computed tomography (CT)-based radiomics models in COVID-19 patients.

**Methods:** CT images of 14,339 COVID-19 patients with overall survival outcome were collected from 19 medical centers. Whole lung segmentations were performed automatically using a previously validated deep learning-based model, and regions of interest were further evaluated and modified by a human observer. All images were resampled to an isotropic voxel size, intensities were discretized into 64-binning size, and 105 radiomics features, including shape, intensity, and texture features were extracted from the lung mask. Radiomics features were normalized using Z-score normalization. High-correlated features using Pearson (R^2^>0.99) were eliminated. We applied the Synthetic Minority Oversampling Technique (SMOT) algorithm in only the training set for different models to overcome unbalance classes. We used 4 feature selection algorithms, namely Analysis of Variance (ANOVA), Kruskal-Wallis (KW), Recursive Feature Elimination (RFE), and Relief. For the classification task, we used seven classifiers, including Logistic Regression (LR), Least Absolute Shrinkage and Selection Operator (LASSO), Linear Discriminant Analysis (LDA), Random Forest (RF), AdaBoost (AB), Naïve Bayes (NB), and Multilayer Perceptron (MLP). The models were built and evaluated using training and testing sets, respectively. Specifically, we evaluated the models using 10 different splitting and cross-validation strategies, including different types of test datasets (e.g. non-harmonized vs. ComBat-harmonized datasets). The sensitivity, specificity, and area under the receiver operating characteristic (ROC) curve (AUC) were reported for models evaluation.

**Results:** In the test dataset (4301) consisting of CT and/or RT-PCR positive cases, AUC, sensitivity, and specificity of 0.83±0.01 (CI95%: 0.81-0.85), 0.81, and 0.72, respectively, were obtained by ANOVA feature selector + RF classifier. In RT-PCR-only positive test sets (3644), similar results were achieved, and there was no statistically significant difference. In ComBat harmonized dataset, Relief feature selector + RF classifier resulted in highest performance of AUC, reaching 0.83±0.01 (CI95%: 0.81-0.85), with sensitivity and specificity of 0.77 and 0.74, respectively. At the same time, ComBat harmonization did not depict statistically significant improvement relevant to non-harmonized dataset. In leave-one-center-out, the combination of ANOVA feature selector and LR classifier resulted in the highest performance of AUC (0.80±0.084) with sensitivity and specificity of 0.77 ± 0.11 and 0.76 ± 0.075, respectively.

**Conclusion:** Lung CT radiomics features can be used towards robust prognostic modeling of COVID-19 in large heterogeneous datasets gathered from multiple centers. As such, CT radiomics-based model has significant potential for use in prospective clinical settings towards improved management of COVID-19 patients.

## INTRODUCTION

The novel coronavirus disease which emerged in 2019 (COVID-19) is now a major cause of death worldwide ^1^. This highly contagious virus can cause a spectrum of pulmonary, hematological, neurological, and systemic complications, making it a highly lethal pathogen ^2^. As of August 23^st^, 2021, there have been >200 million globally confirmed cases of COVID-19, including >4 million deaths and >4 billion vaccinations reported to the world health organization (WHO) [https://covid19.who.int/]. There remains an urgent need for addressing issues such as diagnosis, prognosis, and treatment options ^3^.

Diagnostic tools for COVID-19, such as reverse transcription polymerase chain reaction (RT-PCR) aid to distinguish between negative and positive cases ^4^. Prognostic tools, on the other hand, provide clinicians with insights to optimize treatment strategies, manage the hospitalization of patients both in the wards and intensive care units (ICU), and better handle patient follow-up plans ^5^. Different studies have evaluated clinical and/or non-clinical features for determining the diagnosis and prognosis of patients with COVID-19. Yan *et al.* ^6^ used only clinical features to classify patients into different categories, ranging from mild to critical conditions. Zhou *et al.* ^7^ also aimed at establishing a prognostic model for outcome prediction of patients with COVID-19, utilizing their clinical data.

Computed Tomography (CT) plays a pivotal role in the management of a wide variety of diseases as a fast and non-invasive imaging modality. In the case of COVID-19, CT is used for both diagnostic (e.g. in case of limited access to RT-PCR) and prognostic purposes ^8^. The clinical value of CT imaging relies mainly on the early detection of lung infections and high accuracy in quantifying the disease progression and severity ^9^. Francone *et al.* ^10^ assessed the correlation of CT scores with COVID-19 pneumonia severity and outcome. In another study, Zhao *et al.* ^11^ specified pulmonary involvement severity by measuring the extent of pneumonia and consolidation that appear on CT images. Li *et al.* ^12^ concluded that high scores of CT images are associated with severe COVID-19 pneumonia.

In spite of previously conducted research, there is still a need for studies with more accurate and comprehensive analyses ^13^. In conventional analyses, CT features are visually and subjectively defined, while machine learning (ML) and/or deep learning (DL) models have the potential to provide more comprehensive and objective assessment of images. Towards modeling of outcomes in COVID-19 patients, several ML and DL algorithms have been utilized to assess severity and to predict outcome of patients using CT imaging ^14–17^. To evaluate the sensitivity and specificity of CT for COVID-19 diagnostic purposes, Harmon *et al.* ^14^ achieved a sensitivity of 84% and specificity of 93% in an independent test set of 1337 patients applying DL on CT images. In another machine learning study, Mei *et al.* ^15^ reported a sensitivity of 84.3% and specificity of 82.8% based on combined CT and clinical data. Cai *et al.* ^16^ utilized a random forest model to assess the severity of COVID-19 disease in 99 patients and their need for a longer hospital or ICU stay. Another study by Lessman *et al.* ^17^ reported the model performance of three DL models for severity assessment in COVID-19. In another multi-center study, Meng *et al.* ^18^ differentiated patients with high-risk of mortality versus low-risk ones using a convolutional neural network named De-COVID-Net. Ning *et al.* ^19^ used their pre- trained DL model on a dataset consisting of 351 patients that was capable of distinguishing between non-coronavirus pneumonia, mild coronavirus pneumonia, and severe forms of COVID-19 disease.

Various studies also reported remarkable prediction accuracies utilizing radiomics approaches using CT and chest x-ray imaging modalities ^20–23^. Medical images could be converted into high-dimensional data by means of radiomics, wherein radiomics features are selected from the images and combined using machine learning algorithms to arrive at radiomics signatures as biomarkers of disease^24–34^. In addition to wide usage in several oncologic ^35–37^ as well as non-oncologic diseases ^27, 38^, radiomics studies have indicated that imaging features extracted from CT or chest X-ray images could be used as parameters for outcome prediction of patients with COVID-19 pneumonia. Radiomics analyses have been applied to different aspects of COVID-19, including diagnosis, severity scoring, prognosis, hospital/ICU stay prediction, and survival analysis ^20–23^. In a retrospective study, Fu *et al.* ^39^ constructed a predictive model based on CT radiomics, clinical and laboratory features. This signature could classify COVID-19 patients into stable and unstable (i.e. progressive phenotype). Homayounieh *et al.* ^40^ aimed to predict the severity of pneumonia in patients with COVID-19 using a radiomics model that outperformed models consisting of clinical-only features. Another study by Li *et al.* ^41^ analyzed a radiomics/DL model that distinguished severe from critical COVID-19 pneumonia patients. Cai *et al.* ^42^ developed a model by means of combining CT radiomics features and clinical data to predict RT-PCR negativity during admission. Yue *et al.* ^43^ conducted a multicentric radiomics study on 52 patients to differentiate whether an individual needs a short-term or long-term hospital stay. Another study by Bae *et al.* ^44^ predicted the mortality of patients with COVID-19 using chest x-ray radiomics. Their model could identify whether a patient needs mechanical ventilation or not. Artificial intelligence (AI) has been widely used in radiology to provide diagnostic and prognostic tools to help clinicians during the pandemic. However, owing to the lack of standardization in AI studies in terms of data collection, methodology, and evaluation, most of these studies were not pragmatic when it comes to clinical adoption ^13, 45, 46^. In a recent study by Roberts *et al.* ^13^, possible sources of bias in more than 2000 AI articles in COVID-19 were evaluated in both deep learning and traditional machine learning-based studies. This review showed that bias do exist in most, if not all, of the studies in different domains, including dataset and methodology. In the dataset domain, several articles used public datasets which can contain duplicates, low-quality images, false demographics, or unknown clinical/lab data of patients. These public datasets can also induce bias in the outcome domain as they may fail to supply sufficient information about how they exactly proved a patient is COVID-19 positive or how imaging data were acquired in terms of image acquisition and reconstruction. They also mentioned using small datasets, Frankenstein datasets ^13^, and Toy datasets ^45^ in several articles. In the methodology domain, most of the studies did not provide all methodological information or did not perform a standard AI analysis based on guidelines ^47, 48^.

Overall, Roberts *et al.* ^13^ reviewed 69 traditional machine learning/radiomics studies and reported that 44 were excluded because of Radiomics Quality Score (RQS) ^47^ of less than six or not describing the datasets appropriately. From the remaining 25, six articles performed model evaluation using external validation sets and only four papers reported the significance of their model along with the statistical parameters (agreement level). They also assessed bias in the prognostication studies in the four areas of prediction model risk of bias assessment tool (PROBAST) guide and reported high bias in participants, predictors, outcomes, and analysis areas. Overall, there are several radiomics studies targeting improved COVID-19 diagnosis or prognosis. However, owing to the limited sample size, single-centered nature of most of the databases, and variability in data acquisition and image reconstruction parameters, the models tend to overfit ^13^. Providing a generalizable model which is reproducible on unseen datasets of other centers is highly desired. In this context, we designed a large multi-institutional study to build and evaluate a radiomics model based on a large-scale CT imaging dataset aimed at the prediction of survival (alive or deceased) in COVID-19 infected patients. We built and evaluated our model based on different guidelines and tested different machine learning algorithms in different strategies to evaluate model reproducibility and repeatability in a large dataset.

## MATERIALS AND METHODS

Figure 1 summarizes the different steps adopted in this study. To provide a standard and reproducible study, we completed different checklists/guidelines concerning predictive modeling, radiomics studies, and artificial intelligence studies. The Transparent Reporting of a multivariable prediction model for Individual Prognosis or Diagnosis (TRIPOD) [40] checklist is provided in supplemental Table 1. We also reported the Radiomics Quality Score (RQS) based on Lambin et al. ^47^ and the Checklist for Artificial Intelligence in Medical imaging (CLAIM) ^48^ in supplemental material. These checklists were filled out by two individuals (with consensus) who are experts in radiomics field and not co-authors in this study.

**Figure 1:**
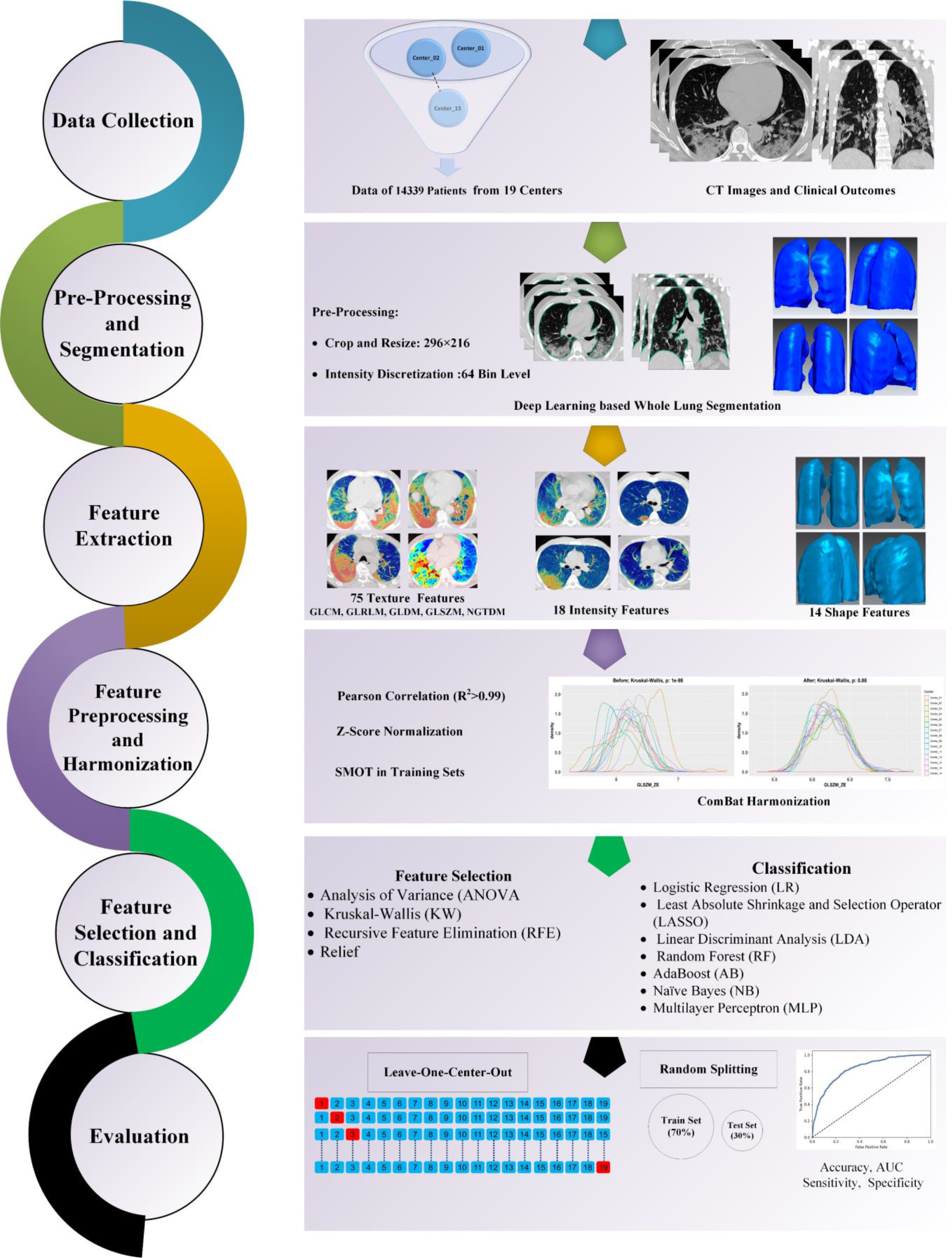
The flow chart of our study represents the different radiomics steps.

**Table 1.** Demographics and data acquisition parameters across different centers.

| Center | Number (Deceased %) | PCR | Gender |  | CTDI<sub>vol</sub> | Tube Current | Age | Slice Thickness |
| --- | --- | --- | --- | --- | --- | --- | --- | --- |
|  |  | % of patients with available PCR result | Male | Female | Mean±SD | Mean±SD | Mean±SD | Mean±SD |
| Center_01 | 152 (26.3%) | 100% | 87 | 65 | 1.57±0.32 | 45± 47.8 | 62.1 ± 17.0 | 1.50 ± 0.50 |
| Center_02 | 264 (41.6%) | 99% | 120 | 144 | 8.47±0.53 | 186.2±34.0 | 71.8 ± 12.4 | 3.44 ± 0.95 |
| Center_03 | 319 (1.5%) | 100% | 168 | 151 | 8.55 ± 4.61 | 185 ± 65.3 | 51.6 ± 17.9 | 2.82 ± 0.99 |
| Center_04 | 373 (22.5%) | 77% | 157 | 216 | 7.52±1.23 | 147.1±43.2 | 54.1 ± 19.3 | 5.22 ± 0.46 |
| Center_05 | 382 (19.1%) | 100% | 215 | 167 | 6.37±0.67 | 128.3±4.5 | 58.2±18.25 | 7.67 ± 1.7 |
| Center_06 | 394 (38.6%) | 100% | 257 | 137 | 5.8 ± 2.52 | 149 ± 48.3 | 56.8 ± 16.8 | 2.34 ± 0.24 |
| Center_07 | 492 (5.2%) | 100% | 276 | 216 | 9.68 ± 2.91 | 349 ± 181 | 33.0 ± 5.66 | 2.57 ± 1.36 |
| Center_08 | 539 (19.1%) | 94% | 287 | 252 | 4.10 ± 4.57 | 167 ± 39.1 | 54.1 ± 17.4 | 7.71 ± 0.77 |
| Center_09 | 608 (3.0 %) | 100% | 274 | 334 | 6.61 ± 4.87 | 164 ± 128 | 55.2 ± 15.99 | 2.10 ± 1.21 |
| Center_10 | 685 (14.8%) | 89% | 348 | 337 | 5.01 ± 3.57 | 171 ± 33.3 | 56.4 ± 18.7 | 7.16 ± 0.81 |
| Center_11 | 866 (13.2%) | 100% | 520 | 346 | 3.40±0.42 | 123.6±9.6 | 55.4 ± 17.9 | 2.04 ± 0.23 |
| Center_12 | 883 (11.2%) | 62% | 448 | 435 | 4.30±1.54 | 166.2±51.3 | 55.4 ± 15.6 | 4.99 ± 0.16 |
| Center_13 | 949 (10.9%) | 76% | 568 | 381 | 5.57±1.21 | 210.7±39.6 | 52.6 ± 19.3 | 2.17 ± 1.51 |
| Center_14 | 1022 (9.7%) | 100% | 526 | 496 | 5.01 ± 3.19 | 91.6 ± 48.6 | 56.2 ± 18.8 | 4.93 ± 0.49 |
| Center_15 | 1024 (1.0%) | 29% | 542 | 482 | 6.82±1.02 | 228.4±27.2 | 48.3 ± 14.5 | 2.48 ± 1.96 |
| Center_16 | 1180 (12.7%) | 84% | 578 | 602 | 5.63± 2.98 | 172 ± 34.1 | 55.4 ± 16.7 | 6.38 ± 0.82 |
| Center_17 | 1323 (12.7%) | 100% | 680 | 643 | 6.14 ± 1.41 | 175 ± 38.2 | 57.4 ± 18.7 | 5.66 ± 0.82 |
| Center_18 | 1355 (2.5%) | 70% | 752 | 603 | 4.21 ± 3.18 | 98.4 ± 41.1 | 55.1 ± 19.1 | 4.75 ± 0.69 |
| Center_19 | 1529 (5.6%) | 93% | 814 | 715 | 8.92±1.40 | 220.7±27.9 | 54.5 ± 19.0 | 6.96 ± 0.34 |

### Patient Population

This study was approved by our local institutional review board (IRB), and written informed consent of patients was waived by the ethics committee as anonymized data were used without any interventional effect on diagnosis, treatment or management of COVID-19 patients.

In the first step, 24,448 patients, from 19 medical centers in Iran, suspected of COVID-19 and with acquired chest CT images were included. Different exclusion criteria were applied to provide a reliable dataset. We excluded: (i) patients without follow-up information or clear evidence of clinical endpoint, or if they were transferred to another medical center (3519 patients), or patients with (ii) negative RT-PCR (1860 patients), (iii) laboratory-confirmed pneumonia of other types (1606 patients), (iv) confirmed lung cancer or metastases from other origins to the lungs (1400 patients), (v) atypical CT findings for other abnormalities (850 patients), (vi) CT images with contrast media administration (58 patients), (vii) severe motion or bulk motion artifacts in CT images were carefully checked (515 patients), (viii) extremely inappropriate positioning which resulted in missing the upper and lower bounds of the lungs (121 patients), or (ix) CT images with extremely low quality or SNR (210 patients).

Considering these criteria, we excluded 10,149 patients from further analysis (Figure 2). Hence, 14,339 chest CT scans (with one scan per patient whose COVID-19 was confirmed either by RT-PCR or CT imaging) were included in this study. Common symptoms of COVID-19, including fever, respiratory symptoms, shortness of breath, dry cough and tiredness were recorded and contact history with COVID-19 patients was also assessed. In each center, CT images were evaluated at least by two radiologists and in case of discrepancy, a third radiologist was involved to settle the disagreement. As defined in the COVID-19 Reporting and Data System (CO-RADS) ^49^, typical manifestations of COVID-19, such as ground-glass opacity, consolidation, crazy-paving pattern, or dominant peripheral distribution of parenchymal abnormalities were considered diagnostic for COVID-19 in CT images.

**Figure 2:**
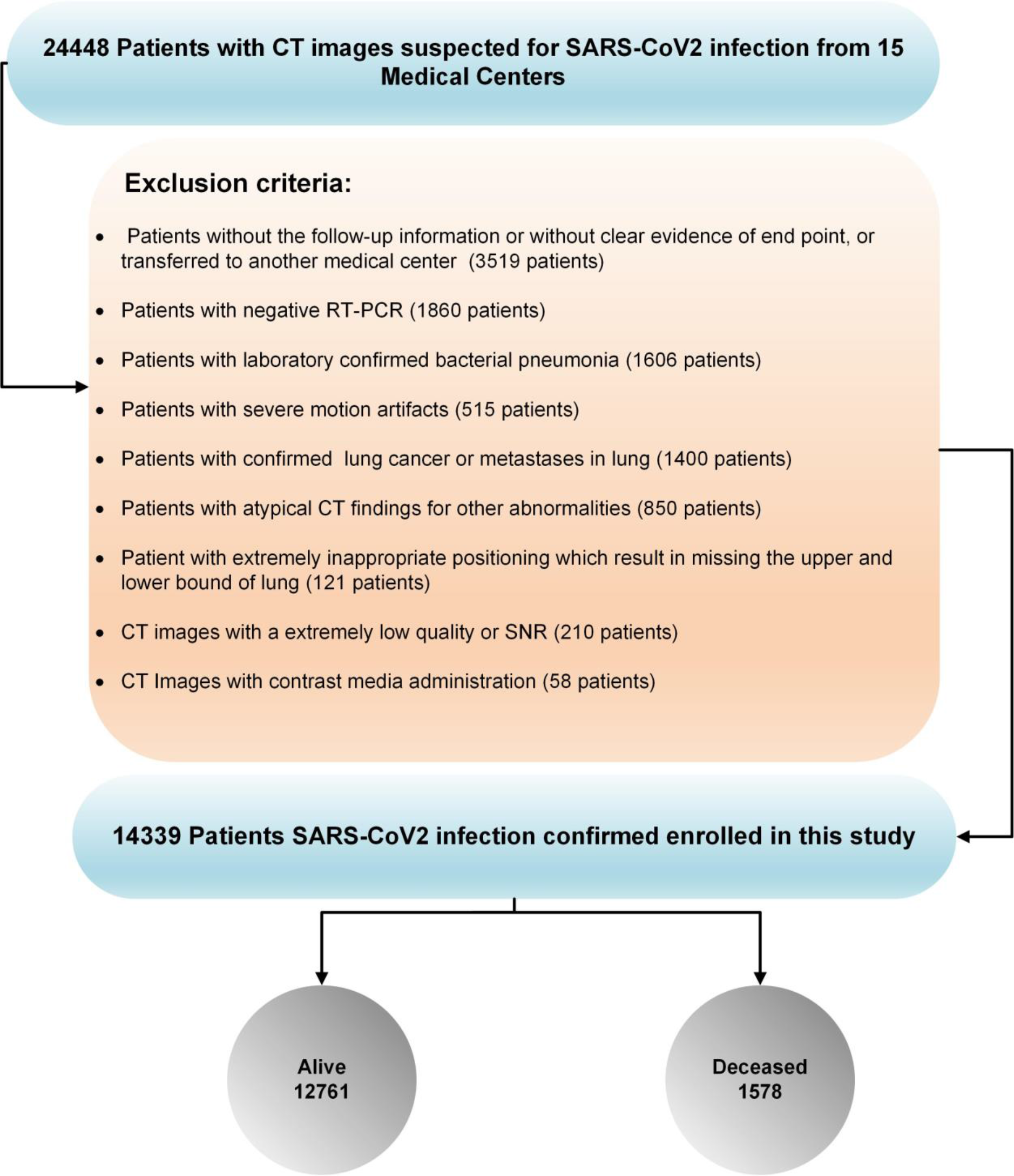
Inclusion and exclusion criteria in this study. Fraction of deceased patients is overrepresented in our data due to our exclusion criteria.

Among these studies, 13,741 CT images were collected from 18 centers in Iran (1560 deceased, 12,171 alive; fraction of deceased cases are significantly overrepresented due to our exclusion criteria), and 608 images were gathered from an online open-access databases from China (Center 9: 18 deceased, 590 alive) ^50^. All patients from Iran received standard treatment regimens according to the interim national COVID-19 treatment guideline [corona.behdasht.gov.ir]. Only one center (Center 10) included outpatient studies and the rest were inpatient-only studies from hospitalized patients. Follow-up was performed 3-4 months after the initial CT scan in the outpatient cases. For admitted patients (inpatient), follow-up was performed until discharge from the hospital, which was considered after careful evaluation of patients by the attending physician based on several criteria, including stable hemodynamics state (BP>90/60, HR<120), absence of fever for > 2 days, absence of any respiratory distress, blood oxygen saturation >93% in ambient oxygen without the need for supplementary oxygen, and no need for hospitalization for any other pathology.

### CT Image Acquisition

All chest CT images from the Iranian centers were acquired according to an institutional variation of the national society of radiology COVID-19 imaging guidelines ^51^. Image acquisition was performed during breath-hold to reduce motion artifacts. Variations in CT imaging protocols among centers were observed which led to considerable variability in image quality and radiation dose. Volumetric CT Dose Index (CTDIvol), as a parameter representing vendor-free information on radiation exposure, was reported to better reflect intra/inter institutional variability of our dataset. Table 1 summarizes the image acquisition characteristics of each center, including the number of images, acquisition parameters (slice thickness, tube current), and CTDIvol.

### Image Segmentation and Image Preprocessing

The lungs were automatically segmented using our DL-based algorithm named COLI-NET which we previously proposed and evaluated ^52^. For efficient radiomics feature extraction (feature extraction time), all images were first cropped to the lung region and then resized to 296×216 to obtain a computationally efficient feature extraction. After reviewing the segmentations, the image voxel was resized to an isotropic voxel size of 1×1×1 mm^3^, and the intensity discretized to 64- binning size ^53^.

### Radiomics Feature Extraction and Harmonization

After image preprocessing, radiomics feature extraction was performed using the PyRadiomics Python library ^54^. Radiomics features, including morphological (n=16), intensity (n=17), and texture features including second-order features, such as Gray Level Co-occurrence Matrix (GLCM, n=24), higher-order features namely Gray Level Size Zone Matrix (GLSZM, n=16), Neighboring Gray Tone Difference Matrix (NGTDM, n=5), Gray Level Run Length Matrix (GLRLM, n=16), and Gray Level Dependence Matrix (GLDM, n=14) were extracted in compliance with the Image Biomarker Standardization Initiative (IBSI) guidelines ^53^.

### Feature Preprocessing

For each feature vector, the mean and standard deviation were calculated (in training sets) and then normalized using Z-Score normalization, which consists of subtracting each feature vector from the mean followed by division by the standard deviation. For Z-score normalization, the mean and standard deviation were calculated for the training set and then applied on test set. Features’ correlation was evaluated using Pearson correlation and features with high correlation (R^2^>0.99) were eliminated. Owing to unbalanced datasets in training and test set, we applied Synthetic Minority Oversampling Technique (SMOT) algorithm to only the training set for the different models.

### Feature Selection and Classification

In this study, we used 4 feature selection algorithms, including Analysis of Variance (ANOVA), Kruskal-Wallis (KW), Recursive Feature Elimination (RFE), and Relief. Feature preprocessing and selection were performed on training sets and then applied on test sets. All test and external validation sets were unseen to feature processing and the selection and model building process. For classification task, we used seven classifiers, including Logistic Regression (LR), Least Absolute Shrinkage and Selection Operator (LASSO), Linear Discriminant Analysis (LDA), Random Forest (RF), AdaBoost (AB), Naïve Bayes (NB) and Multilayer Perceptron (MLP). By cross-combination of four feature selectors and seven classifiers, we tested twenty-eight different combinations.

### Evaluation

For thorough assessment, we trained and evaluated our models using 10 different strategies as summarized in Figure 3. To evaluate the models on whole datasets without considering data variability in each center, we divided the dataset of each center to 70% training and 30% tests sets resulting in the following two strategies (1 and 2):

**Figure 3:**
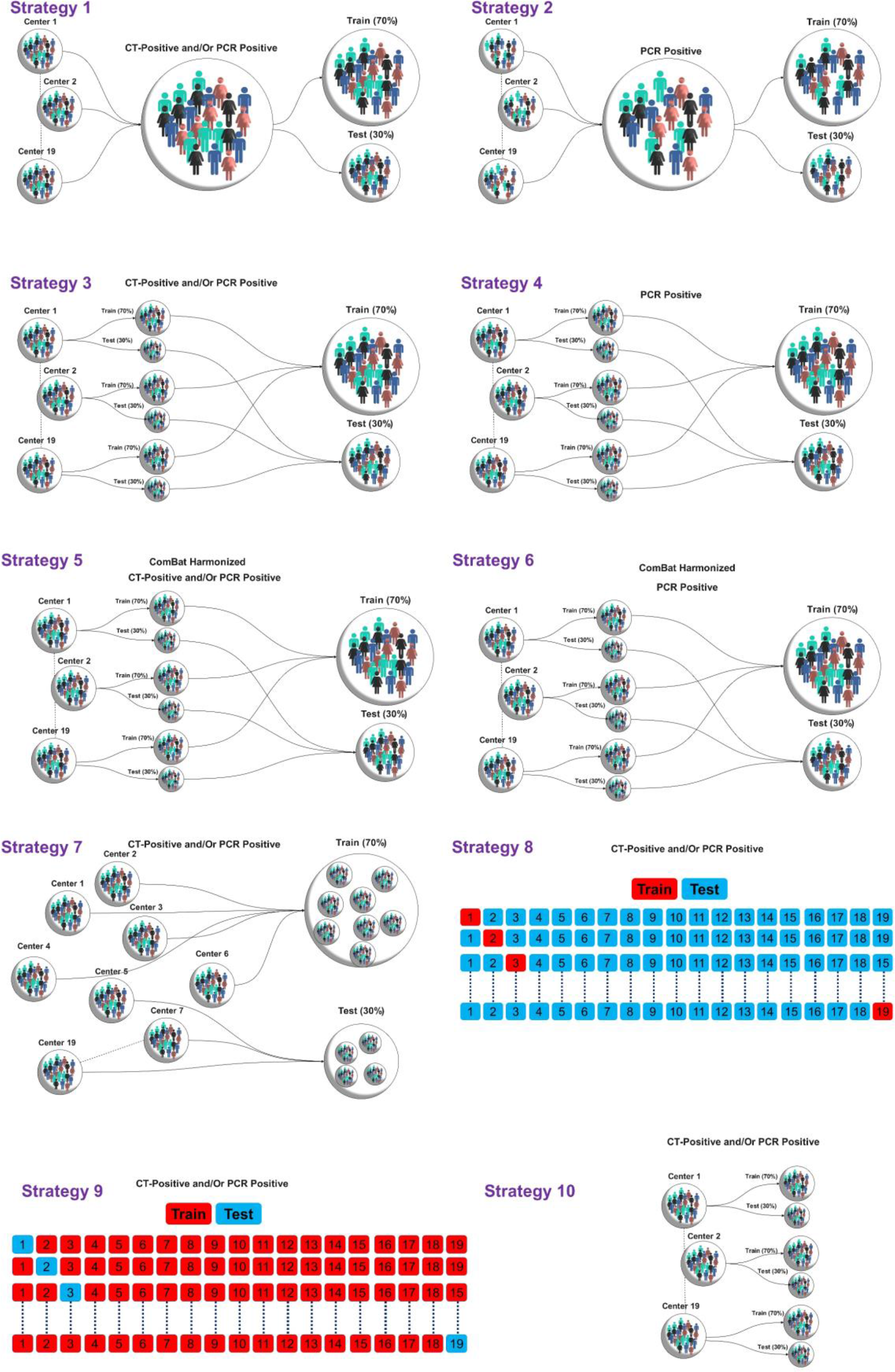
Different strategies implemented in this study for model evaluation

- Random Splitting method #1: Non-harmonized datasets were randomly split into 70% (10,038 patients) and 30% (4301 patients) for training and test sets, respectively, without considering centers. The data included patients whose COVID-19 was confirmed using RT-PCR and patients confirmed only by imaging. This test dataset included both populations.

- Random Splitting method #2: Non-harmonized datasets were randomly split into 70% (8503 patients) and 30% (3644 patients) for the training and test sets, respectively, without considering centers. The train and test sets consisted of only patients with positive RT-PCR.

- To evaluate the models on whole datasets considering data variability in each center, we divided the dataset of each center to 70% training and 30% test sets resulting in the following two strategies (3 and 4):

- Random Splitting method #3: Data from each center (non-harmonized) were randomly split into 70% (10,048 patients) and 30% (4291 patients) for the training and test sets, respectively. As our data included patients whose COVID-19 was confirmed using RT-PCR and patients confirmed only by imaging, this test dataset included both populations.

- Random Splitting method #4: Data from each center were randomly split into 70% (10,704 patients) and 30% (3635 patients) for the training and test sets, respectively. The train and test sets consist of only patients with positive RT-PCR.

To evaluate the models in the whole dataset by removing data variability due to acquisition/reconstruction from different centers, ComBat harmonization proposed by Johnson *et al.* ^55^ was applied to the extracted features to tackle the effect of center-based imaging variability. The impact of ComBat harmonization on radiomics features was assessed by Kruskal-Wallis test. After applying ComBat harmonization, we divided the datasets of each center to 70/30% train/test sets resulting in the following two strategies (3 and 4):

- Random Splitting method #5: Data from each center (ComBat harmonization) were randomly split into 70% (10,048 patients) and 30% (4291 patients) for the training and test sets, respectively. As our data included patients whose COVID-19 was confirmed using RT-PCR and patients confirmed only by imaging, this test dataset included both populations.

- Random Splitting method #6: Data from each center (ComBat harmonization) were randomly split into 70% (10,704 patients) and 30% (3635 patients) for the training and test sets, respectively. The train and test sets consisted of only patients with positive RT-PCR.

To evaluate model generalizability and sensitivity to datasets, we performed model assessment using the following strategies (7 to 9) on the external validation sets:

- Random Splitting method #7: Data (non-harmonized) were randomly split into 70% (10,655 patients) and 30% (3684 patients) for the training and external validation sets, respectively. The center’s number in the test set appears in the test sets.

- Center-based model evaluation #8: we built models on one center’s dataset (non-harmonized) and then evaluated on 18 remaining centers (external validation set), and then repeated this process for all datasets.

- Leave-one-center-out (LOCO) #9: On each of the 19 iterations, 18 centers were used as the training set, and one as the external validation set (unseen data during training). We repeated this process for all center datasets (non-harmonized).

To evaluate of model sensitivity to each dataset, we trained and tested the models in each center separately on each center dataset using the following strategies:

- Random Splitting method #10: Data from each center (non-harmonized) were randomly split into 70% and 30% for the training and test sets, respectively. The models were built and evaluated on each center separately.

All multivariate steps, including feature preprocessing, feature selection and classification were performed separately for each strategy. Classification algorithms were optimized during training using grid search algorithms. The best models were chosen by one standard deviation rule in 10- fold cross-validation and then evaluated on test or external validation sets. The accuracy, sensitivity, specificity, and area under the receiver operating characteristic curve (AUC) were reported for the test or external validation sets (unseen during training). Statistical comparison of AUCs (by 10000 bootstrapping) between models was performed using the DeLong test ^56^. The significance level was considered at a level of 0.05. All multivariate analysis steps were performed using Python Scikit-Learn open-source library.

## RESULTS

Figures 4 depicts the hierarchical clustering heat map of radiomics features distribution in alive and deceased groups for the whole dataset prior to ComBat harmonization. Supplemental Figure 1 shows the cluster heat map of radiomics features in the non-harmonized data set. Figure 5 shows the correlation of radiomics features in the whole dataset, whereas supplemental Figure 2 represents the same for ComBat harmonized features. The statistical differences calculated using the Kruskal-Wallis test are presented in supplemental Table 1 before and after ComBat harmonization. The results of ComBat harmonization showed that the algorithm properly eliminated the center effect on radiomics features in most features. ComBat harmonization data only were used for strategies 5 and 6. Figures 6-8 provide the classifications power indices of AUC, sensitivity and specificity, respectively, for splitting strategies 1-10. More detailed results were presented in supplemental Tables 2-11 for the different strategies.

**Figure 4:**
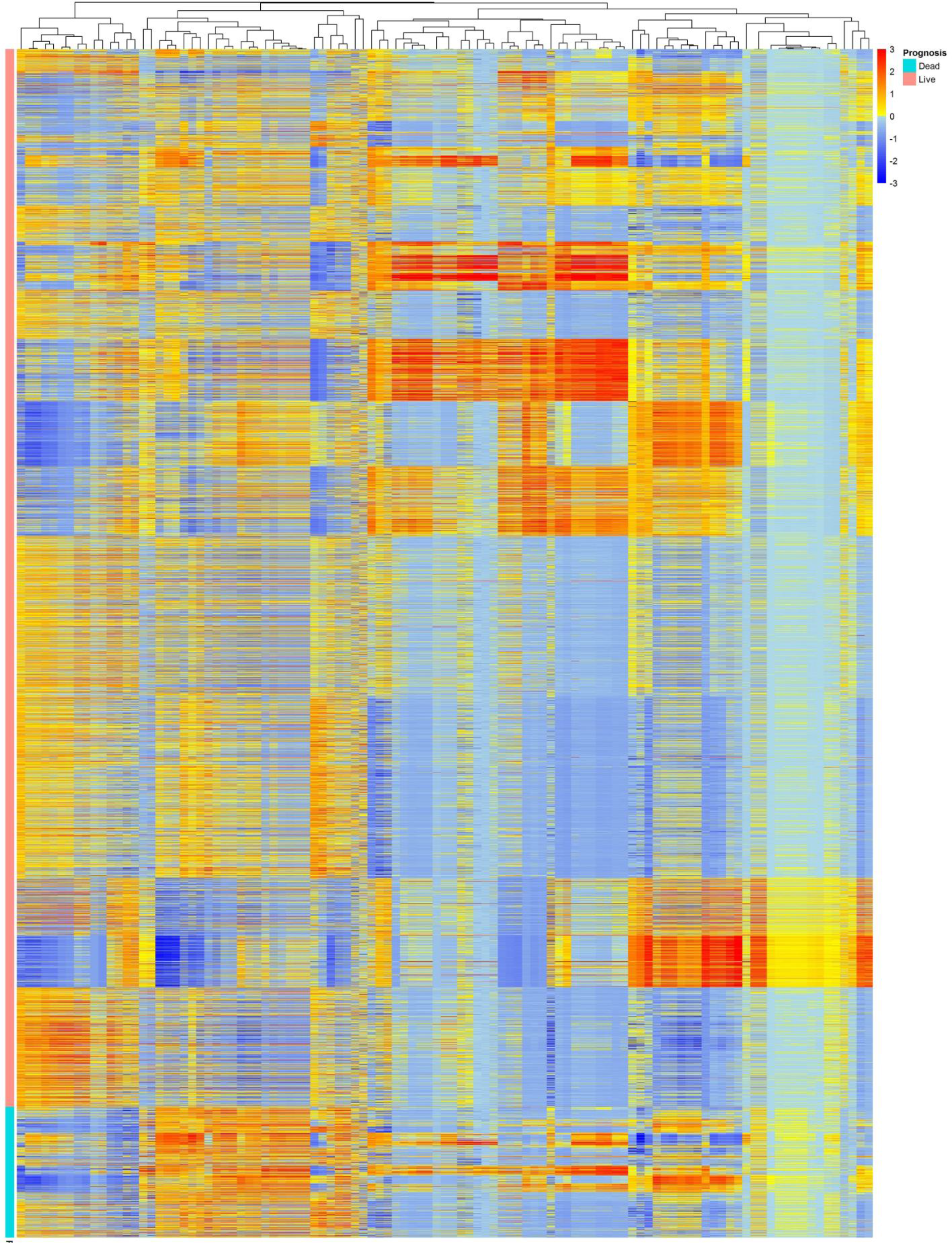
Cluster heat map of radiomics features in non-harmonized data set

**Figure 5.**
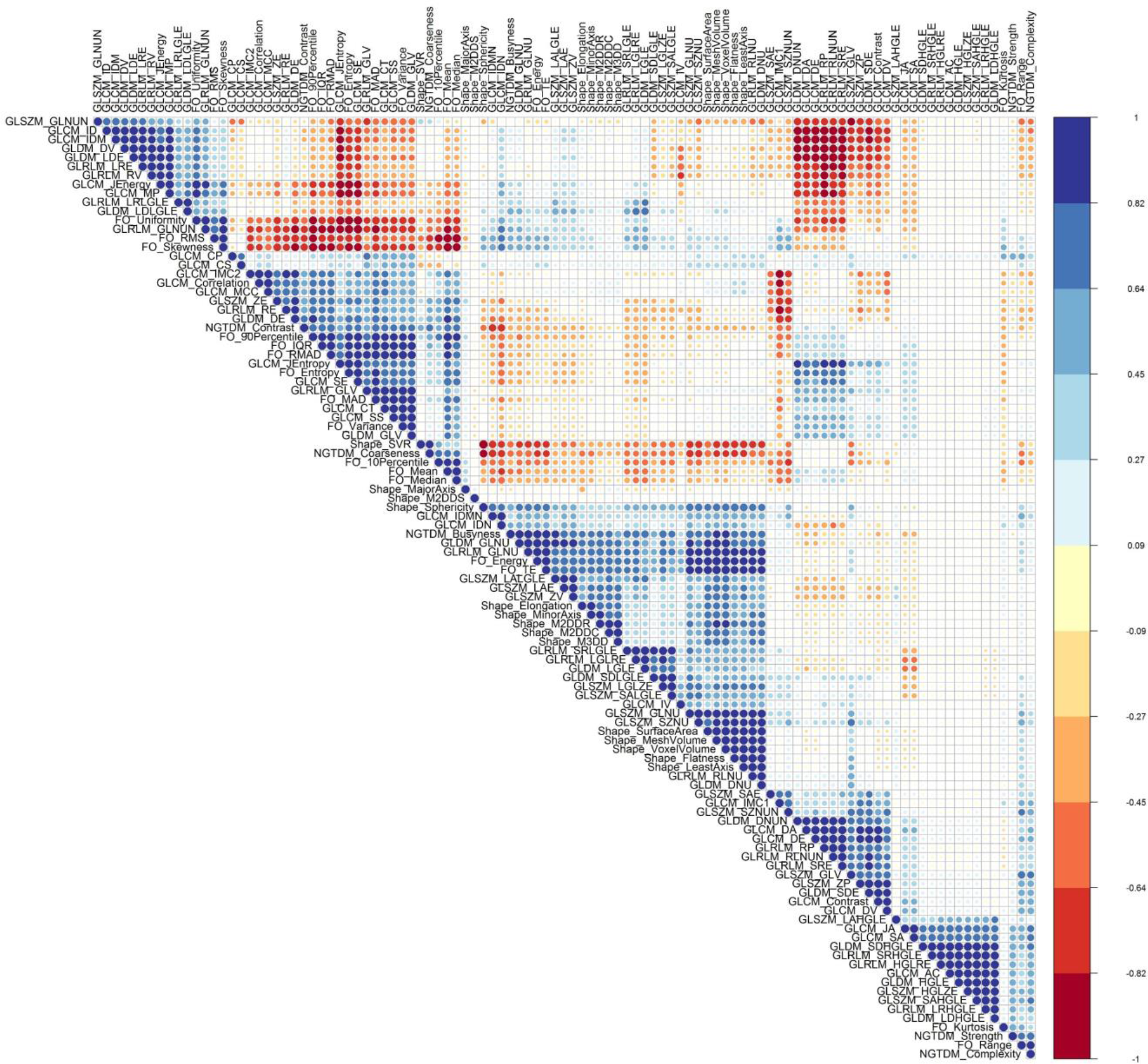
Radiomics feature correlation using Pearson correlation in non-harmonized data set

**Figure 6:**
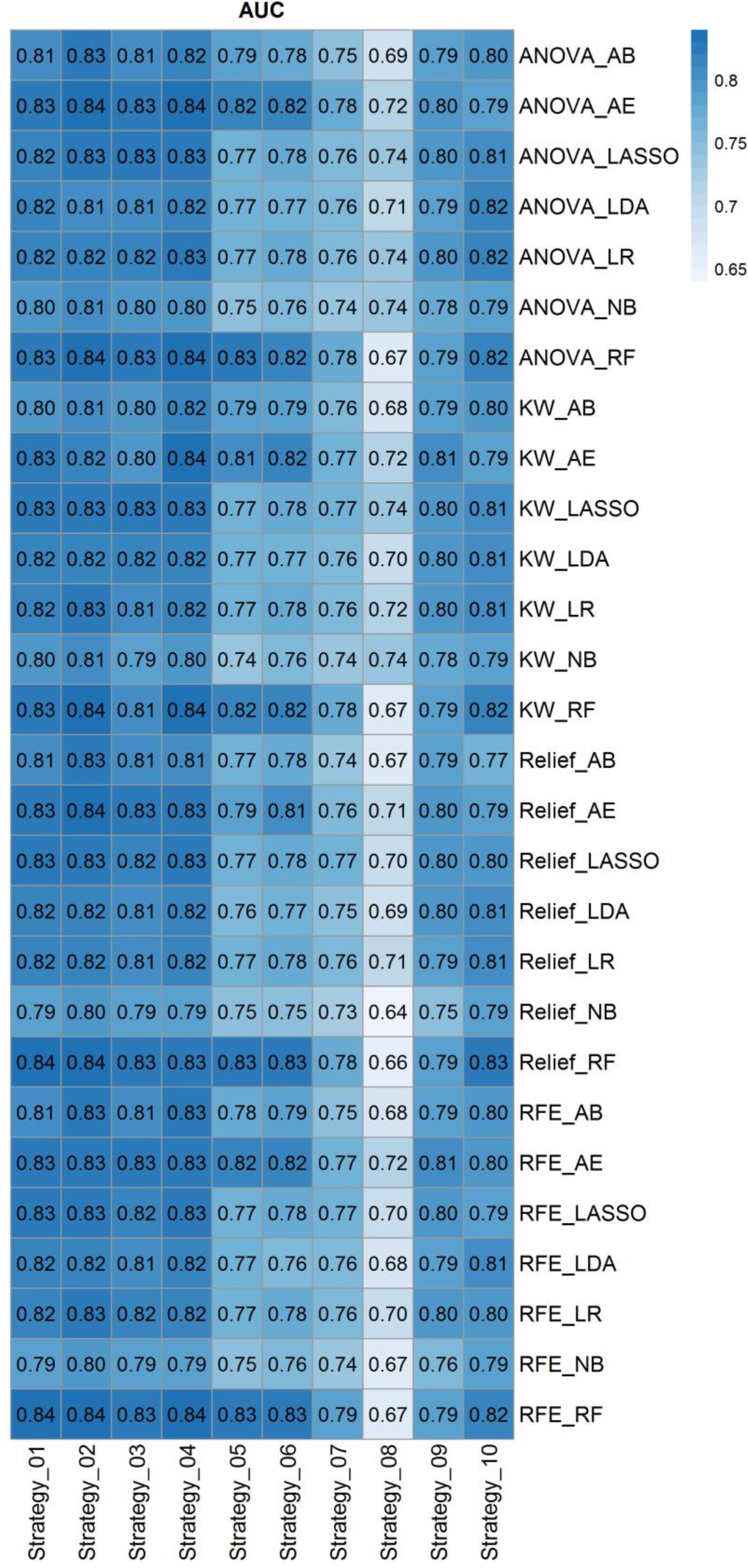
Heat map of AUC for cross combination of feature selectors and classifiers in different ten strategies

**Figure 7:**
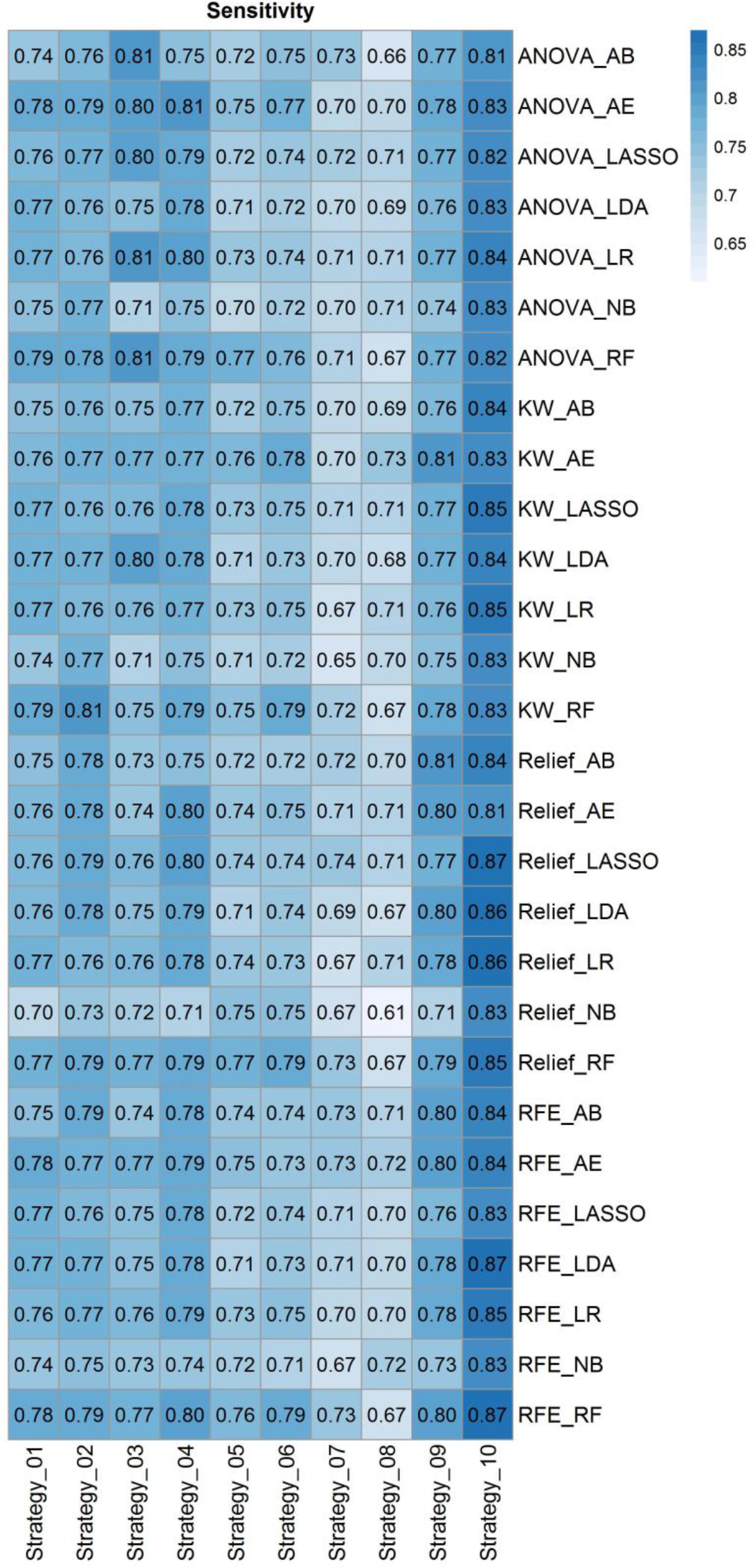
Heat map of Sensitivity for cross combination of feature selectors and classifiers in different ten strategies

**Figure 8:**
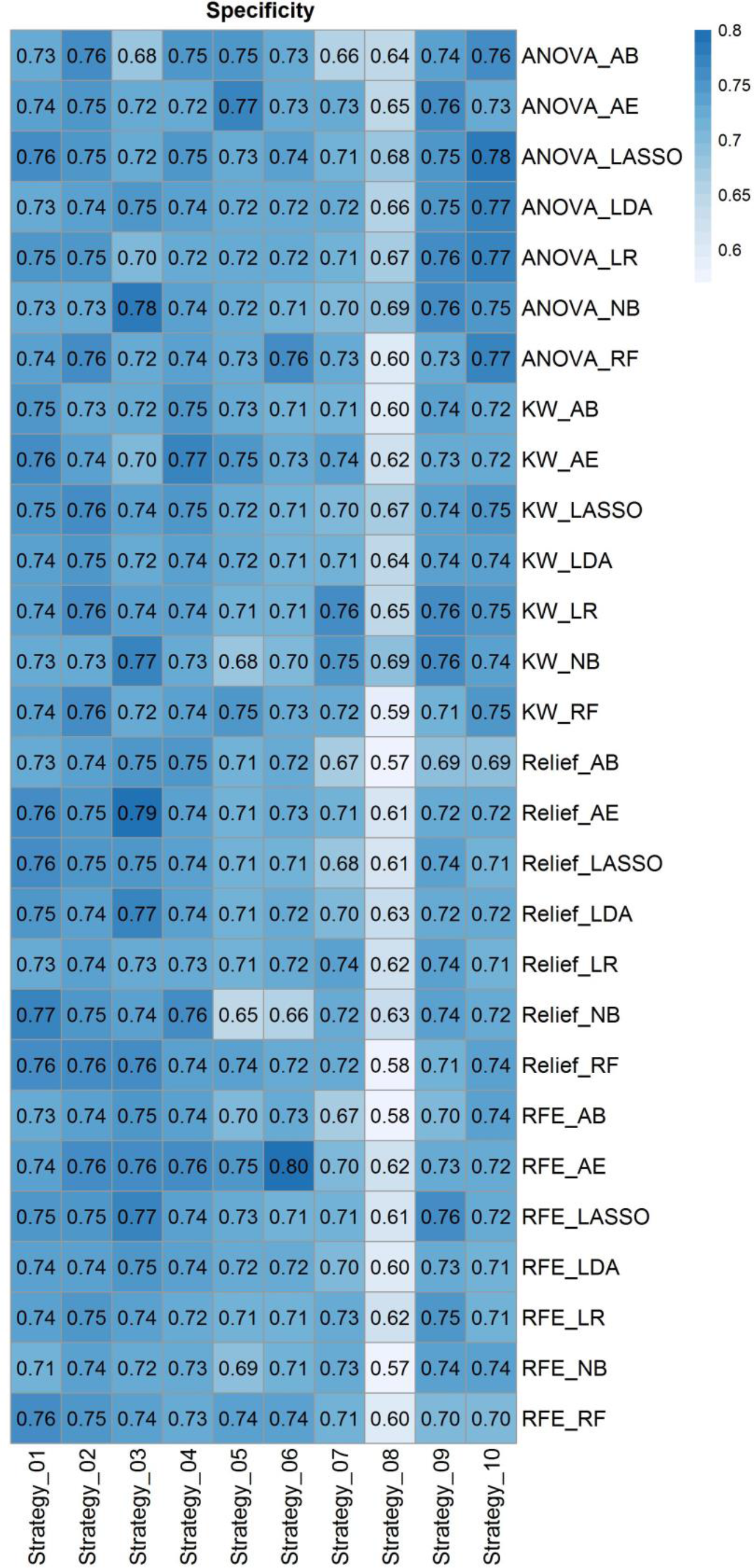
Heat map of Specificity for cross combination of feature selectors and classifiers in different ten strategies

In strategy 1 where the data were randomly split into train and test sets (without considering centers), RFE feature selection and RF classifier results in highest performance of AUC 0.84±0.01 (CI95%: 0.82-0.85) with sensitivity and specificity of 0.78 and 0.76, respectively. In strategy 2 where only PCR positive studies were randomly split into train and test sets (without considering centers), KW feature selection and RF classifier combination resulted in the highest performance with an AUC of 0.84±0.01 (CI95%: 0.82-0.86) and sensitivity and specificity of 0.81 and 0.76, respectively. There was no statistical significant difference between Strategies 1 and 2, the main difference being the inclusion of CT and PCR positive studies in strategy 1 and only PCR positive studies in strategy 2.

In strategy 3 where whole data splitting was performed in each center separately for train and test sets, ANOVA feature selector and RF classifier combination resulted in the highest performance with AUC of 0.83±0.01 (CI95%: 0.81-0.85), sensitivity and specificity of 0.81 and 0.72, respectively. Similar results as above were achieved for strategy, 4 where data splitting was performed in each center separately to train and test set for PCR positive dataset. There were no statistically significant difference between strategies 3 and 4 where the main difference was including CT and PCR positive in strategy 4 and only PCR positive in strategy 5.

In strategy 5 where Combat harmonized whole data splitting was performed in each center separately to the train and test sets, Relief feature selector and RF classifier combination resulted in the highest performance with an AUC of 0.83±0.01 (CI95%: 0.81-0.85), sensitivity and specificity of 0.77 and 0.74, respectively. In strategy 6, where Combat harmonized data splitting was performed in each center separately to the train and test sets for PCR positive studies, Relief feature selector and RF classifier combination resulted in the highest performance with an AUC 0.83±0.01 (CI95%: 0.81-0.84), sensitivity and specificity of 0.79 and 0.72, respectively. . There were no statistically significant differences between strategies 5 and 6. The statistical comparison of AUCs between for ComBat harmonization strategies 5 and 6 and to the same splitting in strategies 3 and 4 using DeLong test didn’t reveal any statistically significant differences.

In strategy 7, where the splitting into train and test sets was performed based on centers (centers appear in training and test sets only once), RFE selector and RF classifier combination resulted in the highest performance with an AUC of 0.79±0.01 (CI95%: 0.76-0.81), sensitivity and specificity of 0.73 and 0.71, respectively. In Figure 9, the ROC curves of the test set for strategies 1-7 as well as the comparison of the different strategies are depicted. In strategy 8 where the model is built based on one center dataset (non-harmonized) and then evaluated on the 18 remaining centers (external validation set), ANOVA feature selector and NB classifier combination resulted in the highest performance with an AUC of 0.74±0.034, sensitivity and specificity of 0.71 ± 0.026 and 0.69 ± 0.033, respectively. The results of each center were presented in supplemental Table 12. To evaluate the model on external validation sets, we reported the results of each center in the LOCO strategy 9. ANOVA feature selector and LR classifier combination resulted in the highest performance with an AUC of 0.80±0.084, sensitivity and specificity of 0.77 ± 0.11 and 0.76 ± 0.075, respectively. In strategy 10, the data from each center (non-harmonized) were randomly split into 70% and 30% for training and test sets, respectively, and the models were built and evaluated on each center separately. ANOVA feature selector and LR classifier combination resulted in the highest performance with an AUC of 0.82±0.10, sensitivity and specificity of 0.84 ± 0.12 and 0.77 ± 0.09, respectively. The results of each center were presented in supplemental Tables 13 and 14 for strategies 9 and 10, respectively.

**Figure 9:**
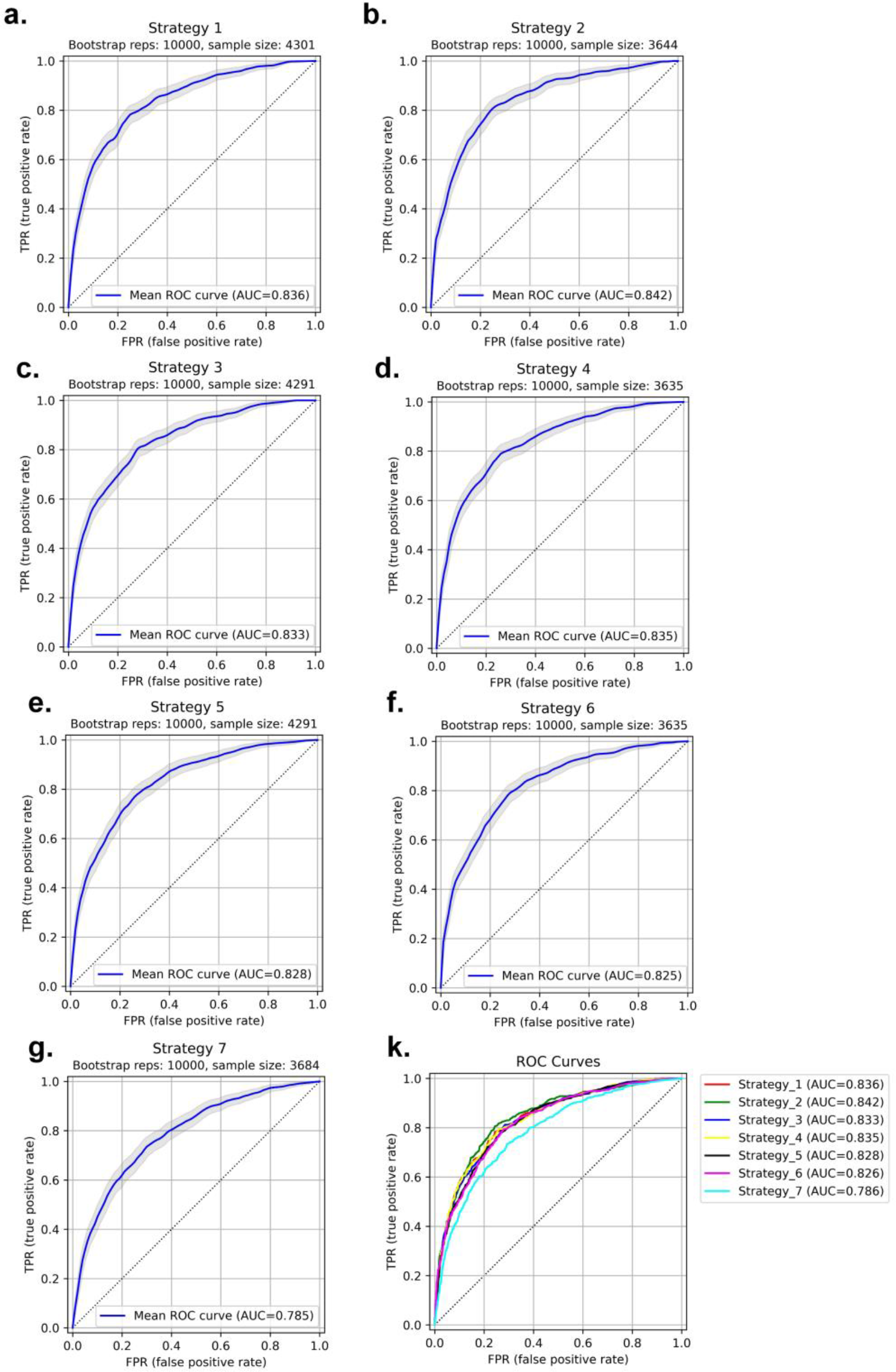
ROC curve for test sets in strategies 1-7. Strategy 1: AUC 0.84±0.01 (a), Strategy 2: AUC 0.84±0.01 (b), Strategy 3: AUC 0.83±0.01 (c), Strategy 4: AUC 0.83±0.01 (d), Strategy 5: AUC 0.83±0.01 (e), Strategy 6: AUC 0.83±0.01 (f), Strategy 7: AUC 0.79±0.01 (g) and different strategies comparison (k)

## DISCUSSION

In this multi-centric study, we conducted a CT-based radiomics analysis to assess the ability of our model in predicting the overall survival of patients with COVID-19 using a large multi-institutional dataset. We included 14,339 patients along with their CT images, segmented the lungs, and extracted distinct radiomics features. We evaluated different combinations of feature selectors and classifiers in different strategies. Since the dataset was gathered from different centers, we applied the ComBat Harmonization algorithm that has been successfully applied in radiomics studies over the extracted features ^57^. As our dataset consisted of imbalanced classes, we first used SMOT algorithm in the training sets. Our model was trained and the results of 3 different testing methods were reported.

Prognostic modeling can be regarded as an important framework towards better understanding of disease, its management, monitoring, and identification of the best treatment options. A number of reports have shown the effectiveness of image-based, laboratory-based, or combined models in outcome prediction of COVID-19 infected patients ^58, 59^. Qiu *et al.* ^60^ constructed a radiomics model trained to classify the severity of COVID-19 lesions (mild vs severe) using CT images. Their study included a medium-to-large number of patients (n=1160) and achieved an AUC of 0.87 in the test dataset. They showed that the radiomics signature is potent in aiding physicians to manage patients in a more precise way. Fu *et al.* ^39^ conducted a similar experiment with a radiomics-based model using CT images and applied it to data from 64 patients to classify them into progressive and stable groups. Their model could accurately perform the given task (AUC=0.83). While the results were promising, their study did not include a large cohort.

A study by Chao *et al.* ^59^ included different types of information, such as CT-based radiomics features, clinical, and demographic data to employ a holistic prognostic model. Their model could predict whether the patients will demand an ICU admission or not with an AUC of 0.88. Tang *et al.* ^61^ also assessed a random forest model for classifying patients into categories of severe and non-severe based on CT imaging radiomics features along with laboratory test results. The model performed well (AUC=0.98) on their dataset consisting of 118 patients. In a study by Wu *et al.* ^62^, the authors assessed the predictive power of a radiomic signature for showing poor patient outcomes defined as ICU admission, need for mechanical ventilation, or death. Their model could reach an AUC of 0.97 in the prediction of 28-day outcomes after CT images were taken. This highly promising result was achieved with the help of clinical data and harmonization of the features. At the same time, in our study, ComBat harmonization did not appear to impact outcome prediction.

One should note that both clinical-only and radiomics-only survival prediction models have advantages. However, studies have shown that radiomics features yield superior accuracy in most cases. In a study by Homayounieh *et al.* ^63^, the authors developed a radiomics-based signature and compared it with a clinical-only signature in terms of mortality prediction. They concluded that radiomics-based model can outperform the clinical-only model with a wide margin (AUC of 0.81 versus 0.68). Their study included 315 adults and was applied to other clinical outcomes as well, such as the prediction of outpatient/inpatient care and ICU admission. In addition, other reports indicated that adding clinical features to the radiomics model only slightly improved the results ^64^. In a recent study, Shiri et al. ^64^ performed a radiomics study for prognostication purpose (alive or deceased) of COVID-19 patients using clinical (demographic, laboratory, and radiological scoring), COVID-19 pneumonia lesion radiomics features and whole lung radiomics features, separately and in combination. They trained a machine learning algorithm, Maximum Relevance Minimum Redundancy (MRMR) as the feature selector and XGBoost as the classifier, on 106 patients and evaluated and reported results on 46 test sets. They reported an AUC of 0.87 ± 0.04 for clinical-only, 0.92 ± 0.03 for whole lung radiomics, 0.92 ± 0.03 for lesion radiomics, 0.91 ± 0.04 for lung + lesion radiomics, 0.92 ± 0.03 for lung radiomics + clinical data, 0.94 ± 0.03 for lesion radiomics + clinical data and 0.95 ± 0.03 for lung + lesion radiomics + clinical data. The lung and lesion radiomics-only models showed similar performance, while the integration of features resulted in the highest accuracy.

Lassau et al. ^65^ combined CT-based DL models, biological and clinical features for severity prediction in 1003 COVID-19 patients, confirmed by either CT or RT-PCR. They showed clinical and biological features correlation with CT markers. Zhang *et al.* ^58^ conducted a diagnostic and prognostic study using 3777 COVID-19 patients. They reported a high positive and negative correlation of lung-lesion CT manifestations with a number of clinical and laboratory tests. They also reported that their diagnostic model (COVID-19 from common pneumonia and normal control) can improve radiologist’s performance from junior to senior level (AUC = 0.98) for progression to severe/critical disease in their prognostic model. They reported an AUC of 0.90 with sensitivity and specificity of 0.80 and 0.86, respectively. Feng *et al.* ^66^ built a machine learning prognostic model using a multicenter COVID-19 dataset. They reported a high correlation of CT features with clinical findings, also utilizing a multivariable model in the validation set consisting of 106 patients. The AUC was 0.89 (95% CI: 0.81–0.98). Recently, Xu et al. ^67^ conducted a multicentric study for the prediction of ICU admission, mechanical ventilation, and mortality of hospitalized patients with COVID-19. CT radiomics features were integrated with demographic and laboratory tests. The evaluation was performed in 1362 patients from nine hospitals reporting an AUC of 0.916, 0.919 and 0.853 for ICU admission, mechanical ventilation, and mortality of hospitalized patients, respectively. For the radiomics-only model, they reached an AUC of 0.86, 0.80, and 0.66 for the above three mentioned outcomes, respectively.

Most previous studies suffered from a common limitation of COVID-19 RT-PCR not being available for the entire dataset when using multicentric data. In our study, COVID-19 positivity was confirmed by either RT-PCR or CT images, and different strategies were adopted to evaluate the models, including random splits and leave-one-center-out. We randomly split the data to train and test sets containing both CT positive and RT-PCR positive patients. Furthermore, to ensure the reproducibility of our results on RT-PCR positive patients, we split the dataset in a way that the test set consisted only of RT-PCR positive patients. To maximize the generalizability of the model and avoid overfitting on training sets, owing to variability in acquisition and reconstruction protocols, our model was developed on multicentric datasets with a wide variety of acquisition and reconstruction parameters. To test the generalizability of our model, we repeated the evaluation of our model using leave-one-center-out cross-validation. The results were reported for 10 different strategies of splitting and cross-validation scenarios.

Several studies reported on the use of CT radiomics or DL algorithms for diagnostic and prognostic purposes in patients with COVID-19 ^58, 59, 68^. However, most studies were performed using a small sample size. Overall, establishing evidence that radiomics features can help prioritize patients based on the severity of their disease and/or predicting their survival requires assessment using larger cohorts for a more generalizable model because of the wide variability in COVID-19 manifestations in different patients. In this study, we provided a large multinational multicentric dataset and evaluated our model in different scenarios to ensure model reproducibility, robustness and generalizability.

While attempting to address bias and limitations to create a generalizable model, the results should be interpreted considering some issues. First, motion artifacts were unavoidable in some COVID-19 patient scans which resulted in overlapping pneumonia regions. We removed patients with severe motion artifacts to omit this effect on model generalization. Second, we enrolled patients with common symptoms of COVID-19 whose infection was confirmed by either RT-PCR or CT imaging (typical manifestation of COVID-19 defined by interim guidelines). We handled this issue by testing different scenarios, including training a model using RT-PCR or CT positive patients and held out only RT-PCR patients in the test set and reported reproducible and repeatable results. Third, we did not include comorbidities (increased risk of adverse outcome), clinical or laboratory data during modeling. However, previous studies showed high correlation of lung features with these findings ^58, 65, 66^. Future studies combining various information to build a holistic model using a large dataset could improve the model’s performance. Forth, we built a prognostic model based on all lung radiomics features. However, COVID-19 can result in imaging manifestations in other organs, such as the heart. Including features from different organs has the potential of improving prognostic performance ^69^. Fifth, therapeutic regimens for different patients were not considered during modeling although providing this information may help improving the accuracy of the model. Sixth, only binary classification was considered for the prognostic model in this study. Future studies should perform survival analysis using time-to-event models to account for the time of adverse event. Lastly, we did not evaluate the impact of image acquisition or reconstruction parameters on radiomics features at the same time. We applied ComBat harmonization algorithm to eliminate center-specific parameter effects on CT radiomics features.

## CONCLUSION

A very large heterogeneous COVID-19 database was gathered from multiple centers and a predictive model of survival outcome derived and extensively tested to evaluate its reproducibility and generalizability. We demonstrated that lung CT radiomics features could be used as biomarkers for prognostic modeling in COVID-19. Through the use of a large imaging dataset, the predictive power of the proposed CT radiomics model is more reliable and may be prospectively used in clinical setting to manage COVID-19 patients.

## Data Availability

All data produced in the present work are contained in the manuscript

## ACKNOWLEDGMENTS

This work was supported by the Swiss National Science Foundation under grant SNRF 320030_176052.

## Data and code availability

Radiomics features and code would be available with request upon publication.

---

TRIPOD Checklist: Prediction Model Development and Validation.
| Section/Topic | Checklist Item |  |  | Page |
| --- | --- | --- | --- | --- |
| Title and abstract |  |  |  |  |
| Title | 1 | D;V | Identify the study as developing and/or validating a multivariable prediction model, the target population, and the outcome to be predicted. | 1 |
| Abstract | 2 | D;V | Provide a summary of objectives, study design, setting, participants, sample size, predictors, outcome, statistical analysis, results, and conclusions. | 1 |
| Introduction |  |  |  |  |
| Background and objectives | 3a | D;V | Explain the medical context (including whether diagnostic or prognostic) and rationale for developing or validating the multivariable prediction model, including references to existing models. | 2-4 |
|  | 3b | D;V | Specify the objectives, including whether the study describes the development or validation of the model or both. | 4 |
| Methods |  |  |  |  |
| Source of data | 4a | D;V | Describe the study design or source of data (e.g., randomized trial, cohort, or registry data), separately for the development and validation data sets, if applicable. | 5 |
|  | 4b | D;V | Specify the key study dates, including start of accrual; end of accrual; and, if applicable, end of follow-up. | 6 |
| Participants | 5a | D;V | Specify key elements of the study setting (e.g., primary care, secondary care, general population) including number and location of centres. | 6 |
|  | 5b | D;V | Describe eligibility criteria for participants. | 5 |
|  | 5c | D;V | Give details of treatments received, if relevant. | 6 |
| Outcome | 6a | D;V | Clearly define the outcome that is predicted by the prediction model, including how and when assessed. | 6 |
|  | 6b | D;V | Report any actions to blind assessment of the outcome to be predicted. | 8-10 |
| Predictors | 7a | D;V | Clearly define all predictors used in developing or validating the multivariable prediction model, including how and when they were measured. | 7 |
|  | 7b | D;V | Report any actions to blind assessment of predictors for the outcome and other predictors. | 8-10 |
| Sample size | 8 | D;V | Explain how the study size was arrived at. | 5-6 |
| Missing data | 9 | D;V | Describe how missing data were handled (e.g., complete-case analysis, single imputation, multiple imputation) with details of any imputation method. | NA |
| Statistical analysis methods | 10a | D | Describe how predictors were handled in the analyses. | 7 |
|  | 10b | D | Specify type of model, all model-building procedures (including any predictor selection), and method for internal validation. | 8-10 |
|  | 10c | V | For validation, describe how the predictions were calculated. | 8-10 |
|  | 10d | D;V | Specify all measures used to assess model performance and, if relevant, to compare multiple models. | 8-10 |
|  | 10e | V | Describe any model updating (e.g., recalibration) arising from the validation, if done. | 8-9 |
| Risk groups | 11 | D;V | Provide details on how risk groups were created, if done. | 6-7 |
| Development vs. validation | 12 | V | For validation, identify any differences from the development data in setting, eligibility criteria, outcome, and predictors. | 8-9<br>Fig. 3 |
| Results |  |  |  |  |
| Participants | 13a | D;V | Describe the flow of participants through the study, including the number of participants with and without the outcome and, if applicable, a summary of the follow-up time. A diagram may be helpful. | 5-6<br>Fig. 2 |
|  | 13b | D;V | Describe the characteristics of the participants (basic demographics, clinical features, available predictors), including the number of participants with missing data for predictors and outcome. Not mentioned in result | 5-6<br>Table 1 |
|  | 13c | V | For validation, show a comparison with the development data of the distribution of important variables (demographics, predictors and outcome). | 10-12 |
| Model development | 14a | D | Specify the number of participants and outcome events in each analysis. | 5-6<br>Fig. 2 |
|  | 14b | D | If done, report the unadjusted association between each candidate predictor and outcome. | 10 |
| Model specification | 15a | D | Present the full prediction model to allow predictions for individuals (i.e., all regression coefficients, and model intercept or baseline survival at a given time point). | NA |
|  | 15b | D | Explain how to use the prediction model. | 10-12 |
| Model performance | 16 | D;V | Report performance measures (with CIs) for the prediction model. | 10-12 |
| Model-updating | 17 | V | If done, report the results from any model updating (i.e., model specification, model performance). | 10-11 |
| **Discussion** |  |  |  |  |
| Limitations | 18 | D;V | Discuss any limitations of the study (such as nonrepresentative sample, few events per predictor, missing data). | 15 |
| Interpretation | 19a | V | For validation, discuss the results with reference to performance in the development data, and any other validation data. | 12-14 |
|  | 19b | D;V | Give an overall interpretation of the results, considering objectives, limitations, results from similar studies, and other relevant evidence. | 14-15 |
| Implications | 20 | D;V | Discuss the potential clinical use of the model and implications for future research. | 15-16 |
| **Other information** |  |  |  |  |
| Supplementary information | 21 | D;V | Provide information about the availability of supplementary resources, such as study protocol, Web calculator, and data sets. | 16 |
| Funding | 22 | D;V | Give the source of funding and the role of the funders for the present study. | 16 |

\*Items relevant only to the development of a prediction model are denoted by D, items relating solely to a validation of a prediction model are denoted by V, and items relating to both are denoted D;V. We recommend using the TRIPOD Checklist in conjunction with the TRIPOD Explanation and Elaboration document.

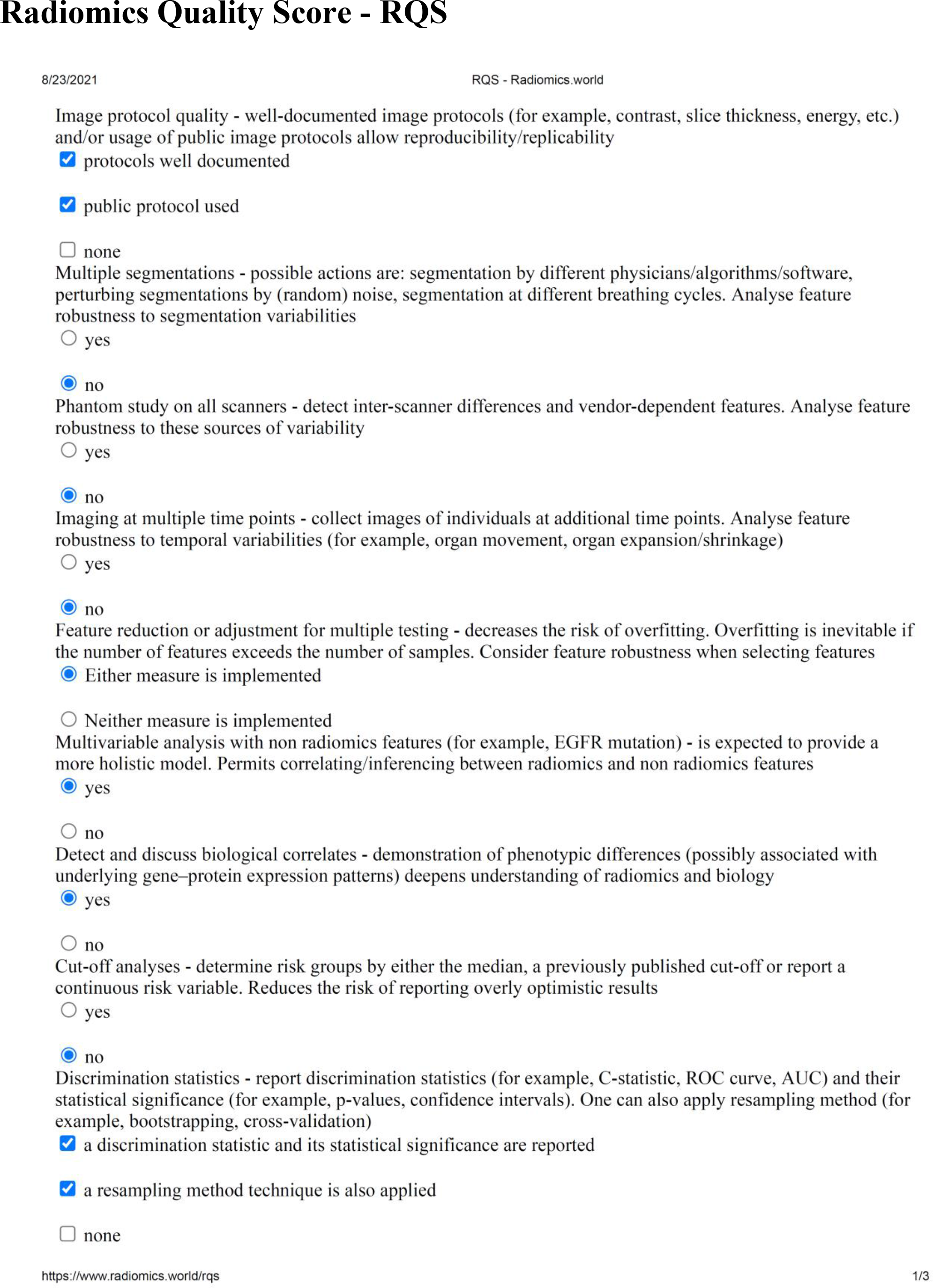
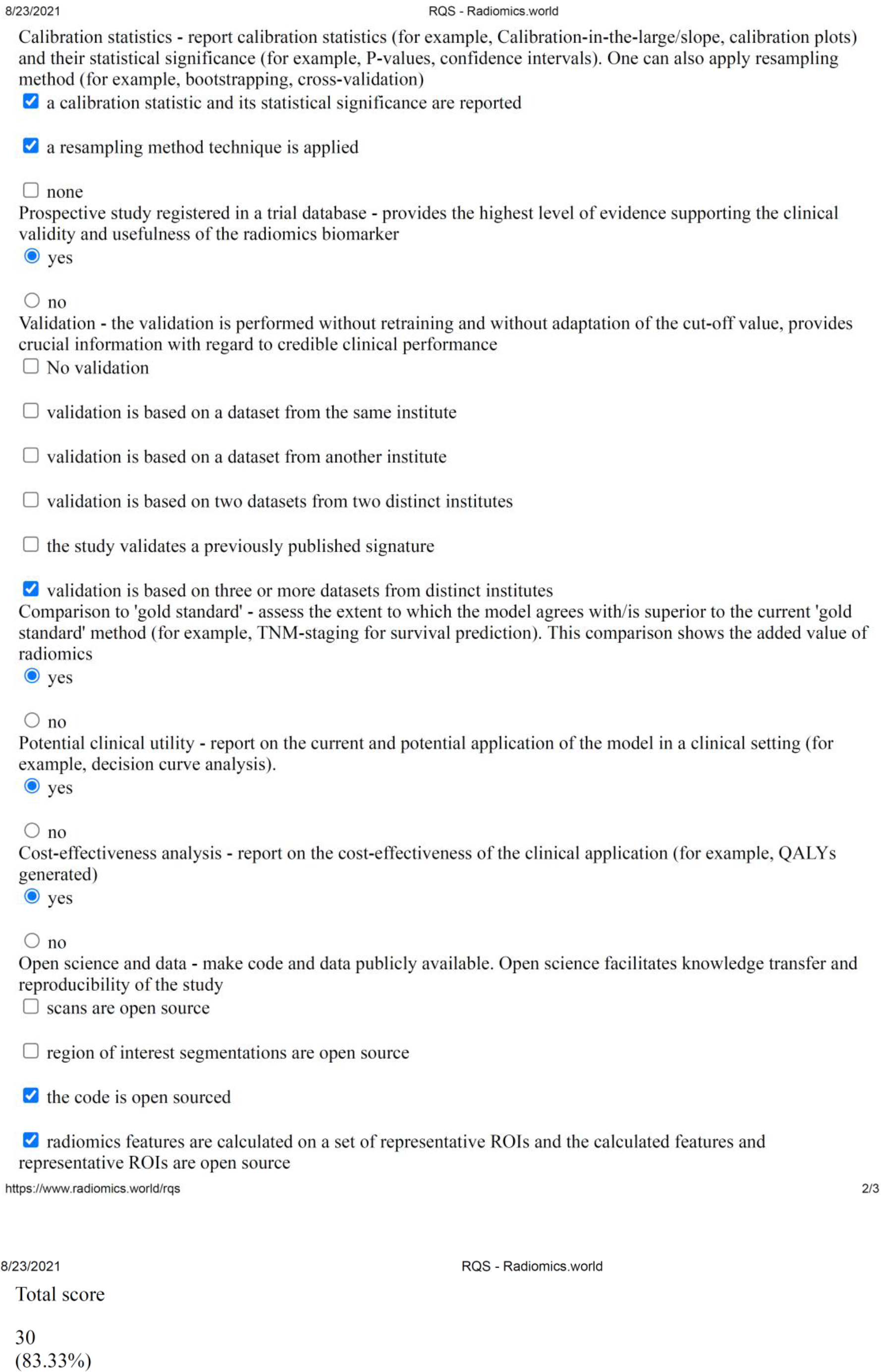

CLAIM: Checklist for Artificial Intelligence in Medical Imaging
| Section / Topic | No. | Item |  |
| --- | --- | --- | --- |
| **TITLE / ABSTRACT** |  |  |  |
|  | 1 | Identification as a study of AI methodology, specifying the category of technology used (e.g., deep learning) | ✓ |
|  | 2 | Structured summary of study design, methods, results, and conclusions | ✓ |
| **INTRODUCTION** |  |  |  |
|  | 3 | Scientific and clinical background, including the intended use and clinical role of the AI approach | ✓ |
|  | 4 | Study objectives and hypotheses | ✓ |
| **METHODS** |  |  |  |
| *Study Design* | 5 | Prospective or retrospective study | ✓ |
|  | 6 | Study goal, such as model creation, exploratory study, feasibility study, non-inferiority trial | ✓ |
| *Data* | 7 | Data sources | ✓ |
|  | 8 | Eligibility criteria: how, where, and when potentially eligible participants or studies were identified (e.g., symptoms, results from previous tests, inclusion in registry, patient-care setting, location, dates) | ✓ |
|  | 9 | Data pre-processing steps | ✓ |
|  | 10 | Selection of data subsets, if applicable | ✓ |
|  | 11 | Definitions of data elements, with references to Common Data Elements | ✓ |
|  | 12 | De-identification methods | ✓ |
|  | 13 | How missing data were handled | ✓ |
| *Ground Truth* | 14 | Definition of ground truth reference standard, in sufficient detail to allow replication | ✓ |
|  | 15 | Rationale for choosing the reference standard (if alternatives exist) | ✓ |
|  | 16 | Source of ground-truth annotations; qualifications and preparation of annotators | ✓ |
|  | 17 | Annotation tools | ✓ |
|  | 18 | Measurement of inter- and intrarater variability; methods to mitigate variability and/or resolve discrepancies | NA |
| *Data Partitions* | 19 | Intended sample size and how it was determined | ✓ |
|  | 20 | How data were assigned to partitions; specify proportions | ✓ |
|  | 21 | Level at which partitions are disjoint (e.g., image, study, patient, institution) | ✓ |
| *Model* | 22 | Detailed description of model, including inputs, outputs, all intermediate layers and connections | ✓ |
|  | 23 | Software libraries, frameworks, and packages | ✓ |
|  | 24 | Initialization of model parameters (e.g., randomization, transfer learning) | ✓ |
| *Training* | 25 | Details of training approach, including data augmentation, hyperparameters, number of models trained | ✓ |
|  | 26 | Method of selecting the final model | ✓ |
|  | 27 | Ensembling techniques, if applicable | ✓ |
| *Evaluation* | 28 | Metrics of model performance | ✓ |
|  | 29 | Statistical measures of significance and uncertainty (e.g., confidence intervals) | ✓ |
|  | 30 | Robustness or sensitivity analysis | ✓ |
|  | 31 | Methods for explainability or interpretability (e.g., saliency maps), and how they were validated | NA |
|  | 32 | Validation or testing on external data | ✓ |
| **RESULTS** |  |  |  |
| *Data* | 33 | Flow of participants or cases, using a diagram to indicate inclusion and exclusion | ✓ |
|  | 34 | Demographic and clinical characteristics of cases in each partition | ✓ |
| *Model performance* | 35 | Performance metrics for optimal model(s) on all data partitions | ✓ |
|  | 36 | Estimates of diagnostic accuracy and their precision (such as 95% confidence intervals) | ✓ |
|  | 37 | Failure analysis of incorrectly classified cases | NA |
| **DISCUSSION** |  |  |  |
|  | 38 | Study limitations, including potential bias, statistical uncertainty, and generalizability | ✓ |
|  | 39 | Implications for practice, including the intended use and/or clinical role | ✓ |
| **OTHER INFORMATION** |  |  |  |
|  | 40 | Registration number and name of registry | ✓ |
|  | 41 | Where the full study protocol can be accessed | ✓ |
|  | 42 | Sources of funding and other support; role of funders | ✓ |

**Supplemental Figure 1.**
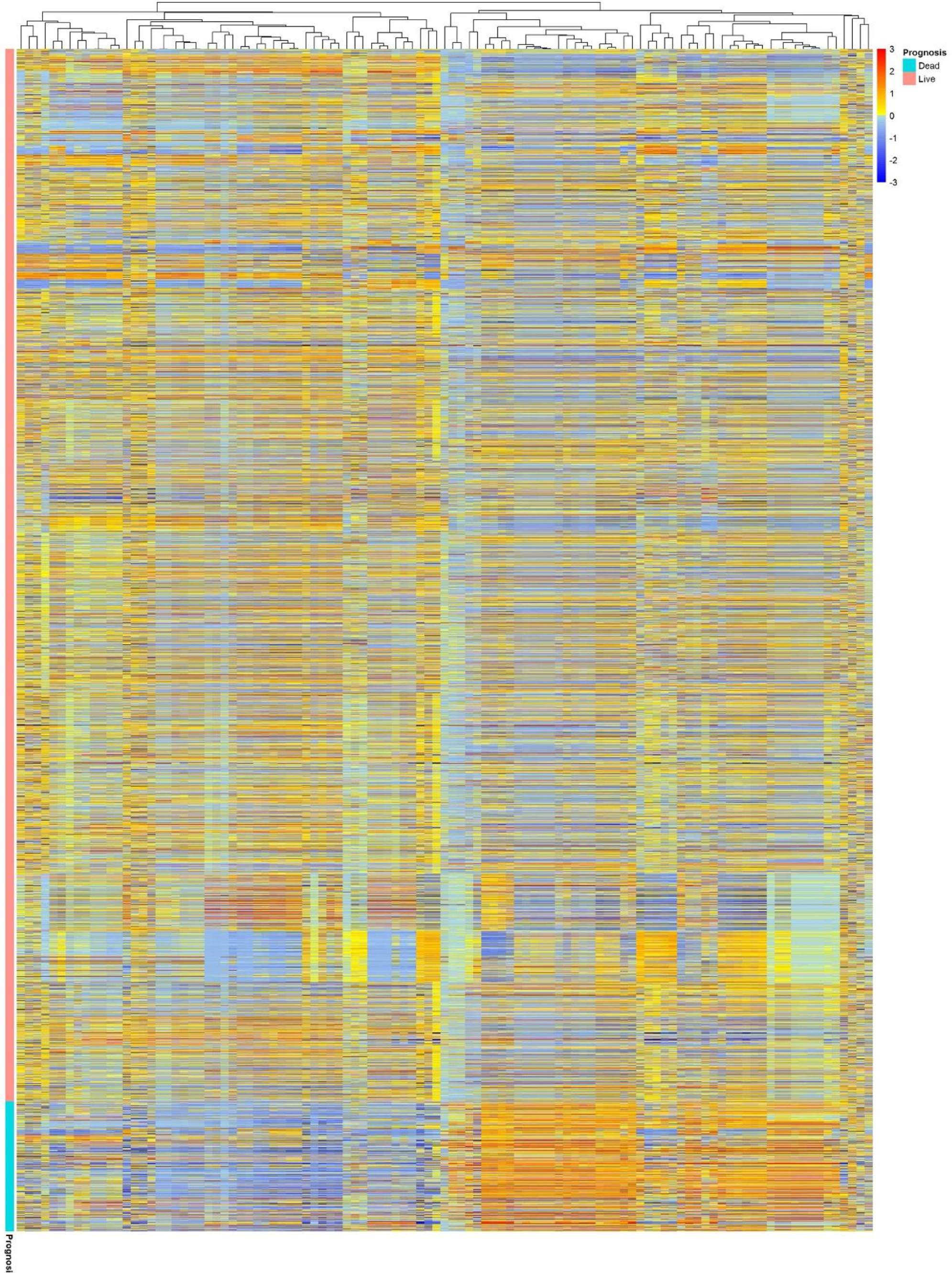
Cluster heat map of radiomics features in the non-harmonized data set.

**Supplemental Figure 2.**
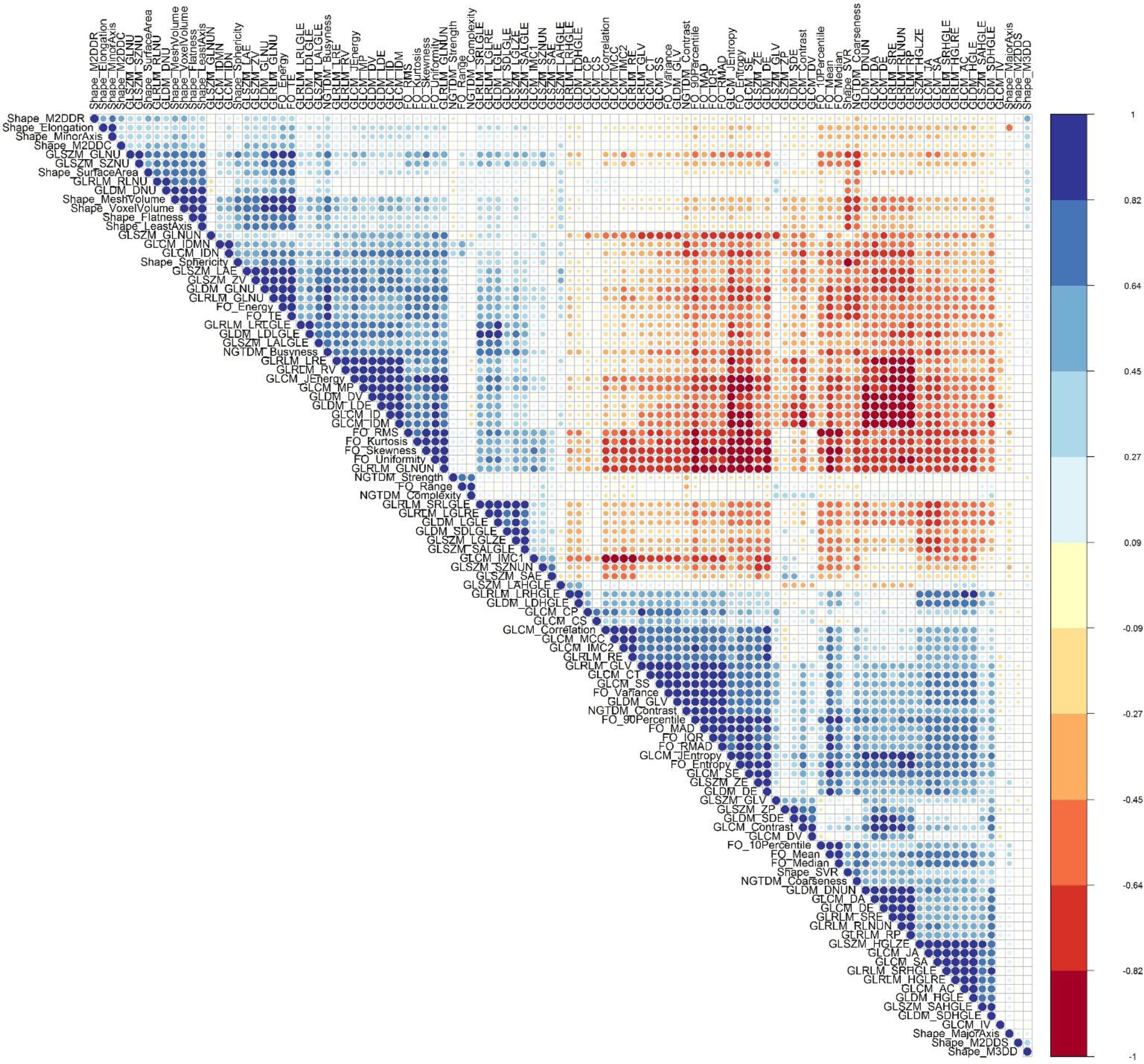
Pearson correlation of ComBat-harmonized radiomics features.

**Supplemental Table 1.**
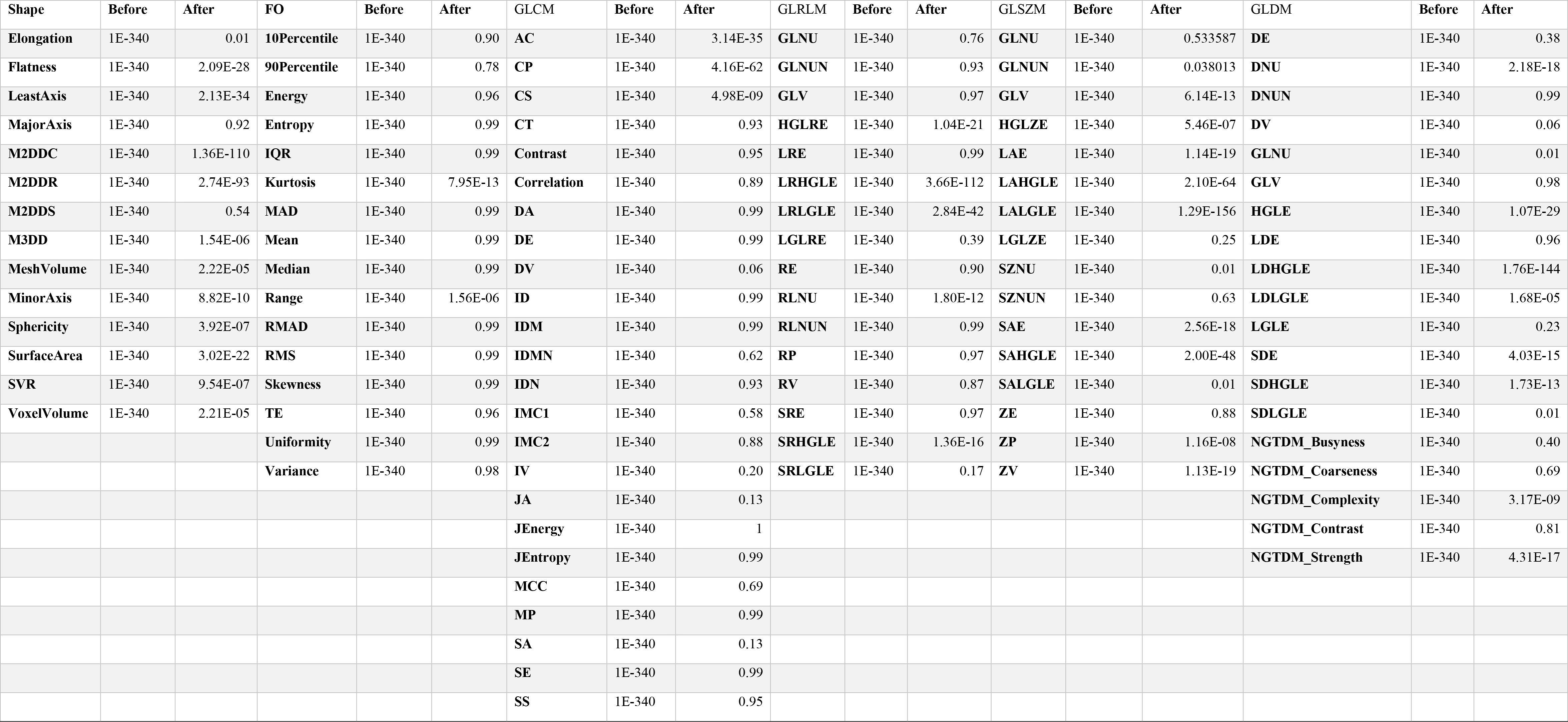
P-values in Kruskal Wallis analysis of radiomics features before and after ComBat Harmonization.

**Supplemental Table 2.** Classification performance indices for different feature selectors (FS) and classifiers in Strategy 1.

| FS | Classifier | AUC | 95% CIs | Std | Acc | Sen | Spe |
| --- | --- | --- | --- | --- | --- | --- | --- |
| **ANOVA** | RF | 0.83 | [0.81-0.85] | 0.010 | 0.74 | 0.79 | 0.74 |
|  | AB | 0.81 | [0.79-0.83] | 0.011 | 0.73 | 0.74 | 0.73 |
|  | AE | 0.83 | [0.81-0.85] | 0.010 | 0.74 | 0.78 | 0.74 |
|  | LDA | 0.82 | [0.80-0.84] | 0.011 | 0.73 | 0.77 | 0.73 |
|  | LR | 0.82 | [0.80-0.84] | 0.010 | 0.75 | 0.77 | 0.75 |
|  | LASSO | 0.82 | [0.80-0.84] | 0.010 | 0.76 | 0.76 | 0.76 |
|  | NB | 0.80 | [0.78-0.82] | 0.011 | 0.73 | 0.75 | 0.73 |
| **KW** | RF | 0.83 | [0.81-0.85] | 0.010 | 0.74 | 0.79 | 0.74 |
|  | AB | 0.80 | [0.78-0.82] | 0.011 | 0.75 | 0.75 | 0.75 |
|  | AE | 0.83 | [0.81-0.85] | 0.010 | 0.76 | 0.76 | 0.76 |
|  | LDA | 0.82 | [0.80-0.84] | 0.011 | 0.74 | 0.77 | 0.74 |
|  | LR | 0.82 | [0.80-0.84] | 0.010 | 0.75 | 0.77 | 0.74 |
|  | LASSO | 0.83 | [0.81-0.84] | 0.010 | 0.75 | 0.77 | 0.75 |
|  | NB | 0.80 | [0.78-0.82] | 0.011 | 0.73 | 0.74 | 0.73 |
| **Relief** | RF | 0.84 | [0.82-0.85] | 0.010 | 0.76 | 0.77 | 0.76 |
|  | AB | 0.81 | [0.79-0.83] | 0.010 | 0.73 | 0.75 | 0.73 |
|  | AE | 0.83 | [0.81-0.84] | 0.010 | 0.76 | 0.76 | 0.76 |
|  | LDA | 0.82 | [0.79-0.84] | 0.011 | 0.75 | 0.76 | 0.75 |
|  | LR | 0.82 | [0.80-0.84] | 0.010 | 0.73 | 0.77 | 0.73 |
|  | LASSO | 0.83 | [0.81-0.85] | 0.010 | 0.76 | 0.76 | 0.76 |
|  | NB | 0.79 | [0.77-0.82] | 0.011 | 0.76 | 0.70 | 0.77 |
| **RFE** | RF | 0.84 | [0.82-0.85] | 0.010 | 0.76 | 0.78 | 0.76 |
|  | AB | 0.81 | [0.79-0.83] | 0.010 | 0.73 | 0.75 | 0.73 |
|  | AE | 0.83 | [0.81-0.85] | 0.010 | 0.74 | 0.78 | 0.74 |
|  | LDA | 0.82 | [0.79-0.84] | 0.011 | 0.74 | 0.77 | 0.74 |
|  | LR | 0.82 | [0.80-0.84] | 0.010 | 0.74 | 0.76 | 0.74 |
|  | LASSO | 0.83 | [0.81-0.85] | 0.010 | 0.75 | 0.77 | 0.75 |
|  | NB | 0.79 | [0.77-0.81] | 0.011 | 0.71 | 0.74 | 0.71 |

**Supplemental Table 3.** Classification performance indices for different feature selectors (FS) and classifiers in Strategy 2.

| FS | Classifier | AUC | 95% CIs | Std | Acc | Sen | Spe |
| --- | --- | --- | --- | --- | --- | --- | --- |
| **ANOVA** | RF | 0.84 | [0.82-0.86] | 0.011 | 0.76 | 0.78 | 0.76 |
|  | AB | 0.83 | [0.80-0.85] | 0.010 | 0.76 | 0.76 | 0.76 |
|  | AE | 0.84 | [0.82-0.86] | 0.010 | 0.75 | 0.79 | 0.75 |
|  | LDA | 0.81 | [0.79-0.84] | 0.011 | 0.74 | 0.76 | 0.74 |
|  | LR | 0.82 | [0.80-0.84] | 0.010 | 0.75 | 0.76 | 0.75 |
|  | LASSO | 0.83 | [0.81-0.85] | 0.010 | 0.75 | 0.77 | 0.75 |
|  | NB | 0.81 | [0.78-0.83] | 0.011 | 0.73 | 0.77 | 0.73 |
| **KW** | RF | 0.84 | [0.82-0.86] | 0.010 | 0.76 | 0.81 | 0.76 |
|  | AB | 0.81 | [0.79-0.83] | 0.011 | 0.73 | 0.76 | 0.73 |
|  | AE | 0.82 | [0.80-0.84] | 0.010 | 0.74 | 0.77 | 0.74 |
|  | LDA | 0.82 | [0.80-0.84] | 0.011 | 0.75 | 0.77 | 0.75 |
|  | LR | 0.83 | [0.80-0.85] | 0.010 | 0.76 | 0.76 | 0.76 |
|  | LASSO | 0.83 | [0.80-0.85] | 0.011 | 0.76 | 0.76 | 0.76 |
|  | NB | 0.81 | [0.78-0.83] | 0.011 | 0.74 | 0.77 | 0.73 |
| **Relief** | RF | 0.84 | [0.82-0.86] | 0.011 | 0.77 | 0.79 | 0.76 |
|  | AB | 0.83 | [0.81-0.85] | 0.010 | 0.75 | 0.78 | 0.74 |
|  | AE | 0.84 | [0.82-0.86] | 0.010 | 0.75 | 0.78 | 0.75 |
|  | LDA | 0.82 | [0.80-0.84] | 0.011 | 0.75 | 0.78 | 0.74 |
|  | LR | 0.82 | [0.79-0.84] | 0.011 | 0.74 | 0.76 | 0.74 |
|  | LASSO | 0.83 | [0.81-0.85] | 0.010 | 0.76 | 0.79 | 0.75 |
|  | NB | 0.80 | [0.78-0.82] | 0.012 | 0.75 | 0.73 | 0.75 |
| **RFE** | RF | 0.84 | [0.82-0.86] | 0.011 | 0.75 | 0.79 | 0.75 |
|  | AB | 0.83 | [0.81-0.85] | 0.010 | 0.74 | 0.79 | 0.74 |
|  | AE | 0.83 | [0.81-0.85] | 0.010 | 0.76 | 0.77 | 0.76 |
|  | LDA | 0.82 | [0.80-0.84] | 0.011 | 0.74 | 0.77 | 0.74 |
|  | LR | 0.83 | [0.81-0.85] | 0.010 | 0.75 | 0.77 | 0.75 |
|  | LASSO | 0.83 | [0.81-0.85] | 0.010 | 0.75 | 0.76 | 0.75 |
|  | NB | 0.80 | [0.78-0.83] | 0.012 | 0.75 | 0.75 | 0.74 |

**Supplemental Table 4.**
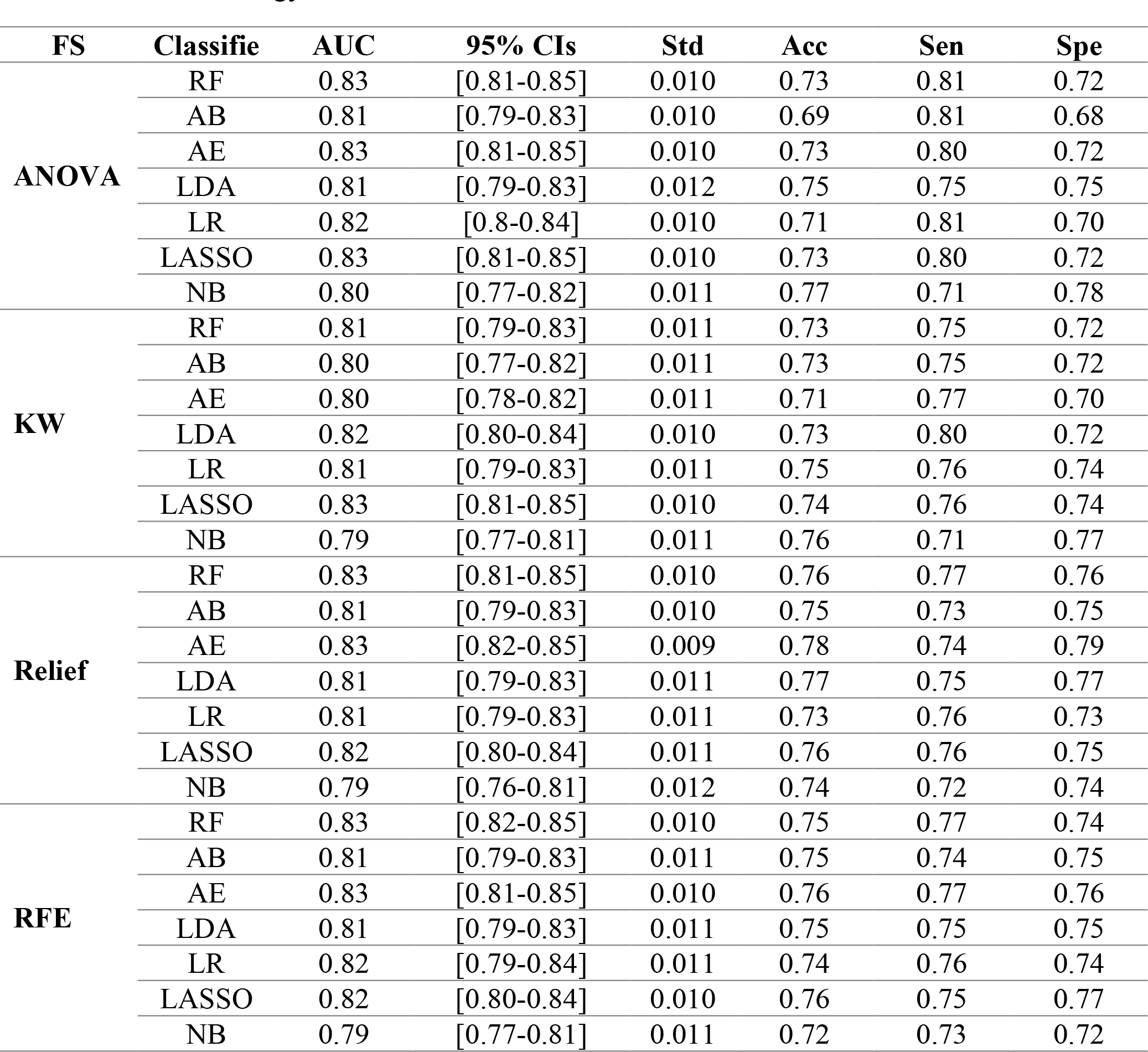
Classification performance indices for different feature selectors (FS) and classifiers in Strategy 3.

| **FS** | **Classifie** | **AUC** | **95% CIs** | **Std** | **Acc** | **Sen** | **Spe** |
| --- | --- | --- | --- | --- | --- | --- | --- |
| **ANOVA** | RF | 0.83 | [0.81-0.85] | 0.010 | 0.73 | 0.81 | 0.72 |
|  | AB | 0.81 | [0.79-0.83] | 0.010 | 0.69 | 0.81 | 0.68 |
|  | AE | 0.83 | [0.81-0.85] | 0.010 | 0.73 | 0.80 | 0.72 |
|  | LDA | 0.81 | [0.79-0.83] | 0.012 | 0.75 | 0.75 | 0.75 |
|  | LR | 0.82 | [0.8-0.84] | 0.010 | 0.71 | 0.81 | 0.70 |
|  | LASSO | 0.83 | [0.81-0.85] | 0.010 | 0.73 | 0.80 | 0.72 |
|  | NB | 0.80 | [0.77-0.82] | 0.011 | 0.77 | 0.71 | 0.78 |
| **KW** | RF | 0.81 | [0.79-0.83] | 0.011 | 0.73 | 0.75 | 0.72 |
|  | AB | 0.80 | [0.77-0.82] | 0.011 | 0.73 | 0.75 | 0.72 |
|  | AE | 0.80 | [0.78-0.82] | 0.011 | 0.71 | 0.77 | 0.70 |
|  | LDA | 0.82 | [0.80-0.84] | 0.010 | 0.73 | 0.80 | 0.72 |
|  | LR | 0.81 | [0.79-0.83] | 0.011 | 0.75 | 0.76 | 0.74 |
|  | LASSO | 0.83 | [0.81-0.85] | 0.010 | 0.74 | 0.76 | 0.74 |
|  | NB | 0.79 | [0.77-0.81] | 0.011 | 0.76 | 0.71 | 0.77 |
| **Relief** | RF | 0.83 | [0.81-0.85] | 0.010 | 0.76 | 0.77 | 0.76 |
|  | AB | 0.81 | [0.79-0.83] | 0.010 | 0.75 | 0.73 | 0.75 |
|  | AE | 0.83 | [0.82-0.85] | 0.009 | 0.78 | 0.74 | 0.79 |
|  | LDA | 0.81 | [0.79-0.83] | 0.011 | 0.77 | 0.75 | 0.77 |
|  | LR | 0.81 | [0.79-0.83] | 0.011 | 0.73 | 0.76 | 0.73 |
|  | LASSO | 0.82 | [0.80-0.84] | 0.011 | 0.76 | 0.76 | 0.75 |
|  | NB | 0.79 | [0.76-0.81] | 0.012 | 0.74 | 0.72 | 0.74 |
| **RFE** | RF | 0.83 | [0.82-0.85] | 0.010 | 0.75 | 0.77 | 0.74 |
|  | AB | 0.81 | [0.79-0.83] | 0.011 | 0.75 | 0.74 | 0.75 |
|  | AE | 0.83 | [0.81-0.85] | 0.010 | 0.76 | 0.77 | 0.76 |
|  | LDA | 0.81 | [0.79-0.83] | 0.011 | 0.75 | 0.75 | 0.75 |
|  | LR | 0.82 | [0.79-0.84] | 0.011 | 0.74 | 0.76 | 0.74 |
|  | LASSO | 0.82 | [0.80-0.84] | 0.010 | 0.76 | 0.75 | 0.77 |
|  | NB | 0.79 | [0.77-0.81] | 0.011 | 0.72 | 0.73 | 0.72 |

**Supplemental Table 5.** Classification performance indices for different feature selectors (FS) and classifiers in Strategy 4.

| FS | Classifier | AUC | 95% CIs | Std | Acc | Sen | Spe |
| --- | --- | --- | --- | --- | --- | --- | --- |
| **ANOVA** | RF | 0.84 | [0.82-0.85] | 0.010 | 0.7477 | 0.79 | 0.74 |
|  | AB | 0.82 | [0.80-0.84] | 0.010 | 0.7483 | 0.75 | 0.75 |
|  | AE | 0.84 | [0.82-0.86] | 0.009 | 0.7354 | 0.81 | 0.72 |
|  | LDA | 0.82 | [0.80-0.84] | 0.011 | 0.7458 | 0.78 | 0.74 |
|  | LR | 0.83 | [0.81-0.85] | 0.010 | 0.7285 | 0.80 | 0.72 |
|  | LASSO | 0.83 | [0.81-0.85] | 0.010 | 0.751 | 0.79 | 0.75 |
|  | NB | 0.80 | [0.78-0.82] | 0.011 | 0.7389 | 0.75 | 0.74 |
| **KW** | RF | 0.84 | [0.82-0.85] | 0.010 | 0.7491 | 0.79 | 0.74 |
|  | AB | 0.82 | [0.80-0.84] | 0.010 | 0.7497 | 0.77 | 0.75 |
|  | AE | 0.84 | [0.82-0.86] | 0.009 | 0.7684 | 0.77 | 0.77 |
|  | LDA | 0.82 | [0.80-0.84] | 0.011 | 0.7461 | 0.78 | 0.74 |
|  | LR | 0.82 | [0.80-0.84] | 0.010 | 0.7461 | 0.77 | 0.74 |
|  | LASSO | 0.83 | [0.81-0.85] | 0.010 | 0.7574 | 0.78 | 0.75 |
|  | NB | 0.80 | [0.78-0.82] | 0.011 | 0.734 | 0.75 | 0.73 |
| **Relief** | RF | 0.83 | [0.81-0.85] | 0.010 | 0.7475 | 0.79 | 0.74 |
|  | AB | 0.81 | [0.79-0.83] | 0.010 | 0.7494 | 0.75 | 0.75 |
|  | AE | 0.83 | [0.81-0.85] | 0.010 | 0.7494 | 0.80 | 0.74 |
|  | LDA | 0.82 | [0.80-0.84] | 0.011 | 0.7428 | 0.79 | 0.74 |
|  | LR | 0.82 | [0.80-0.84] | 0.010 | 0.7409 | 0.78 | 0.73 |
|  | LASSO | 0.83 | [0.81-0.85] | 0.010 | 0.7466 | 0.81 | 0.74 |
|  | NB | 0.79 | [0.76-0.81] | 0.011 | 0.7494 | 0.71 | 0.76 |
| **RFE** | RF | 0.84 | [0.82-0.85] | 0.010 | 0.7384 | 0.80 | 0.73 |
|  | AB | 0.83 | [0.81-0.84] | 0.010 | 0.7422 | 0.78 | 0.74 |
|  | AE | 0.83 | [0.81-0.85] | 0.010 | 0.7618 | 0.79 | 0.76 |
|  | LDA | 0.82 | [0.80-0.84] | 0.011 | 0.7409 | 0.78 | 0.74 |
|  | LR | 0.82 | [0.80-0.84] | 0.010 | 0.7329 | 0.79 | 0.72 |
|  | LASSO | 0.83 | [0.81-0.85] | 0.010 | 0.7483 | 0.78 | 0.74 |
|  | NB | 0.79 | [0.77-0.81] | 0.011 | 0.7359 | 0.74 | 0.73 |

**Supplemental Table 6.** Classification performance indices for different feature selectors (FS) and classifiers in Strategy 5.

| FS | Classifier | AUC | 95% CIs | Std | Acc | Sen | Spe |
| --- | --- | --- | --- | --- | --- | --- | --- |
| **ANOVA** | RF | 0.83 | [0.81-0.85] | 0.010 | 0.73 | 0.77 | 0.73 |
|  | AB | 0.79 | [0.76-0.81] | 0.012 | 0.74 | 0.72 | 0.75 |
|  | AE | 0.82 | [0.79-0.84] | 0.011 | 0.76 | 0.75 | 0.77 |
|  | LDA | 0.77 | [0.74-0.79] | 0.013 | 0.72 | 0.71 | 0.72 |
|  | LR | 0.77 | [0.75-0.80] | 0.012 | 0.72 | 0.73 | 0.72 |
|  | LASSO | 0.77 | [0.75-0.80] | 0.012 | 0.73 | 0.72 | 0.73 |
|  | NB | 0.75 | [0.72-0.77] | 0.013 | 0.72 | 0.70 | 0.72 |
| **KW** | RF | 0.82 | [0.80-0.84] | 0.010 | 0.75 | 0.75 | 0.75 |
|  | AB | 0.79 | [0.76-0.81] | 0.012 | 0.73 | 0.72 | 0.73 |
|  | AE | 0.81 | [0.79-0.83] | 0.011 | 0.75 | 0.76 | 0.75 |
|  | LDA | 0.77 | [0.74-0.79] | 0.013 | 0.72 | 0.71 | 0.72 |
|  | LR | 0.77 | [0.75-0.80] | 0.012 | 0.71 | 0.73 | 0.71 |
|  | LASSO | 0.77 | [0.75-0.80] | 0.012 | 0.72 | 0.73 | 0.72 |
|  | NB | 0.74 | [0.71-0.76] | 0.013 | 0.68 | 0.71 | 0.68 |
| **Relief** | RF | 0.83 | [0.81-0.85] | 0.010 | 0.74 | 0.77 | 0.74 |
|  | AB | 0.77 | [0.74-0.79] | 0.012 | 0.71 | 0.72 | 0.71 |
|  | AE | 0.79 | [0.77-0.81] | 0.012 | 0.72 | 0.74 | 0.71 |
|  | LDA | 0.76 | [0.74-0.78] | 0.012 | 0.71 | 0.71 | 0.71 |
|  | LR | 0.77 | [0.75-0.79] | 0.012 | 0.71 | 0.74 | 0.71 |
|  | LASSO | 0.77 | [0.75-0.80] | 0.012 | 0.71 | 0.74 | 0.71 |
|  | NB | 0.75 | [0.72-0.77] | 0.013 | 0.66 | 0.75 | 0.65 |
| **RFE** | RF | 0.83 | [0.81-0.85] | 0.010 | 0.74 | 0.76 | 0.74 |
|  | AB | 0.78 | [0.76-0.80] | 0.012 | 0.71 | 0.74 | 0.70 |
|  | AE | 0.82 | [0.80-0.84] | 0.011 | 0.75 | 0.75 | 0.75 |
|  | LDA | 0.77 | [0.74-0.79] | 0.013 | 0.72 | 0.71 | 0.72 |
|  | LR | 0.77 | [0.75-0.80] | 0.012 | 0.71 | 0.73 | 0.71 |
|  | LASSO | 0.77 | [0.75-0.80] | 0.012 | 0.73 | 0.72 | 0.73 |
|  | NB | 0.75 | [0.72-0.77] | 0.013 | 0.70 | 0.72 | 0.69 |

**Supplemental Table 7.**
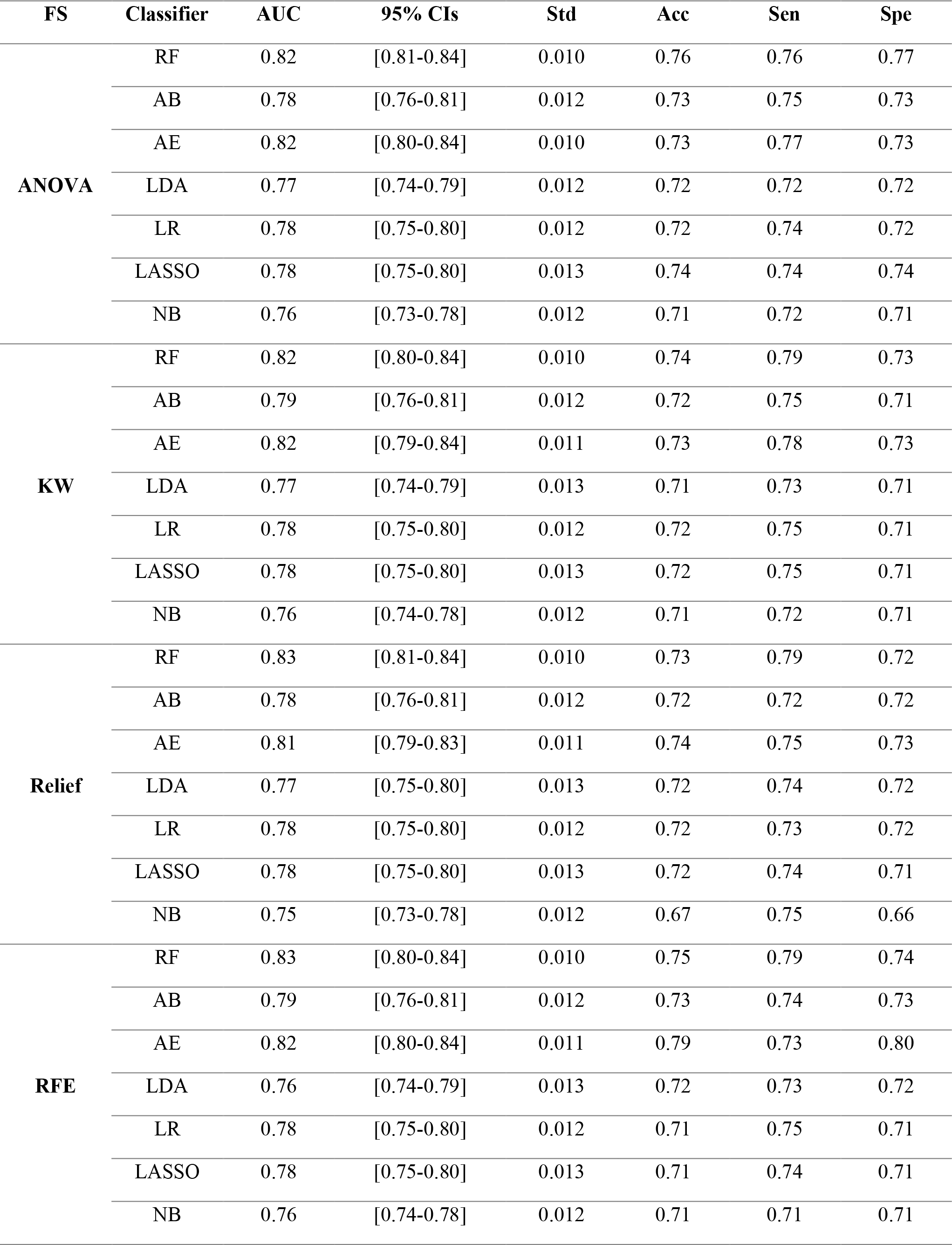
Classification performance indices of different feature selector and classifiers in Strategy 6.

**Supplemental Table 8.** Classification performance indices of different feature selector and classifiers in Strategy 7.

| **FS** | **Classifier** | **AUC** | **95% CIs** | **Std** | **Acc** | **Sen** | **Spe** |
| --- | --- | --- | --- | --- | --- | --- | --- |
| **ANOVA** | RF | 0.78 | [0.76-0.80] | 0.011 | 0.73 | 0.71 | 0.73 |
|  | AB | 0.75 | [0.72-0.77] | 0.012 | 0.67 | 0.73 | 0.66 |
|  | AE | 0.78 | [0.76-0.80] | 0.011 | 0.73 | 0.70 | 0.73 |
|  | LDA | 0.76 | [0.73-0.78] | 0.012 | 0.72 | 0.70 | 0.72 |
|  | LR | 0.76 | [0.74-0.78] | 0.012 | 0.71 | 0.71 | 0.71 |
|  | LASSO | 0.76 | [0.74-0.79] | 0.012 | 0.71 | 0.72 | 0.71 |
|  | NB | 0.74 | [0.71-0.76] | 0.013 | 0.70 | 0.70 | 0.70 |
| **KW** | RF | 0.78 | [0.76-0.80] | 0.011 | 0.72 | 0.72 | 0.72 |
|  | AB | 0.76 | [0.73-0.78] | 0.012 | 0.71 | 0.70 | 0.71 |
|  | AE | 0.77 | [0.75-0.80] | 0.012 | 0.74 | 0.70 | 0.74 |
|  | LDA | 0.76 | [0.74-0.79] | 0.012 | 0.71 | 0.70 | 0.71 |
|  | LR | 0.76 | [0.74-0.78] | 0.012 | 0.75 | 0.67 | 0.76 |
|  | LASSO | 0.77 | [0.74-0.79] | 0.012 | 0.71 | 0.71 | 0.71 |
|  | NB | 0.74 | [0.72-0.77] | 0.013 | 0.74 | 0.65 | 0.75 |
| **Relief** | RF | 0.78 | [0.76-0.80] | 0.011 | 0.72 | 0.73 | 0.72 |
|  | AB | 0.74 | [0.72-0.76] | 0.012 | 0.68 | 0.72 | 0.67 |
|  | AE | 0.76 | [0.74-0.79] | 0.012 | 0.71 | 0.71 | 0.71 |
|  | LDA | 0.75 | [0.73-0.78] | 0.012 | 0.70 | 0.69 | 0.71 |
|  | LR | 0.76 | [0.74-0.78] | 0.012 | 0.73 | 0.67 | 0.74 |
|  | LASSO | 0.77 | [0.74-0.79] | 0.012 | 0.68 | 0.74 | 0.68 |
|  | NB | 0.73 | [0.70-0.75] | 0.013 | 0.72 | 0.67 | 0.72 |
| **RFE** | RF | 0.79 | [0.76-0.81] | 0.011 | 0.71 | 0.73 | 0.71 |
|  | AB | 0.75 | [0.72-0.77] | 0.012 | 0.68 | 0.73 | 0.67 |
|  | AE | 0.77 | [0.75-0.79] | 0.012 | 0.71 | 0.73 | 0.70 |
|  | LDA | 0.76 | [0.73-0.78] | 0.012 | 0.71 | 0.71 | 0.70 |
|  | LR | 0.76 | [0.74-0.79] | 0.012 | 0.72 | 0.70 | 0.73 |
|  | LASSO | 0.77 | [0.74-0.79] | 0.012 | 0.71 | 0.71 | 0.71 |
|  | NB | 0.74 | [0.72-0.77] | 0.013 | 0.72 | 0.67 | 0.73 |

**Supplemental Table 9.** Classification performance indices of different feature selector and classifiers in Strategy 8.

| FS | Classifier | AUC | Acc | Sen | Spe |
| --- | --- | --- | --- | --- | --- |
| **ANOVA** | RF | $0.67 \pm 0.072$ | $0.61 \pm 0.085$ | $0.67 \pm 0.058$ | $0.6 \pm 0.096$ |
| | AB | $0.69 \pm 0.064$ | $0.65 \pm 0.059$ | $0.66 \pm 0.059$ | $0.64 \pm 0.065$ |
| | AE | $0.72 \pm 0.073$ | $0.66 \pm 0.068$ | $0.7 \pm 0.069$ | $0.65 \pm 0.077$ |
| | LDA | $0.71 \pm 0.062$ | $0.66 \pm 0.058$ | $0.69 \pm 0.037$ | $0.66 \pm 0.063$ |
| | LR | $0.74 \pm 0.051$ | $0.68 \pm 0.047$ | $0.71 \pm 0.039$ | $0.67 \pm 0.051$ |
| | LASSO | $0.74 \pm 0.051$ | $0.68 \pm 0.042$ | $0.71 \pm 0.041$ | $0.68 \pm 0.045$ |
| | NB | $0.74 \pm 0.034$ | $0.69 \pm 0.03$ | $0.71 \pm 0.026$ | $0.69 \pm 0.033$ |
| **KW** | RF | $0.67 \pm 0.08$ | $0.6 \pm 0.09$ | $0.67 \pm 0.074$ | $0.59 \pm 0.1$ |
| | AB | $0.68 \pm 0.079$ | $0.61 \pm 0.064$ | $0.69 \pm 0.063$ | $0.6 \pm 0.068$ |
| | AE | $0.72 \pm 0.07$ | $0.63 \pm 0.097$ | $0.73 \pm 0.067$ | $0.62 \pm 0.11$ |
| | LDA | $0.7 \pm 0.074$ | $0.65 \pm 0.071$ | $0.68 \pm 0.05$ | $0.64 \pm 0.079$ |
| | LR | $0.72 \pm 0.066$ | $0.65 \pm 0.051$ | $0.71 \pm 0.037$ | $0.65 \pm 0.056$ |
| | LASSO | $0.74 \pm 0.053$ | $0.67 \pm 0.043$ | $0.71 \pm 0.039$ | $0.67 \pm 0.046$ |
| | NB | $0.74 \pm 0.059$ | $0.69 \pm 0.049$ | $0.7 \pm 0.043$ | $0.69 \pm 0.052$ |
| **Relief** | RF | $0.66 \pm 0.066$ | $0.59 \pm 0.069$ | $0.67 \pm 0.057$ | $0.58 \pm 0.077$ |
| | AB | $0.67 \pm 0.062$ | $0.58 \pm 0.073$ | $0.7 \pm 0.079$ | $0.57 \pm 0.084$ |
| | AE | $0.71 \pm 0.057$ | $0.62 \pm 0.076$ | $0.71 \pm 0.039$ | $0.61 \pm 0.088$ |
| | LDA | $0.69 \pm 0.061$ | $0.63 \pm 0.05$ | $0.67 \pm 0.051$ | $0.63 \pm 0.052$ |
| | LR | $0.71 \pm 0.063$ | $0.63 \pm 0.068$ | $0.71 \pm 0.043$ | $0.62 \pm 0.077$ |
| | LASSO | $0.7 \pm 0.069$ | $0.62 \pm 0.064$ | $0.71 \pm 0.053$ | $0.61 \pm 0.07$ |
| | NB | $0.64 \pm 0.087$ | $0.63 \pm 0.1$ | $0.61 \pm 0.12$ | $0.63 \pm 0.12$ |
| **RFE** | RF | $0.67 \pm 0.071$ | $0.6 \pm 0.064$ | $0.67 \pm 0.053$ | $0.6 \pm 0.069$ |
| | AB | $0.68 \pm 0.049$ | $0.6 \pm 0.059$ | $0.71 \pm 0.058$ | $0.58 \pm 0.07$ |
| | AE | $0.72 \pm 0.066$ | $0.63 \pm 0.079$ | $0.72 \pm 0.029$ | $0.62 \pm 0.089$ |
| | LDA | $0.68 \pm 0.058$ | $0.61 \pm 0.06$ | $0.7 \pm 0.047$ | $0.6 \pm 0.069$ |
| | LR | $0.7 \pm 0.062$ | $0.63 \pm 0.059$ | $0.7 \pm 0.056$ | $0.62 \pm 0.067$ |
| | LASSO | $0.7 \pm 0.062$ | $0.62 \pm 0.056$ | $0.7 \pm 0.052$ | $0.61 \pm 0.063$ |
| | NB | $0.67 \pm 0.064$ | $0.59 \pm 0.099$ | $0.72 \pm 0.081$ | $0.57 \pm 0.12$ |

**Supplemental Table 10.** Classification performance indices of different feature selector and classifiers in Strategy 9.

| FS | Classifier | AUC | Acc | Sen | Spe |
| --- | --- | --- | --- | --- | --- |
| **ANOVA** | RF | $0.79 \pm 0.08$ | $0.74 \pm 0.079$ | $0.77 \pm 0.086$ | $0.73 \pm 0.078$ |
| | AB | $0.79 \pm 0.08$ | $0.75 \pm 0.073$ | $0.77 \pm 0.099$ | $0.74 \pm 0.074$ |
| | AE | $0.80 \pm 0.08$ | $0.77 \pm 0.075$ | $0.78 \pm 0.095$ | $0.76 \pm 0.078$ |
| | LDA | $0.79 \pm 0.08$ | $0.76 \pm 0.072$ | $0.76 \pm 0.1$ | $0.75 \pm 0.075$ |
| | LR | $0.80 \pm 0.08$ | $0.76 \pm 0.075$ | $0.77 \pm 0.11$ | $0.76 \pm 0.075$ |
| | LASSO | $0.80 \pm 0.08$ | $0.75 \pm 0.076$ | $0.77 \pm 0.099$ | $0.75 \pm 0.076$ |
| | NB | $0.78 \pm 0.08$ | $0.75 \pm 0.068$ | $0.74 \pm 0.11$ | $0.76 \pm 0.07$ |
| **KW** | RF | $0.79 \pm 0.08$ | $0.72 \pm 0.093$ | $0.78 \pm 0.11$ | $0.71 \pm 0.098$ |
| | AB | $0.79 \pm 0.086$ | $0.74 \pm 0.088$ | $0.76 \pm 0.16$ | $0.74 \pm 0.11$ |
| | AE | $0.81 \pm 0.083$ | $0.73 \pm 0.095$ | $0.81 \pm 0.13$ | $0.73 \pm 0.1$ |
| | LDA | $0.80 \pm 0.084$ | $0.74 \pm 0.092$ | $0.77 \pm 0.13$ | $0.74 \pm 0.11$ |
| | LR | $0.80 \pm 0.084$ | $0.76 \pm 0.085$ | $0.76 \pm 0.14$ | $0.76 \pm 0.097$ |
| | LASSO | $0.80 \pm 0.084$ | $0.75 \pm 0.095$ | $0.77 \pm 0.14$ | $0.74 \pm 0.11$ |
| | NB | $0.78 \pm 0.083$ | $0.76 \pm 0.07$ | $0.75 \pm 0.12$ | $0.76 \pm 0.074$ |
| **Relief** | RF | $0.79 \pm 0.086$ | $0.72 \pm 0.098$ | $0.79 \pm 0.12$ | $0.71 \pm 0.1$ |
| | AB | $0.79 \pm 0.08$ | $0.71 \pm 0.092$ | $0.81 \pm 0.08$ | $0.69 \pm 0.1$ |
| | AE | $0.8 \pm 0.087$ | $0.73 \pm 0.093$ | $0.8 \pm 0.11$ | $0.72 \pm 0.1$ |
| | LDA | $0.8 \pm 0.084$ | $0.73 \pm 0.092$ | $0.8 \pm 0.11$ | $0.72 \pm 0.1$ |
| | LR | $0.79 \pm 0.077$ | $0.74 \pm 0.083$ | $0.78 \pm 0.11$ | $0.74 \pm 0.09$ |
| | LASSO | $0.8 \pm 0.086$ | $0.75 \pm 0.095$ | $0.77 \pm 0.13$ | $0.74 \pm 0.11$ |
| | NB | $0.75 \pm 0.075$ | $0.73 \pm 0.065$ | $0.71 \pm 0.11$ | $0.74 \pm 0.074$ |
| **RFE** | RF | $0.79 \pm 0.082$ | $0.71 \pm 0.089$ | $0.8 \pm 0.099$ | $0.7 \pm 0.096$ |
| | AB | $0.79 \pm 0.077$ | $0.72 \pm 0.073$ | $0.8 \pm 0.081$ | $0.7 \pm 0.08$ |
| | AE | $0.81 \pm 0.082$ | $0.74 \pm 0.087$ | $0.8 \pm 0.13$ | $0.73 \pm 0.099$ |
| | LDA | $0.79 \pm 0.089$ | $0.74 \pm 0.093$ | $0.78 \pm 0.11$ | $0.73 \pm 0.11$ |
| | LR | $0.8 \pm 0.084$ | $0.75 \pm 0.09$ | $0.78 \pm 0.1$ | $0.75 \pm 0.092$ |
| | LASSO | $0.8 \pm 0.082$ | $0.75 \pm 0.073$ | $0.76 \pm 0.17$ | $0.76 \pm 0.09$ |
| | NB | $0.76 \pm 0.069$ | $0.74 \pm 0.048$ | $0.73 \pm 0.092$ | $0.74 \pm 0.052$ |

**Supplemental Table 11.** Classification performance indices of different feature selector and classifiers in Strategy 10.

| FS | Classifier | AUC | Acc | Sen | Spe |
| --- | --- | --- | --- | --- | --- |
| **ANOVA** | RF | $0.82 \pm 0.095$ | $0.78 \pm 0.11$ | $0.82 \pm 0.097$ | $0.77 \pm 0.11$ |
| | AB | $0.8 \pm 0.13$ | $0.77 \pm 0.11$ | $0.81 \pm 0.11$ | $0.76 \pm 0.12$ |
| | AE | $0.79 \pm 0.11$ | $0.74 \pm 0.11$ | $0.83 \pm 0.11$ | $0.73 \pm 0.11$ |
| | LDA | $0.82 \pm 0.1$ | $0.77 \pm 0.094$ | $0.83 \pm 0.11$ | $0.77 \pm 0.093$ |
| | LR | $0.82 \pm 0.1$ | $0.78 \pm 0.093$ | $0.84 \pm 0.12$ | $0.77 \pm 0.094$ |
| | LASSO | $0.81 \pm 0.099$ | $0.78 \pm 0.085$ | $0.82 \pm 0.12$ | $0.78 \pm 0.085$ |
| | NB | $0.79 \pm 0.12$ | $0.76 \pm 0.12$ | $0.83 \pm 0.12$ | $0.75 \pm 0.13$ |
| **KW** | RF | $0.82 \pm 0.09$ | $0.76 \pm 0.11$ | $0.83 \pm 0.11$ | $0.75 \pm 0.12$ |
| | AB | $0.8 \pm 0.14$ | $0.74 \pm 0.15$ | $0.84 \pm 0.14$ | $0.72 \pm 0.16$ |
| | AE | $0.79 \pm 0.11$ | $0.73 \pm 0.14$ | $0.83 \pm 0.13$ | $0.72 \pm 0.16$ |
| | LDA | $0.81 \pm 0.1$ | $0.75 \pm 0.1$ | $0.84 \pm 0.14$ | $0.74 \pm 0.11$ |
| | LR | $0.81 \pm 0.097$ | $0.75 \pm 0.11$ | $0.85 \pm 0.11$ | $0.75 \pm 0.12$ |
| | LASSO | $0.81 \pm 0.095$ | $0.76 \pm 0.1$ | $0.85 \pm 0.11$ | $0.75 \pm 0.11$ |
| | NB | $0.79 \pm 0.11$ | $0.75 \pm 0.12$ | $0.83 \pm 0.15$ | $0.74 \pm 0.13$ |
| **Relief** | RF | $0.83 \pm 0.1$ | $0.75 \pm 0.12$ | $0.85 \pm 0.11$ | $0.74 \pm 0.13$ |
| | AB | $0.77 \pm 0.14$ | $0.71 \pm 0.15$ | $0.84 \pm 0.13$ | $0.69 \pm 0.15$ |
| | AE | $0.79 \pm 0.11$ | $0.74 \pm 0.11$ | $0.81 \pm 0.13$ | $0.72 \pm 0.12$ |
| | LDA | $0.81 \pm 0.11$ | $0.74 \pm 0.1$ | $0.86 \pm 0.11$ | $0.72 \pm 0.11$ |
| | LR | $0.81 \pm 0.11$ | $0.73 \pm 0.13$ | $0.86 \pm 0.13$ | $0.71 \pm 0.14$ |
| | LASSO | $0.8 \pm 0.11$ | $0.73 \pm 0.12$ | $0.87 \pm 0.12$ | $0.71 \pm 0.13$ |
| | NB | $0.79 \pm 0.12$ | $0.74 \pm 0.13$ | $0.83 \pm 0.12$ | $0.72 \pm 0.14$ |
| **RFE** | RF | $0.82 \pm 0.099$ | $0.72 \pm 0.13$ | $0.87 \pm 0.092$ | $0.7 \pm 0.13$ |
| | AB | $0.8 \pm 0.14$ | $0.76 \pm 0.13$ | $0.84 \pm 0.1$ | $0.74 \pm 0.13$ |
| | AE | $0.8 \pm 0.1$ | $0.74 \pm 0.1$ | $0.84 \pm 0.13$ | $0.72 \pm 0.11$ |
| | LDA | $0.81 \pm 0.11$ | $0.73 \pm 0.11$ | $0.87 \pm 0.1$ | $0.71 \pm 0.12$ |
| | LR | $0.8 \pm 0.12$ | $0.73 \pm 0.14$ | $0.85 \pm 0.15$ | $0.71 \pm 0.15$ |
| | LASSO | $0.79 \pm 0.11$ | $0.73 \pm 0.13$ | $0.83 \pm 0.16$ | $0.72 \pm 0.14$ |
| | NB | $0.79 \pm 0.12$ | $0.75 \pm 0.13$ | $0.83 \pm 0.13$ | $0.74 \pm 0.14$ |

**Supplemental Table 12.**
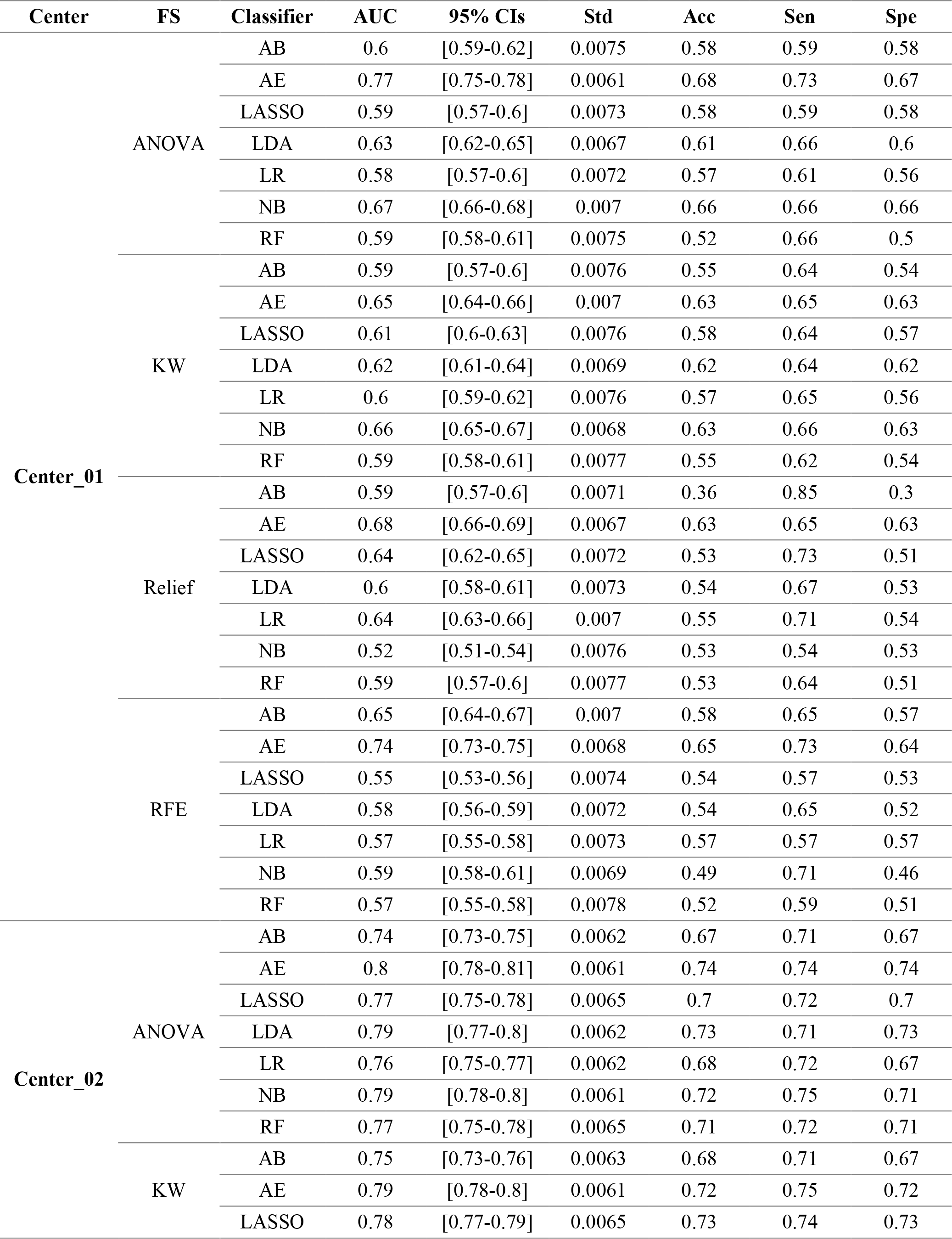
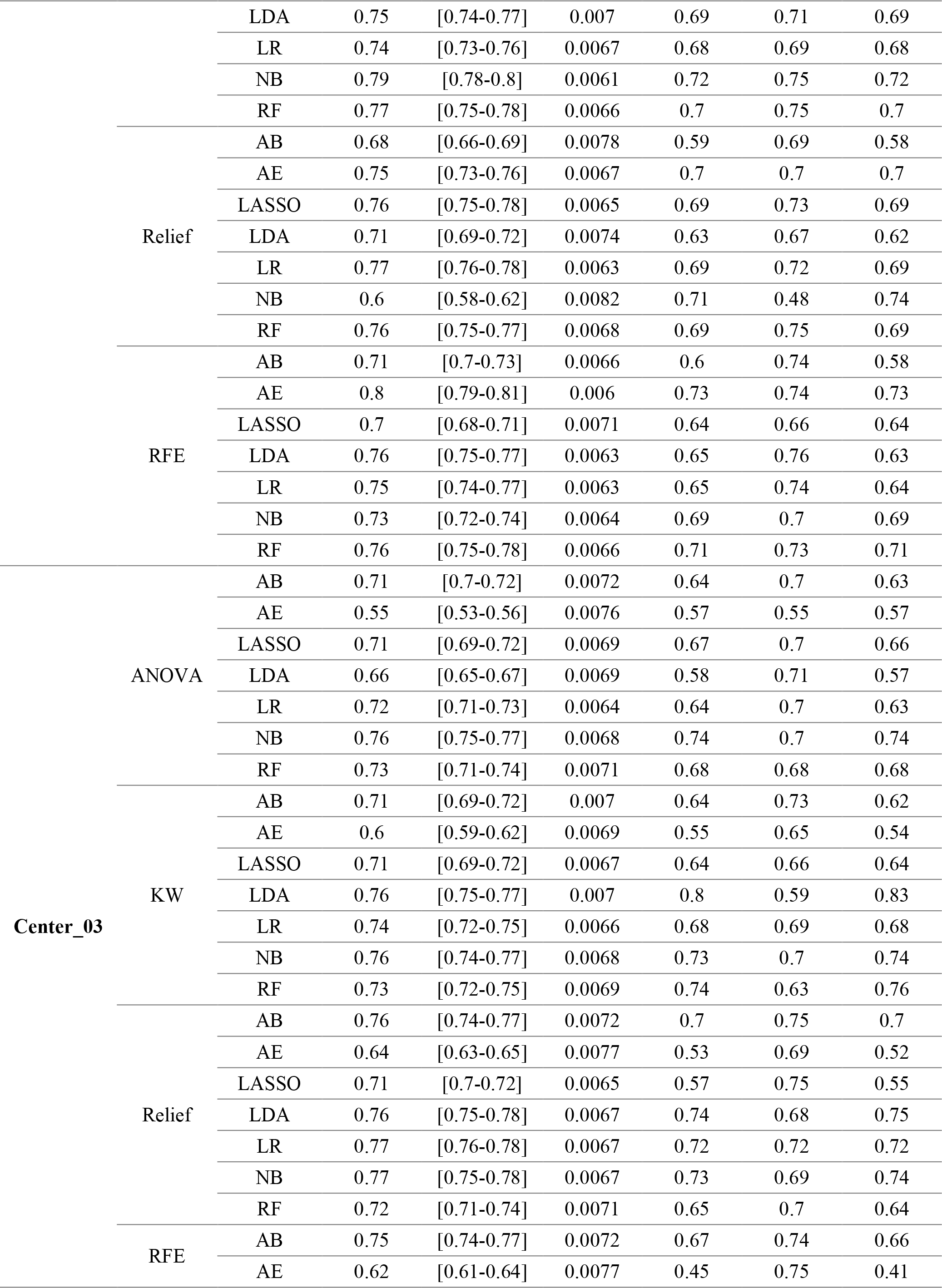
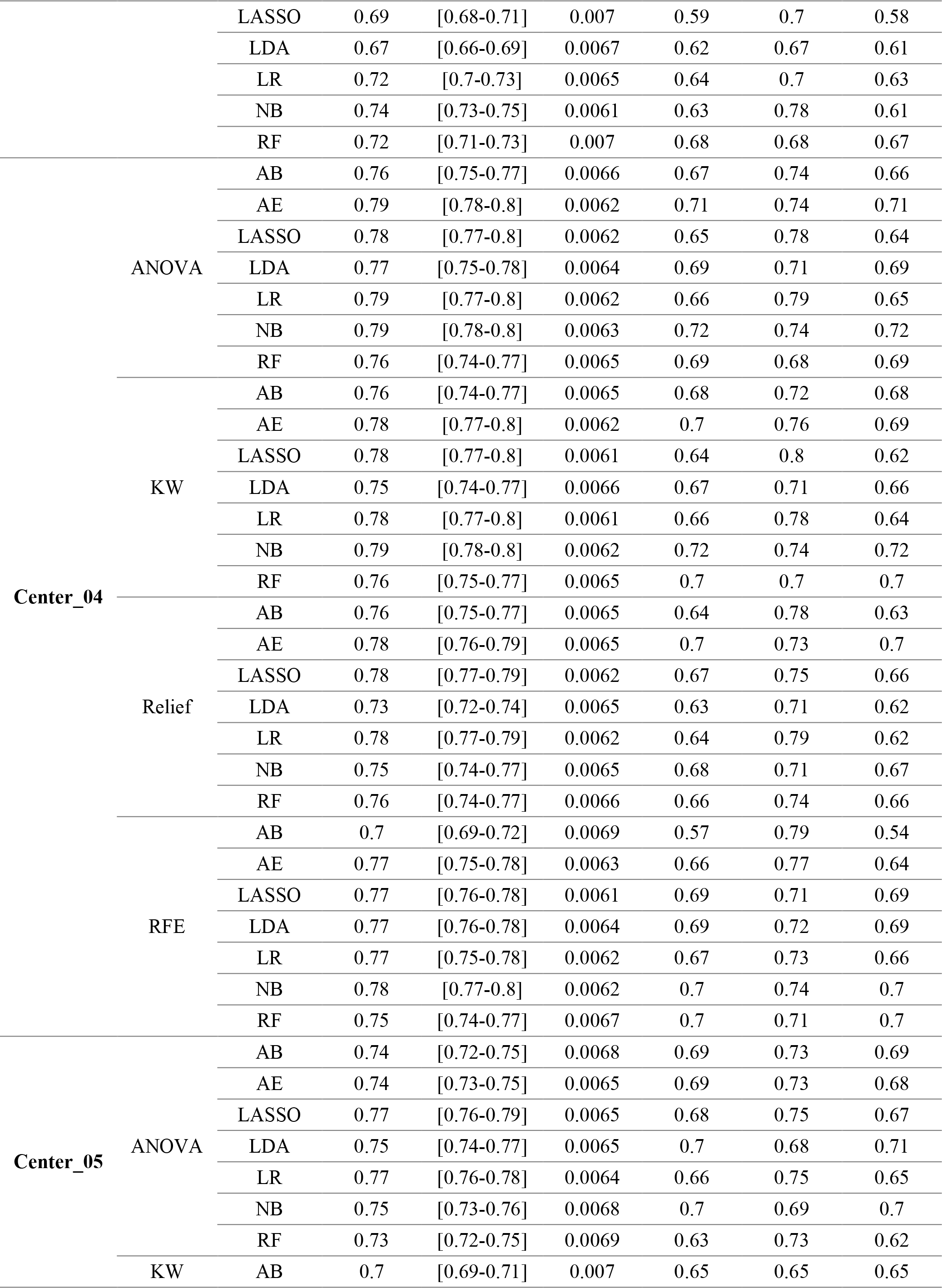
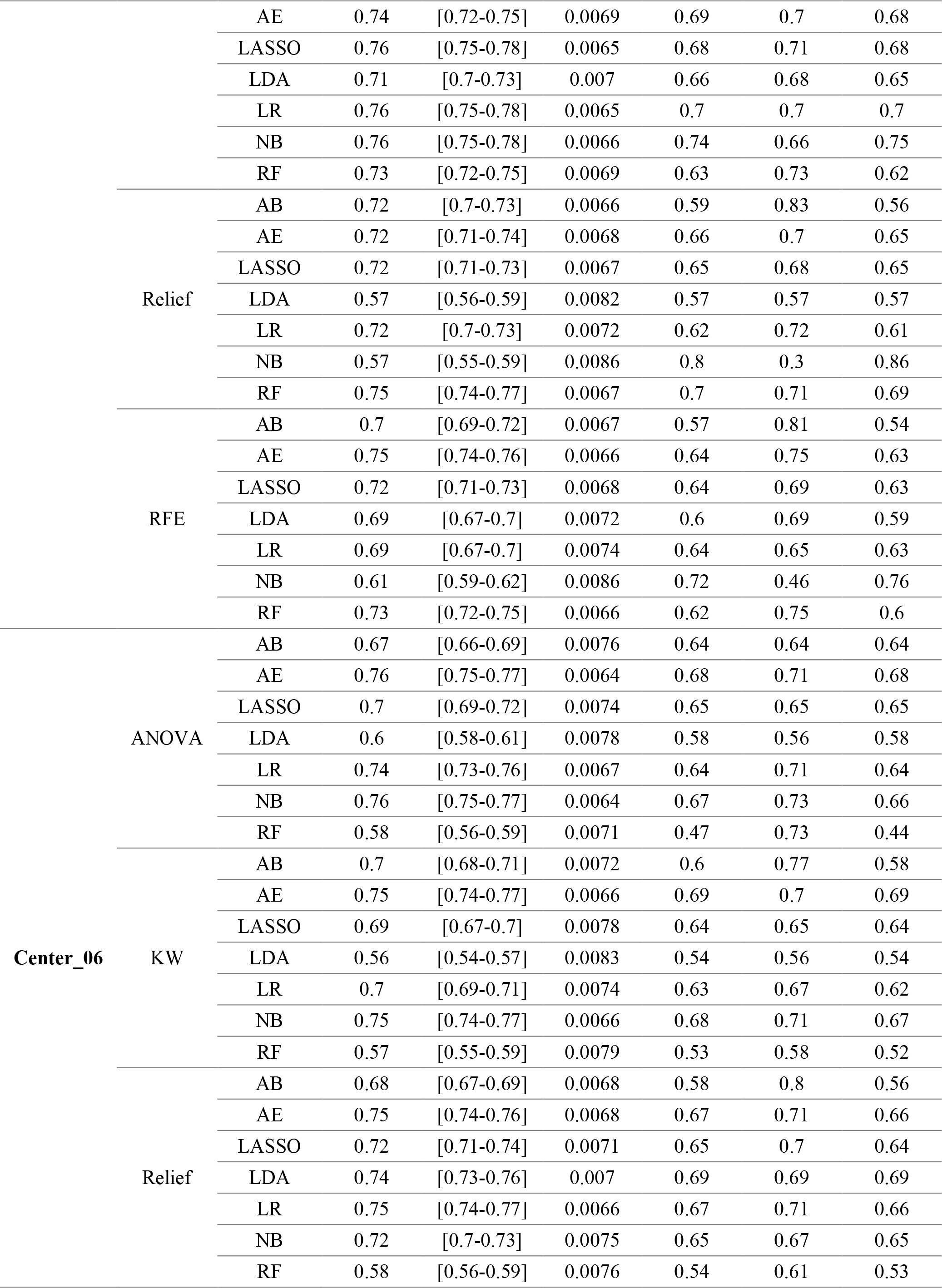
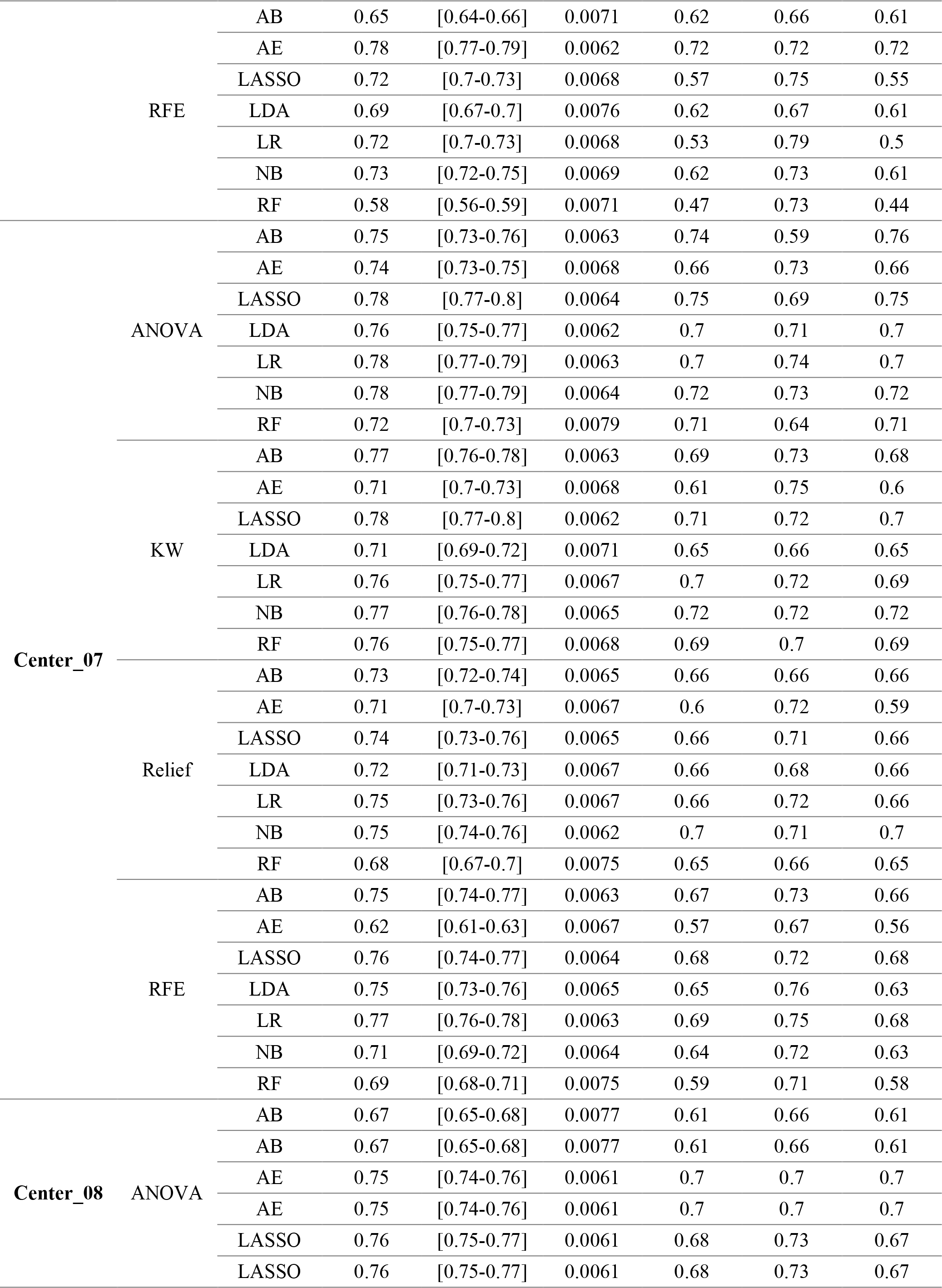
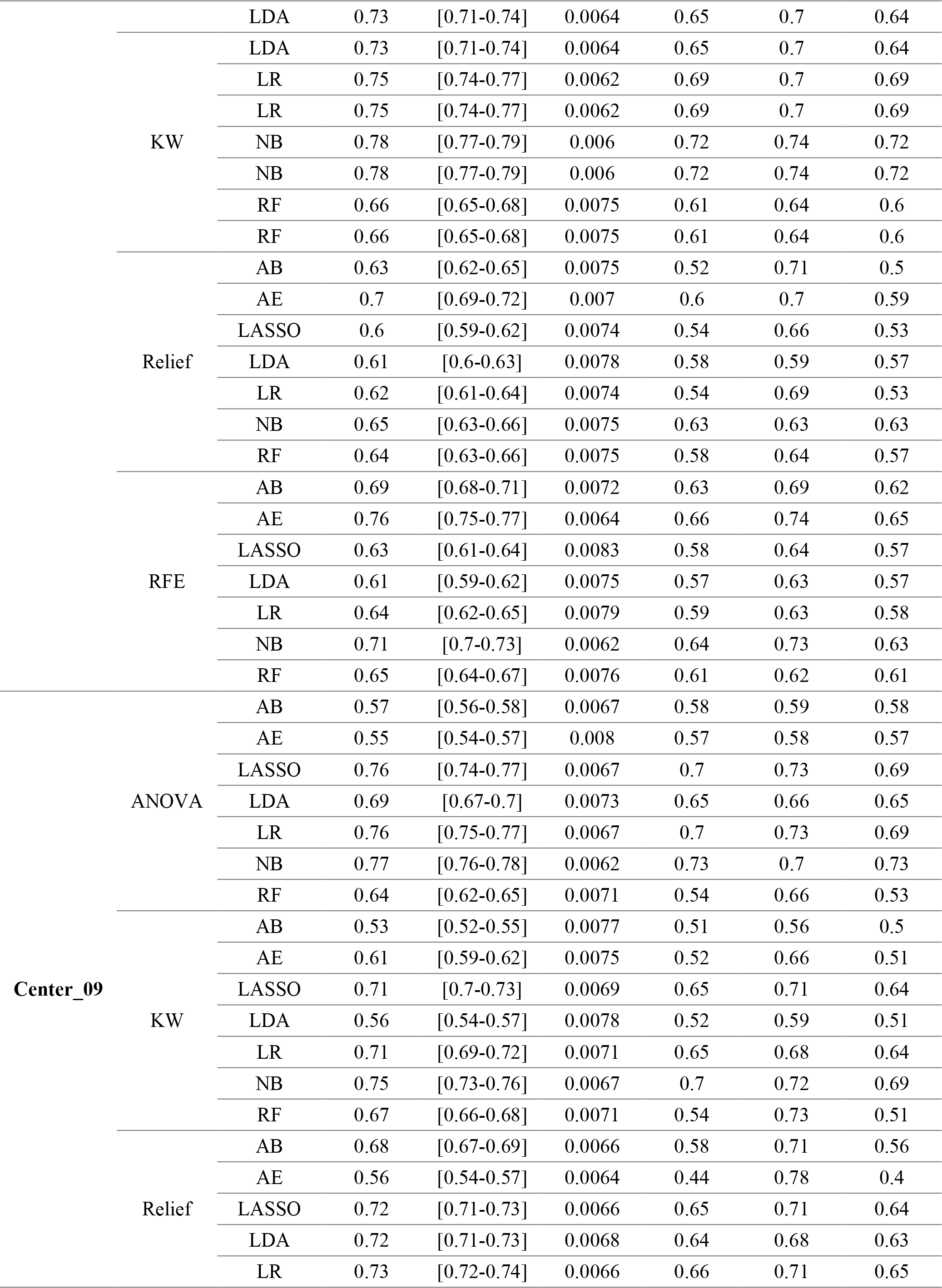
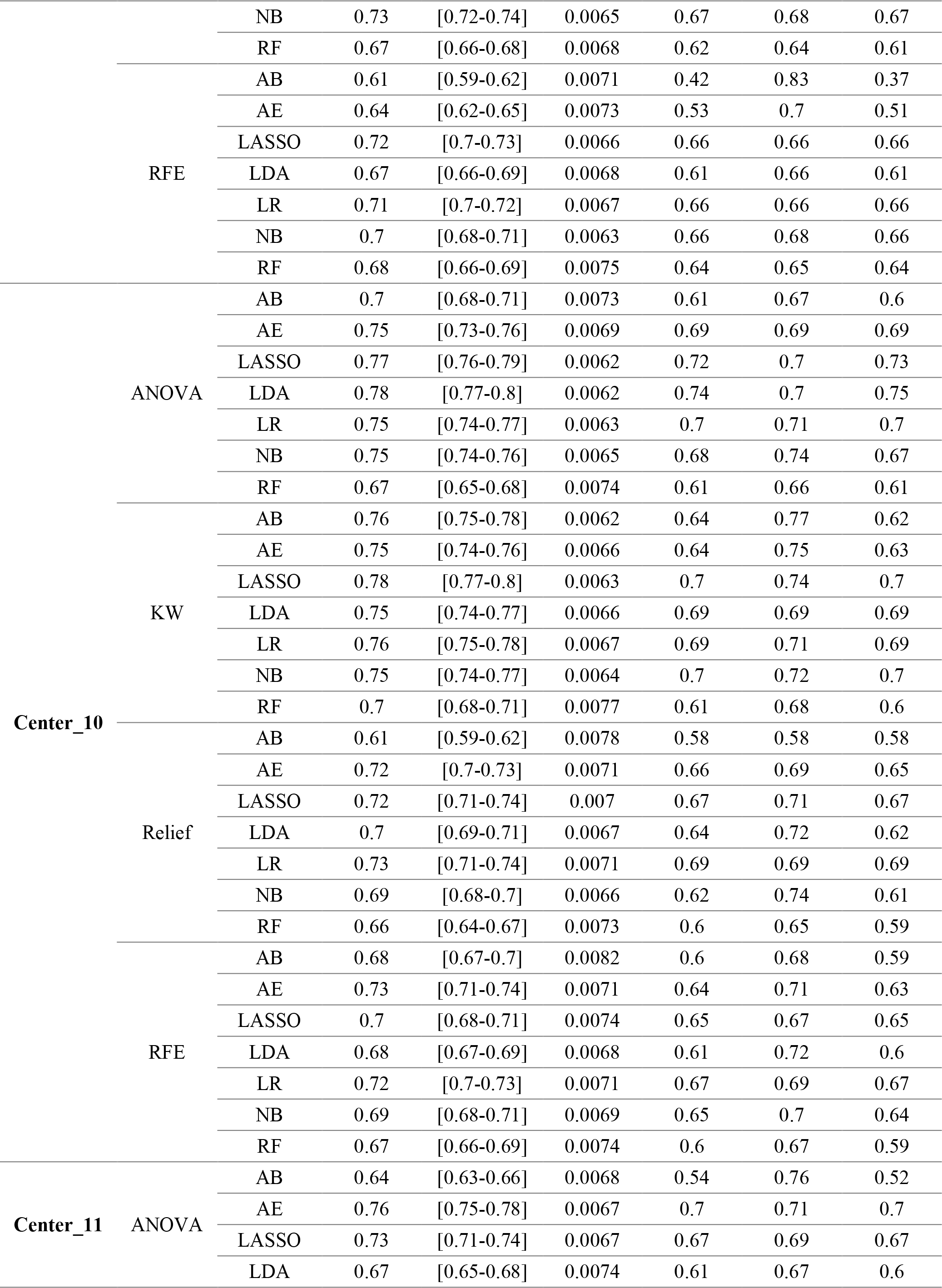
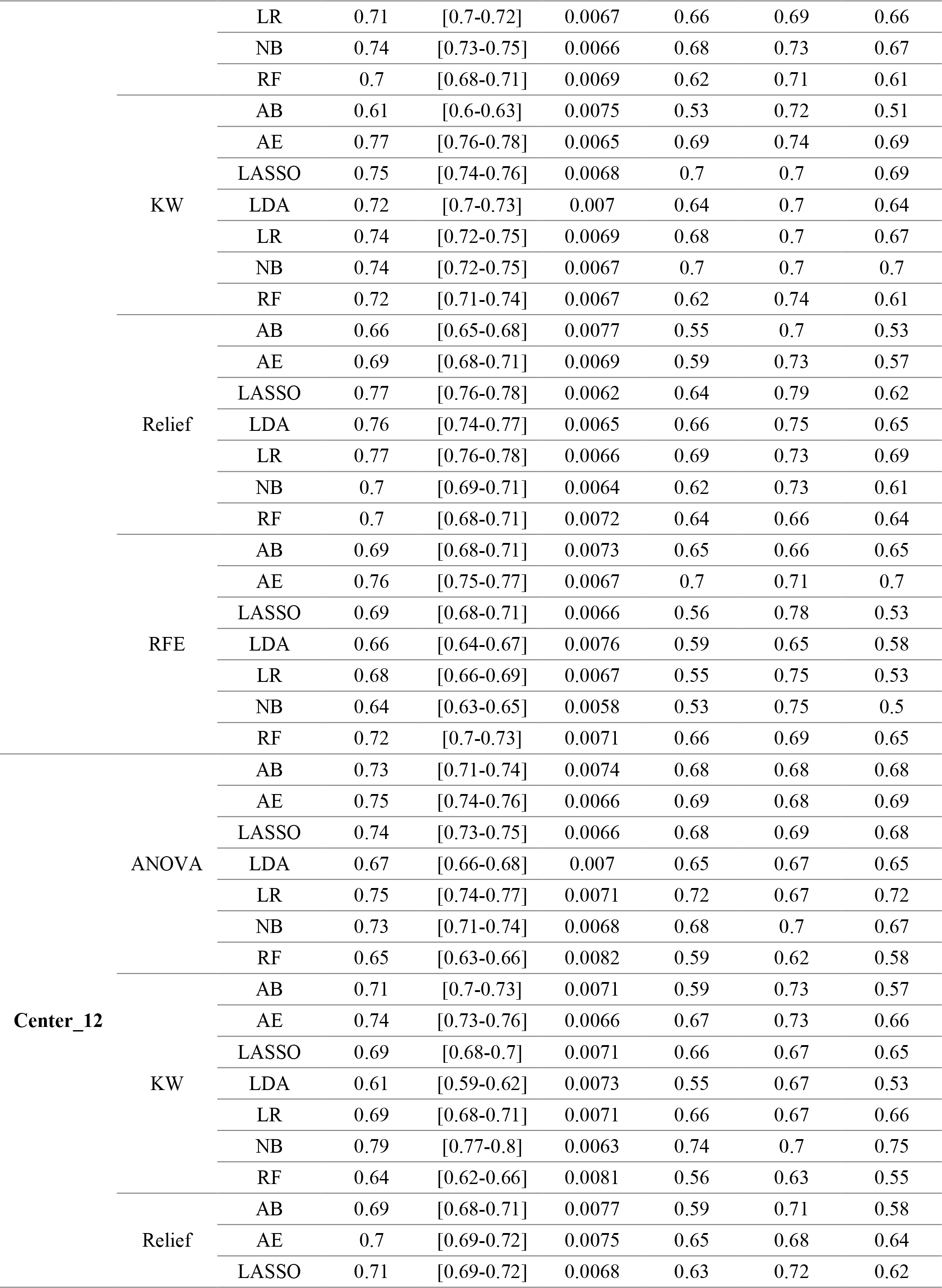
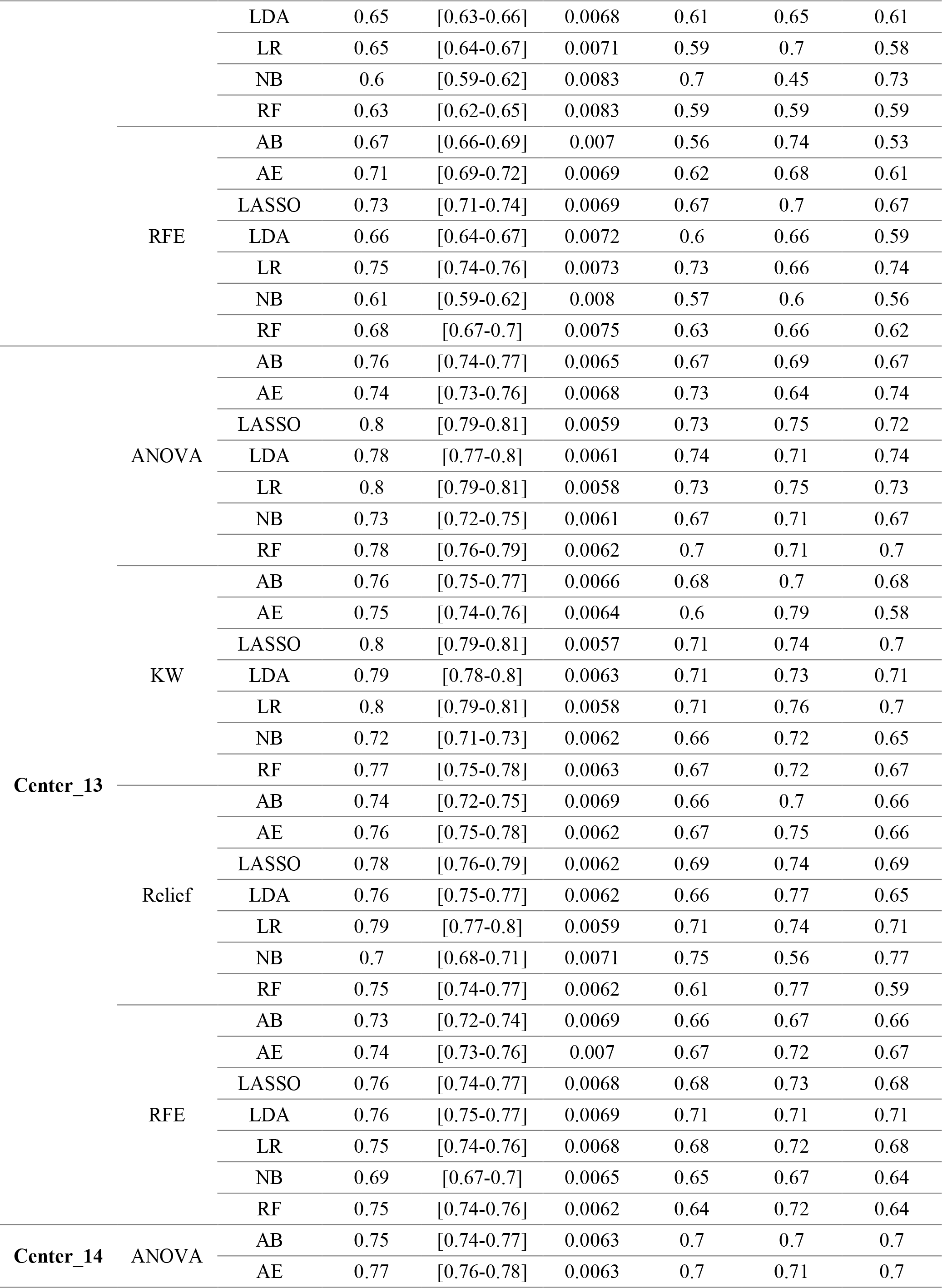
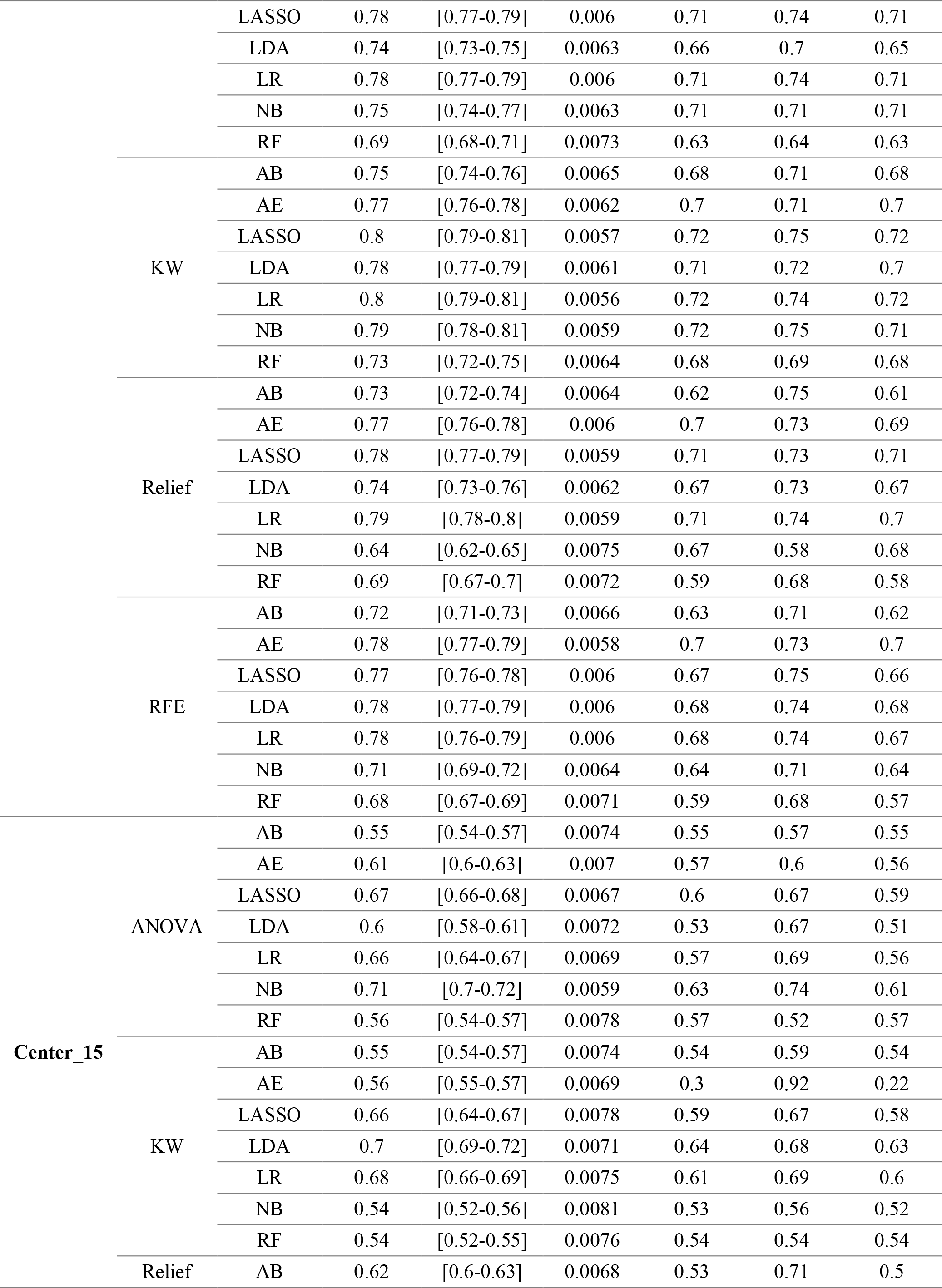
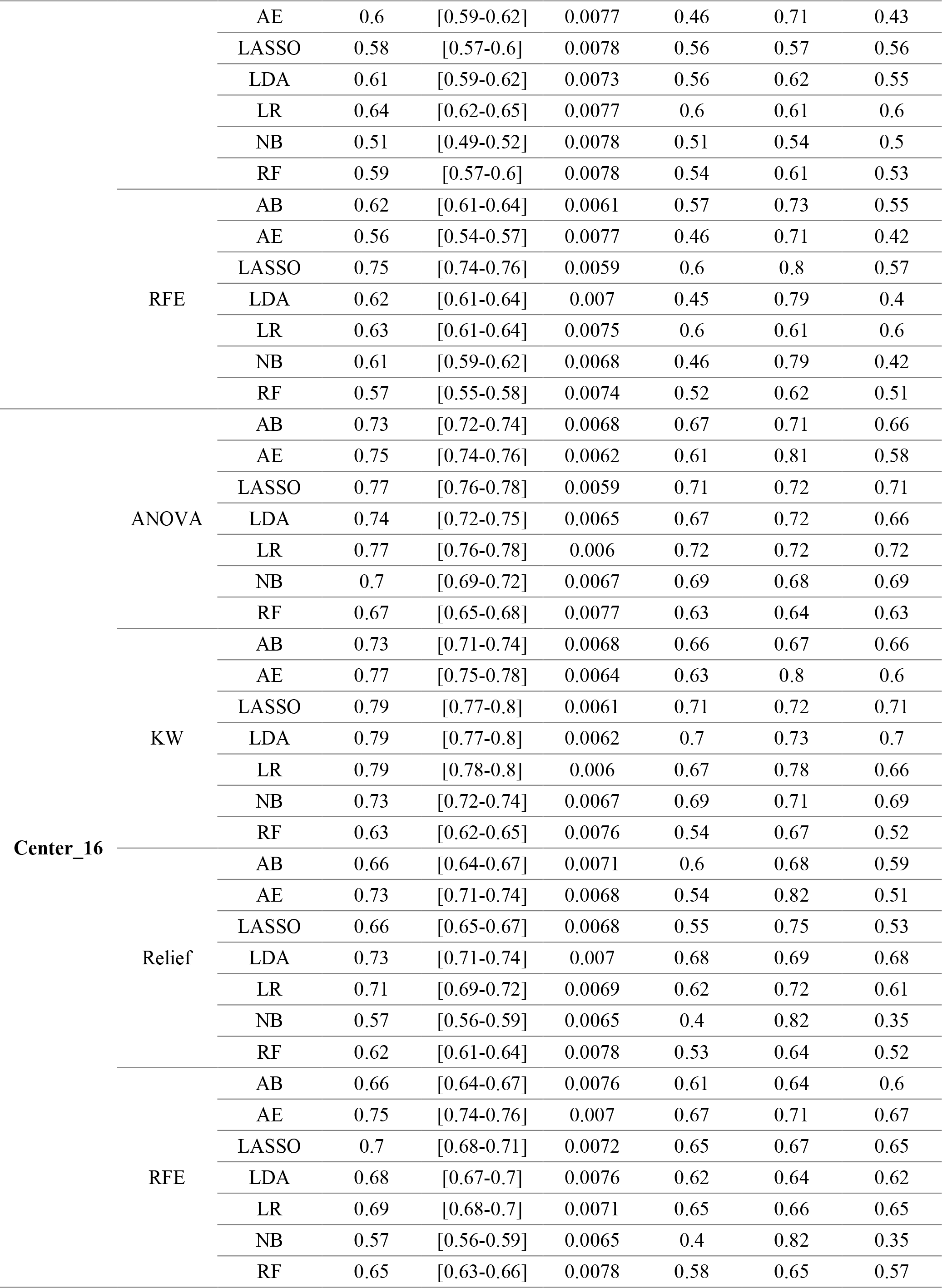
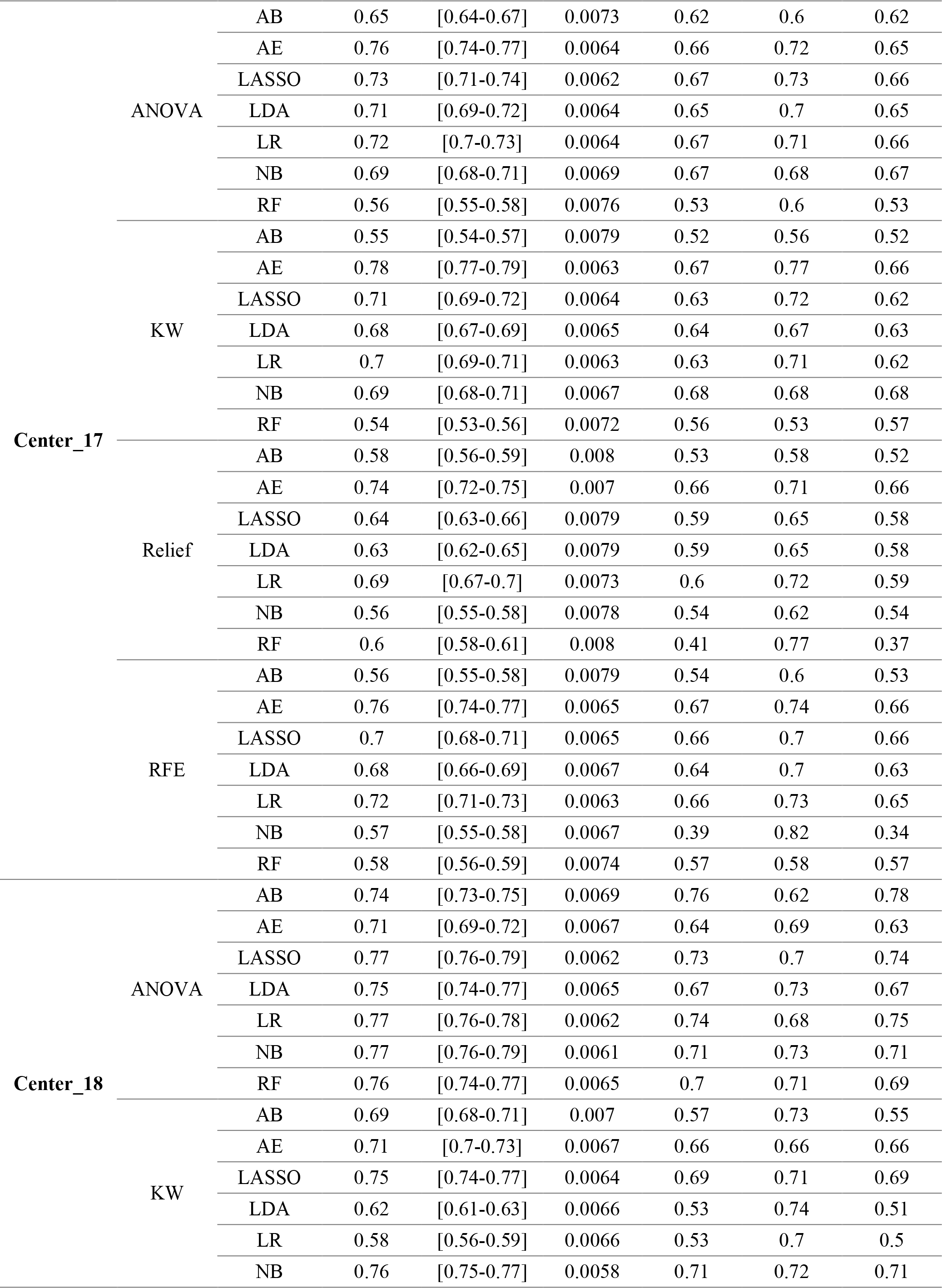
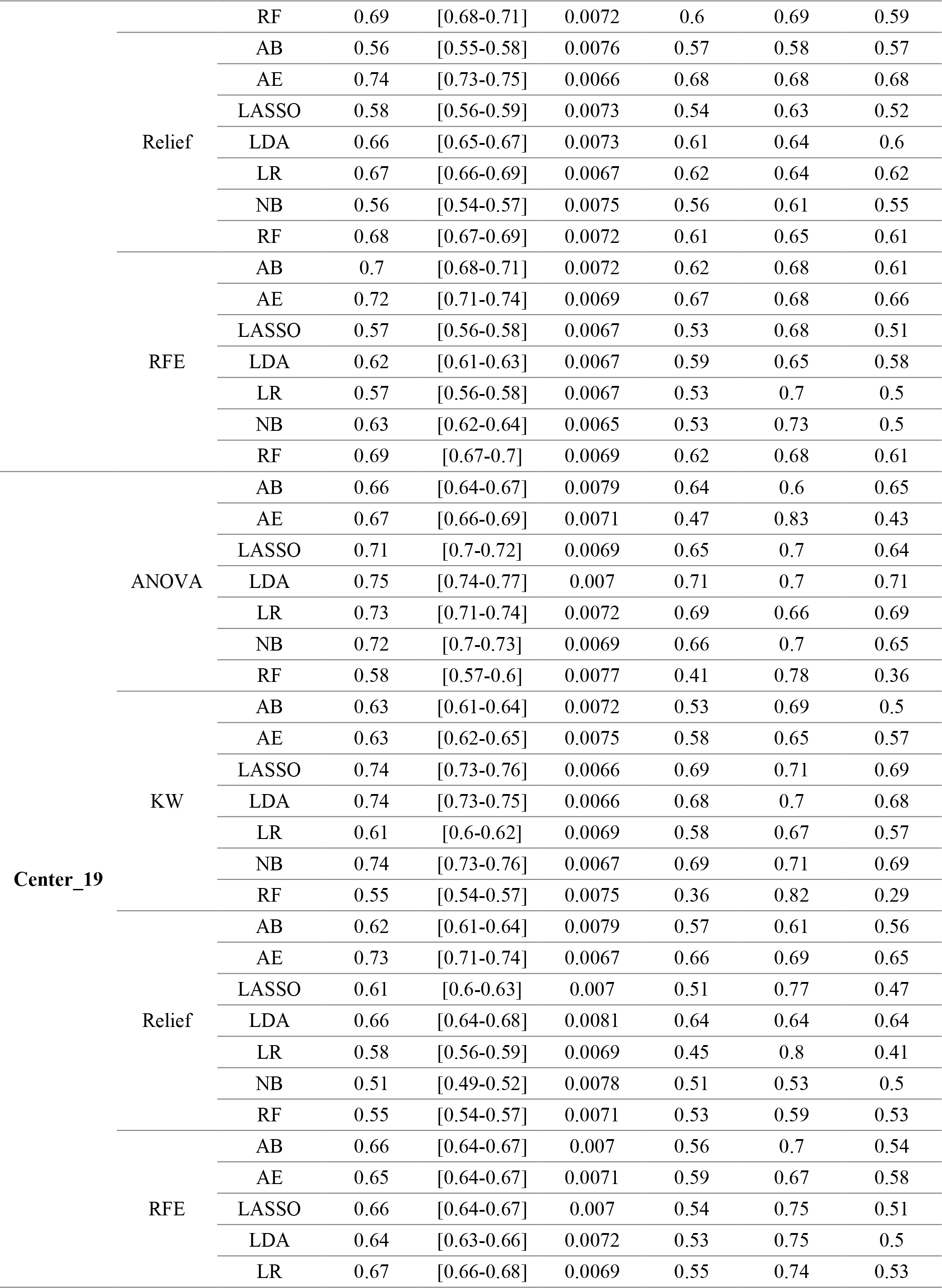
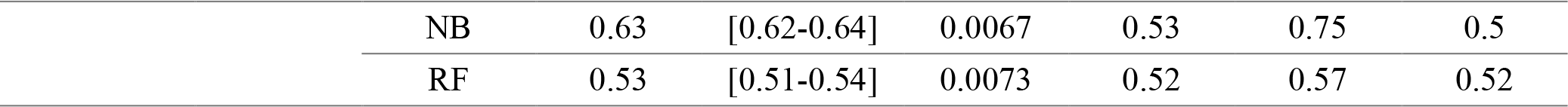
Classification performance indices of different feature selector and classifiers in Strategy 8.

**Supplemental Table 13.**
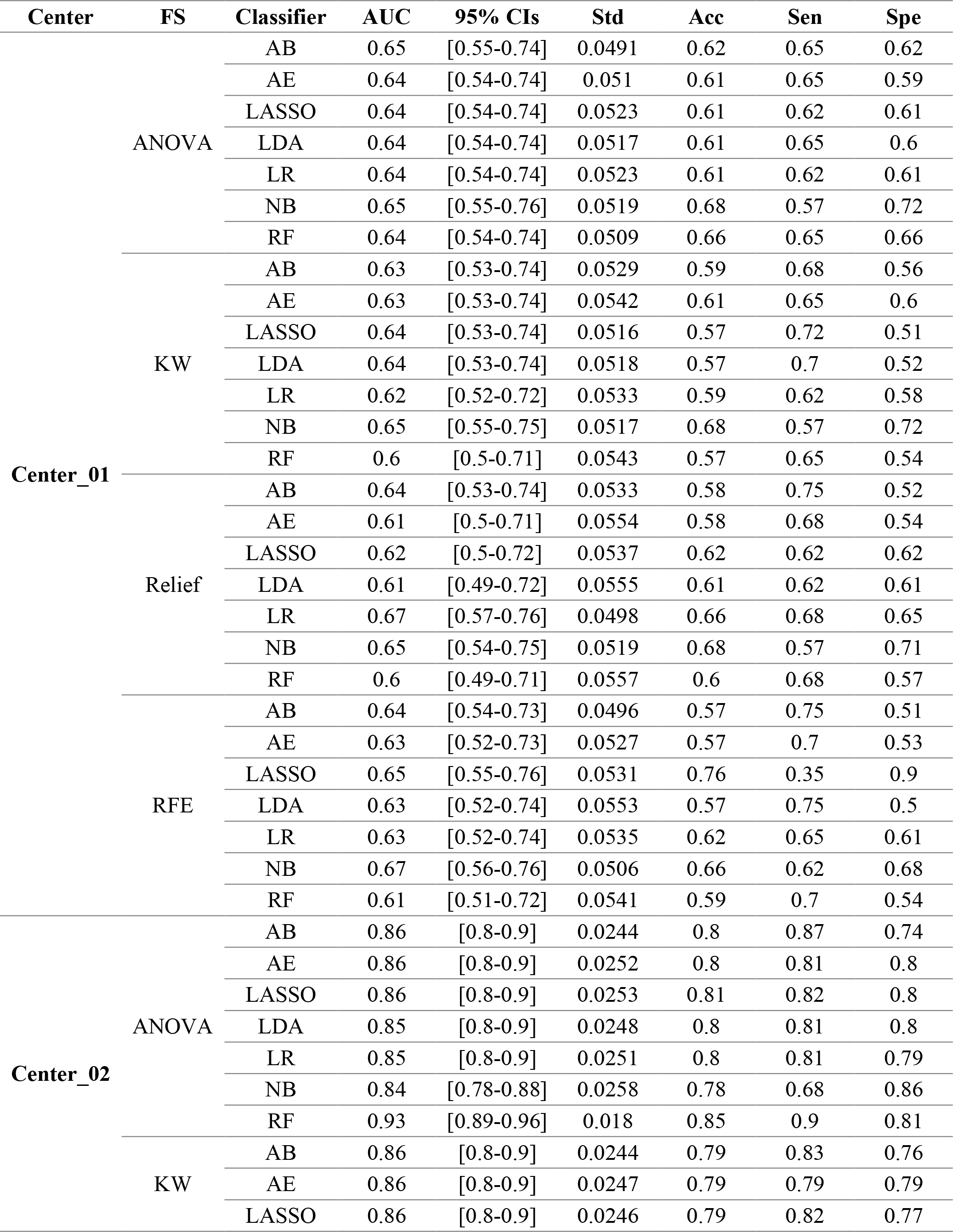
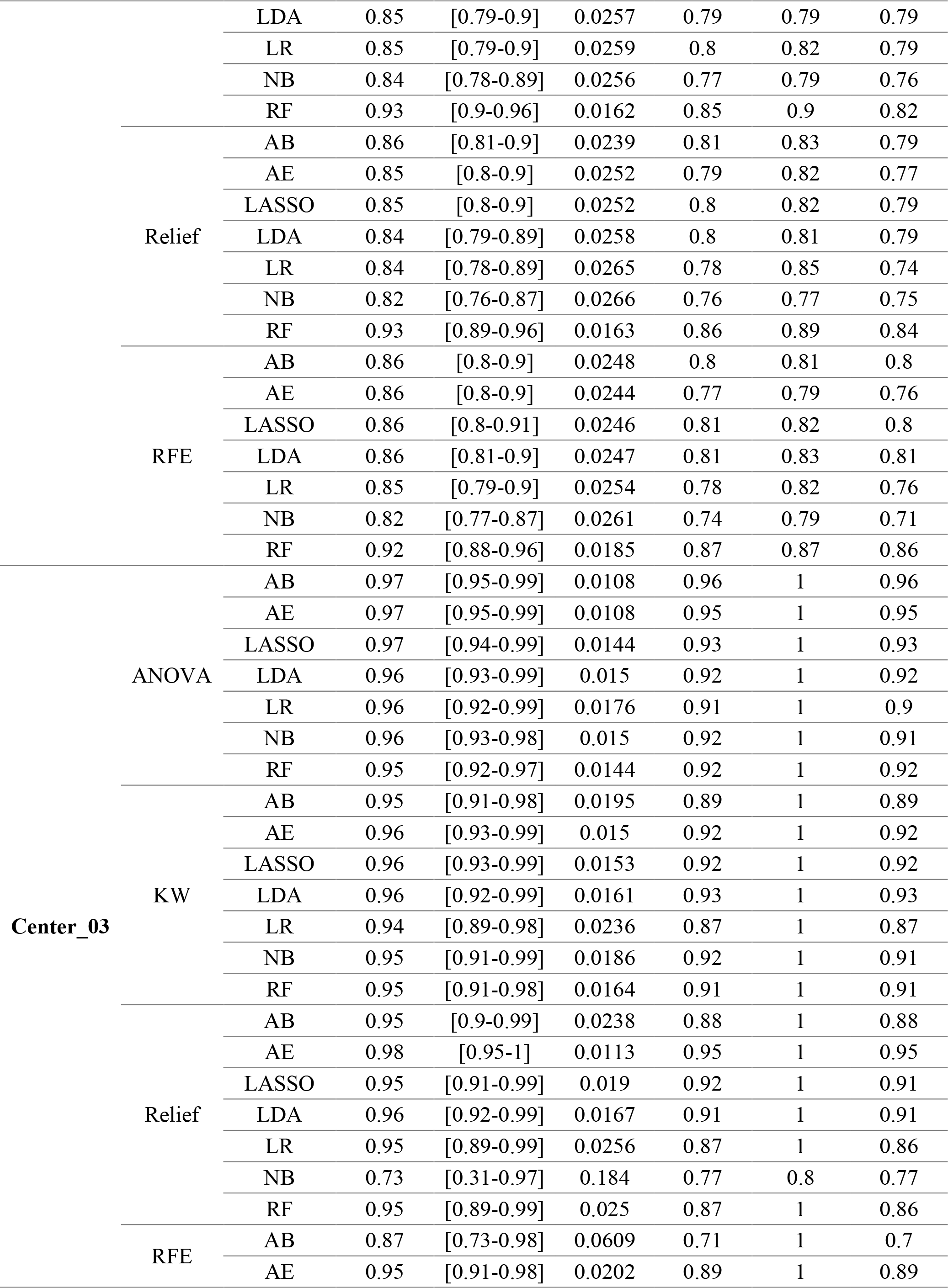
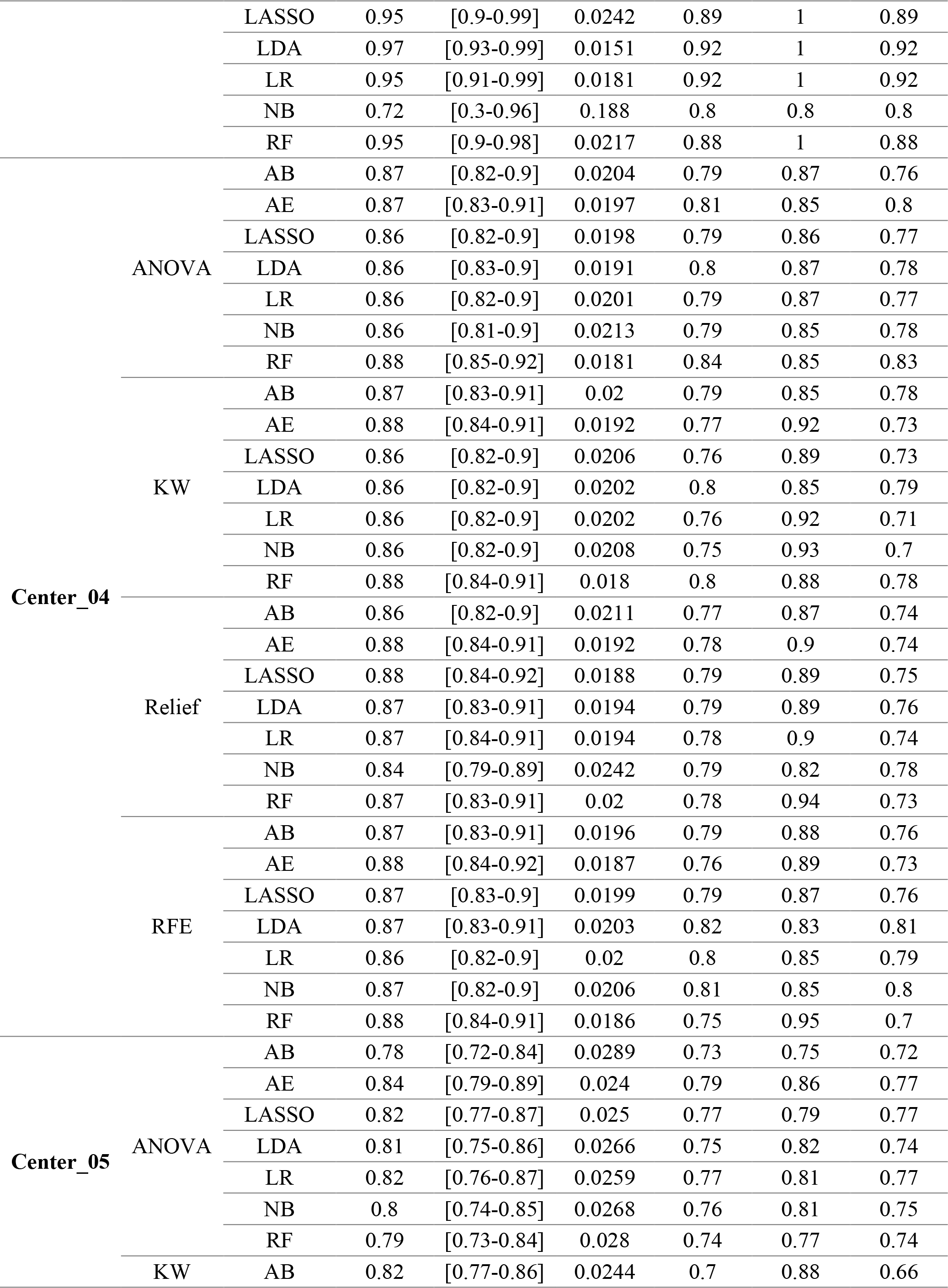
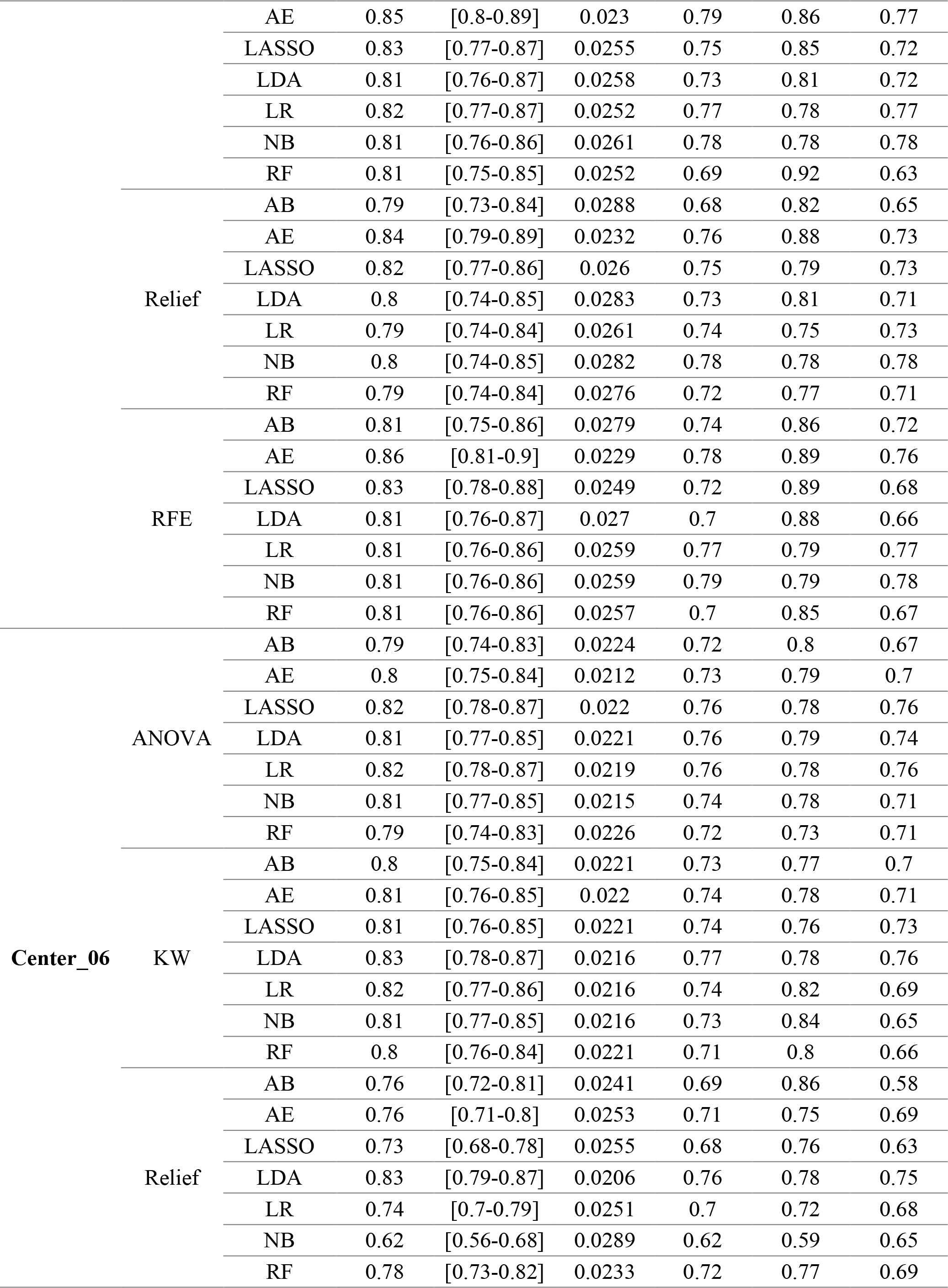
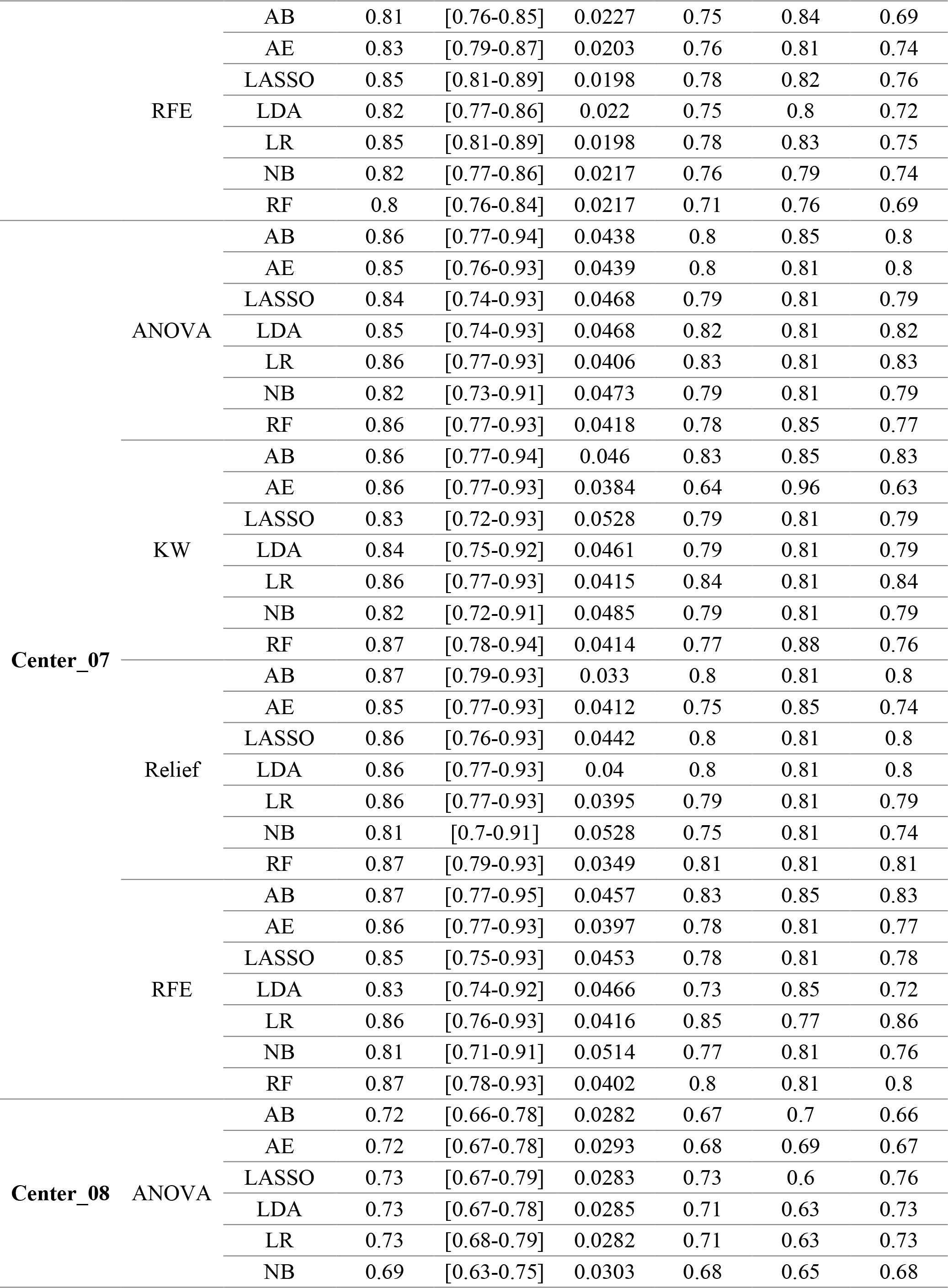
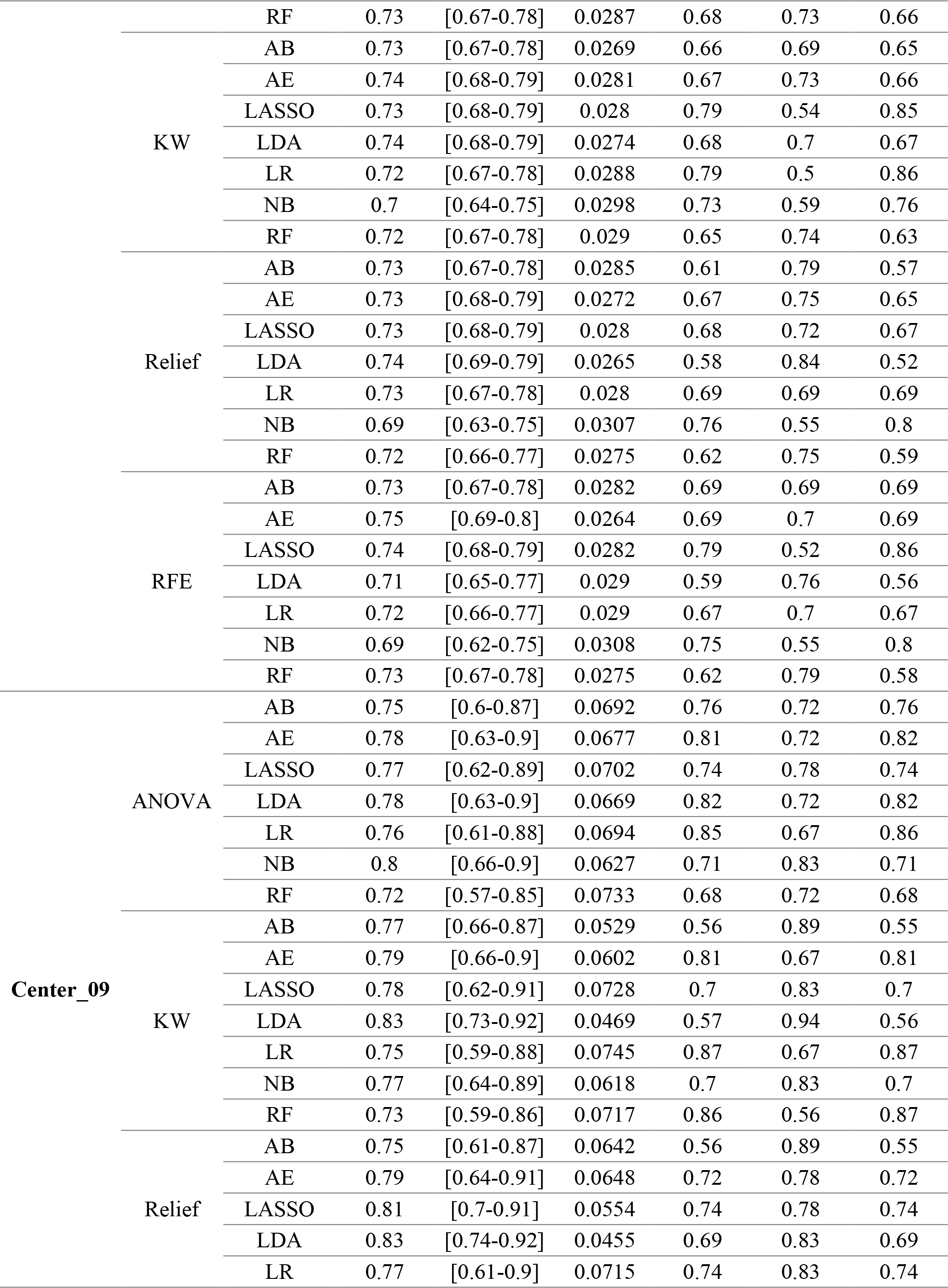
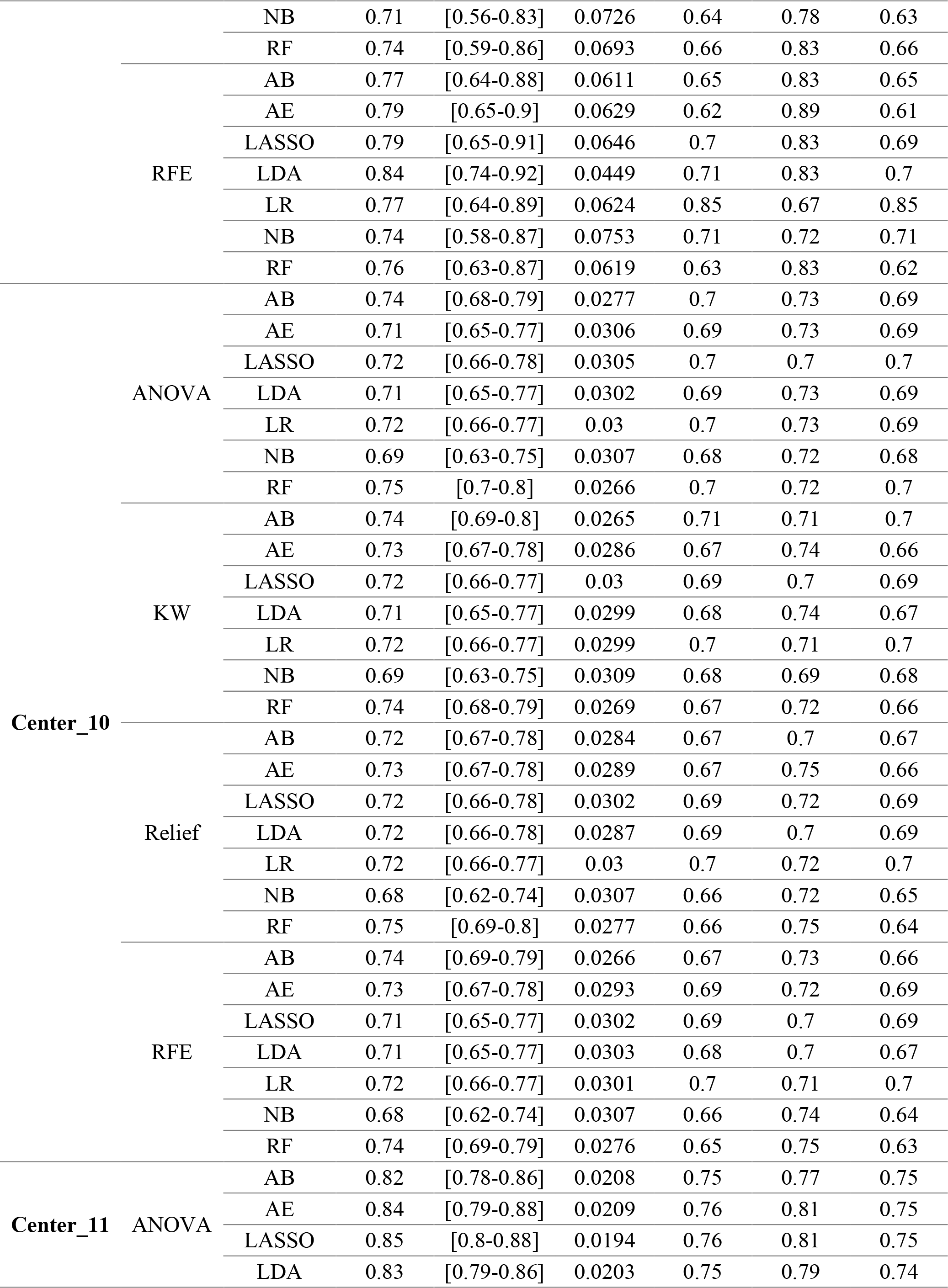
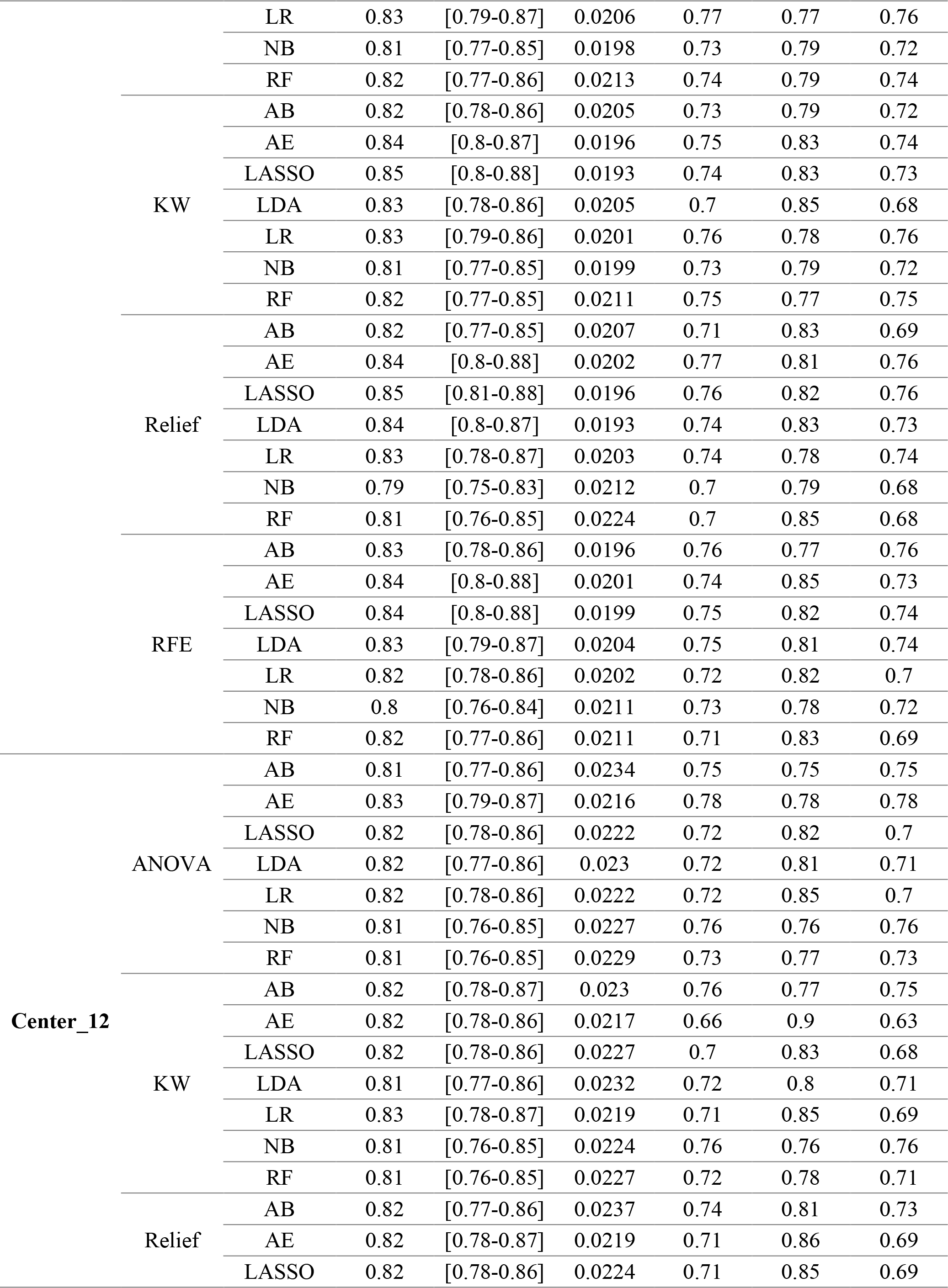
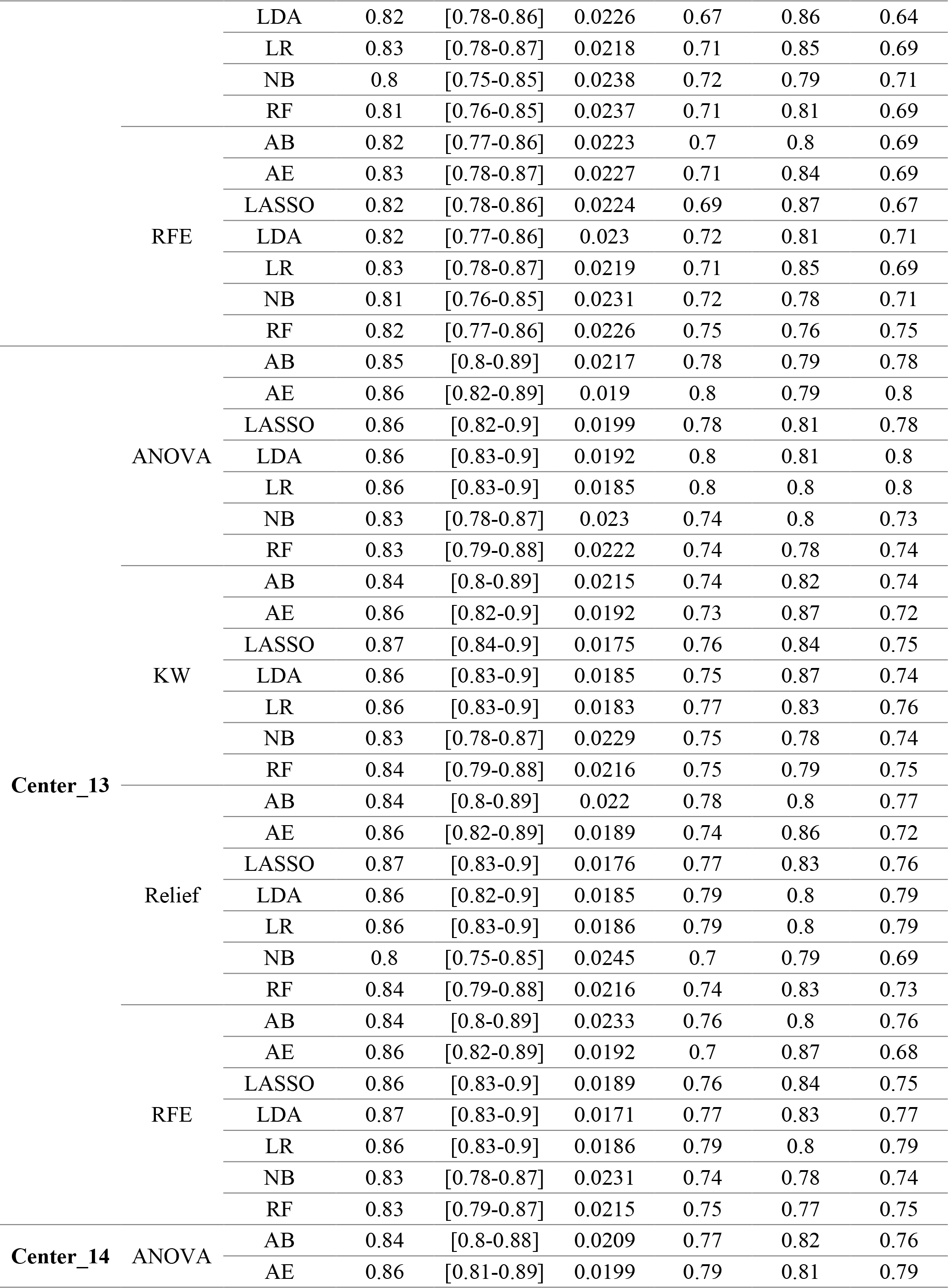
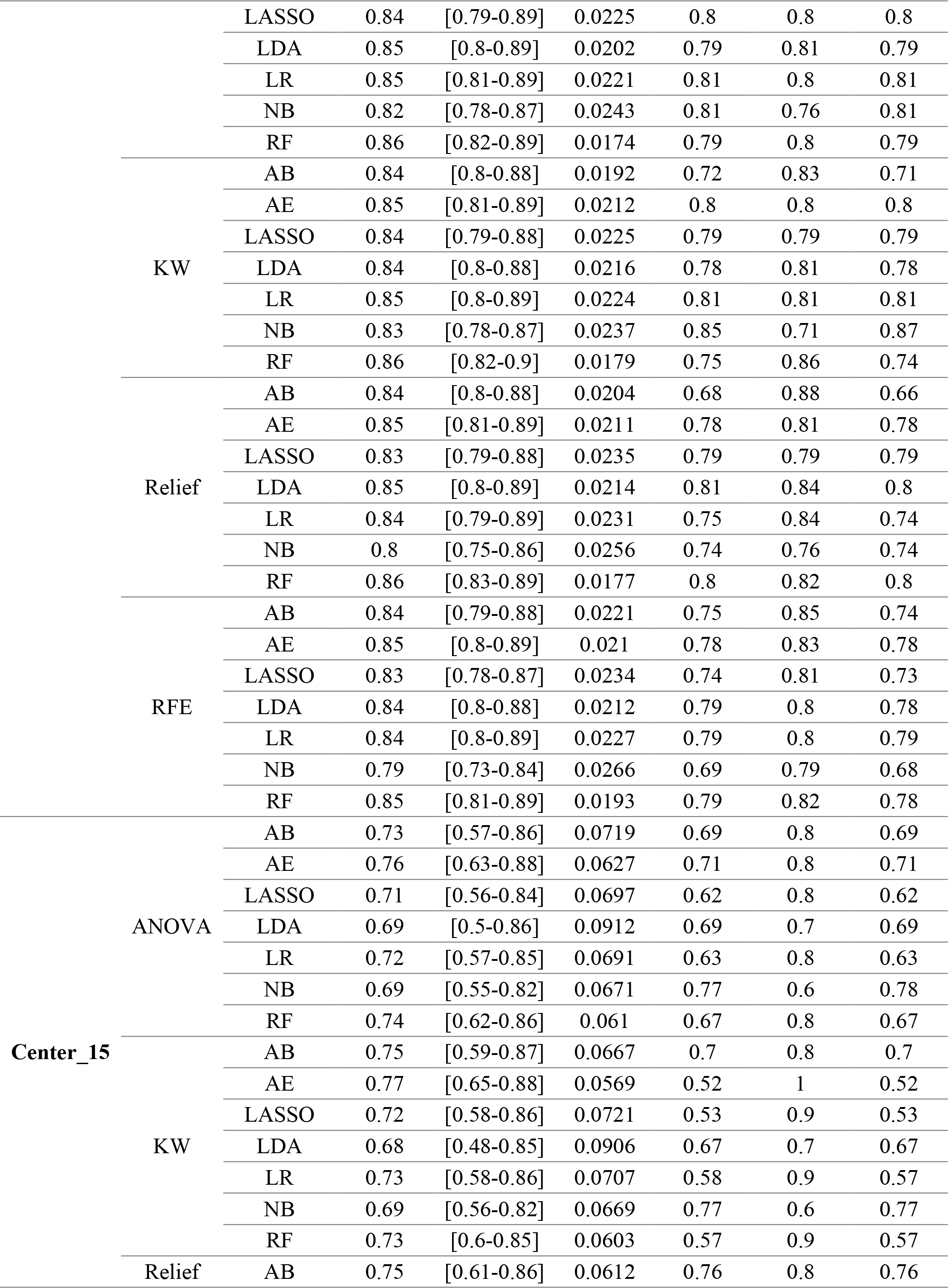
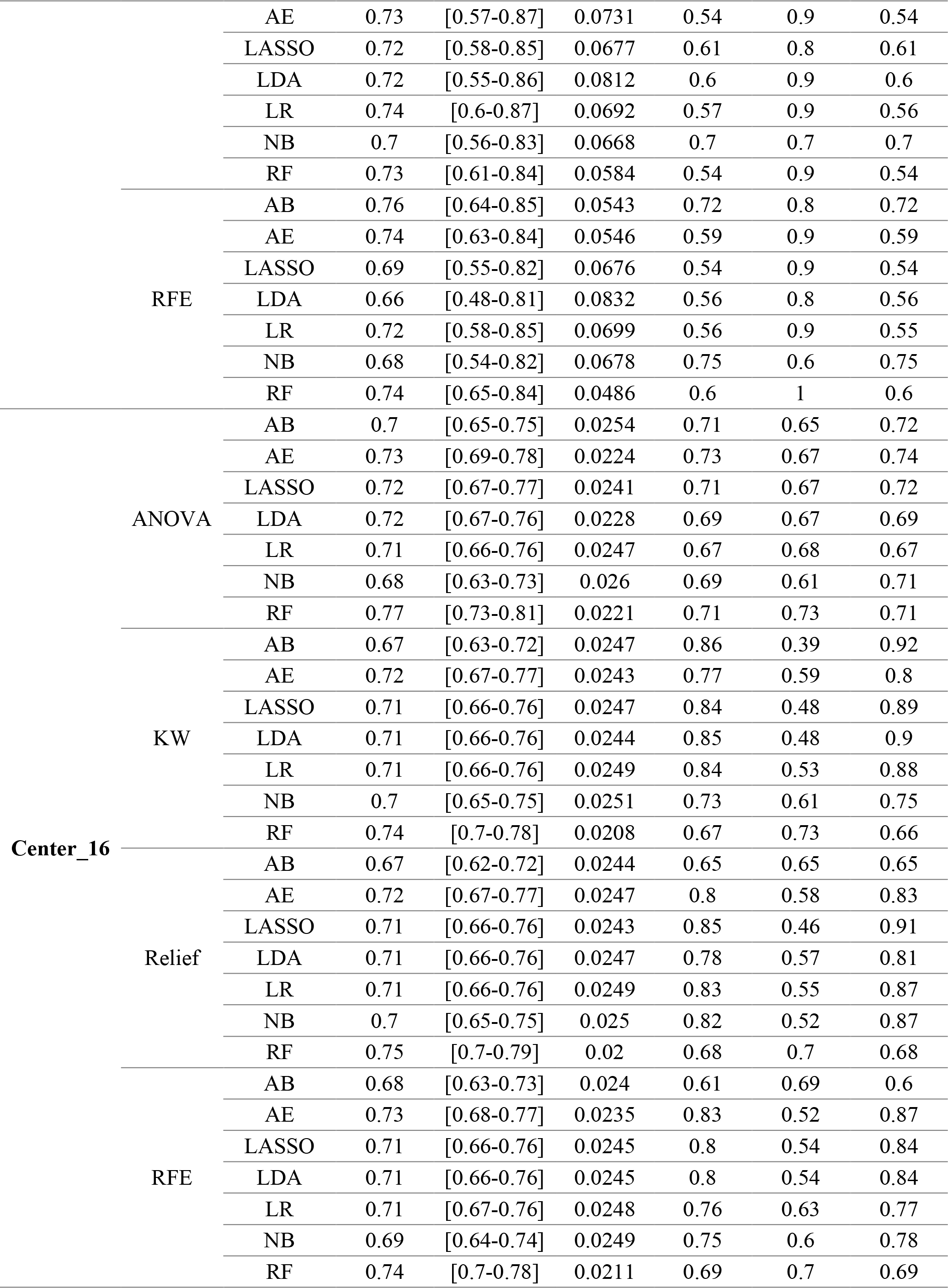
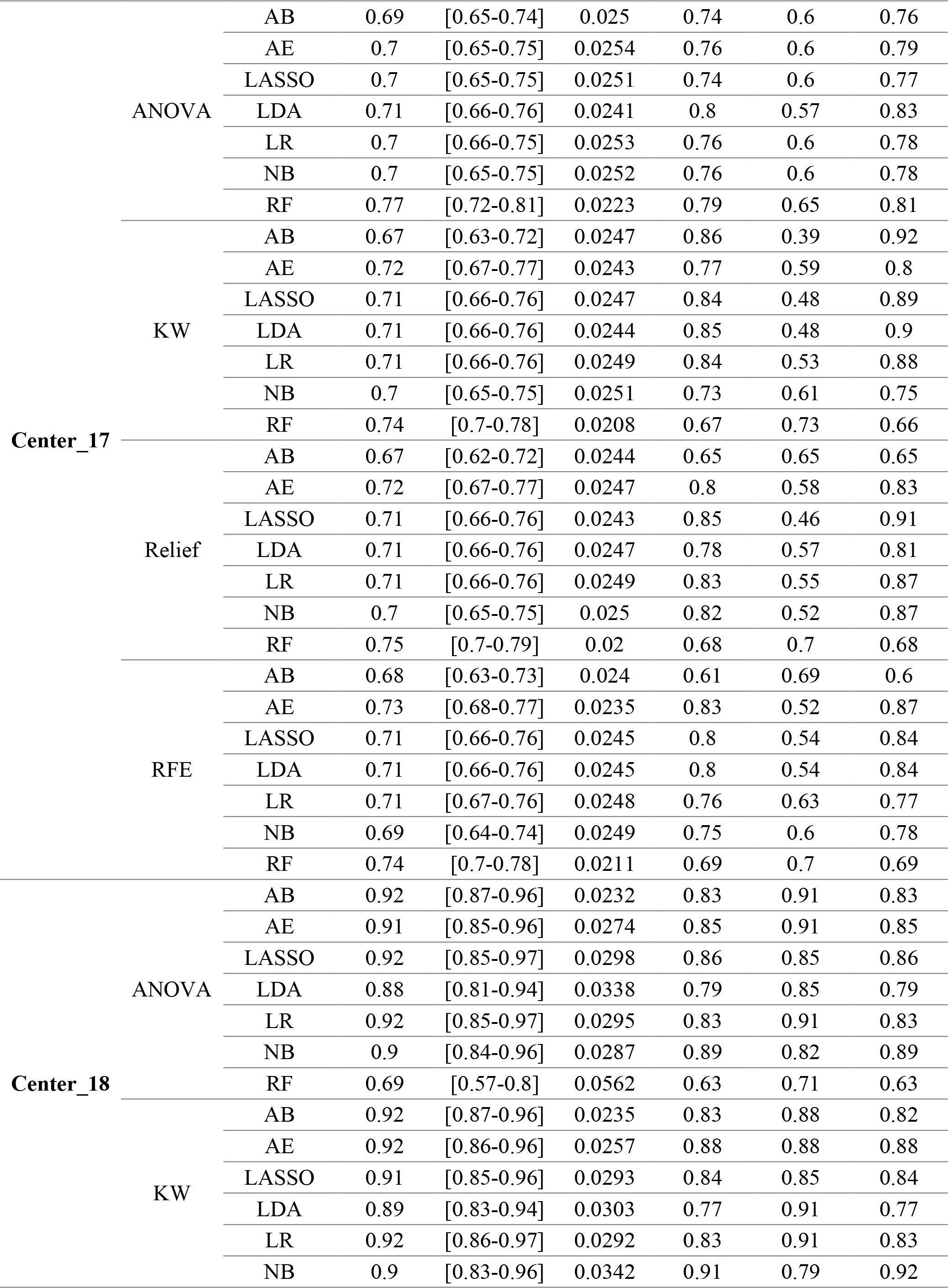
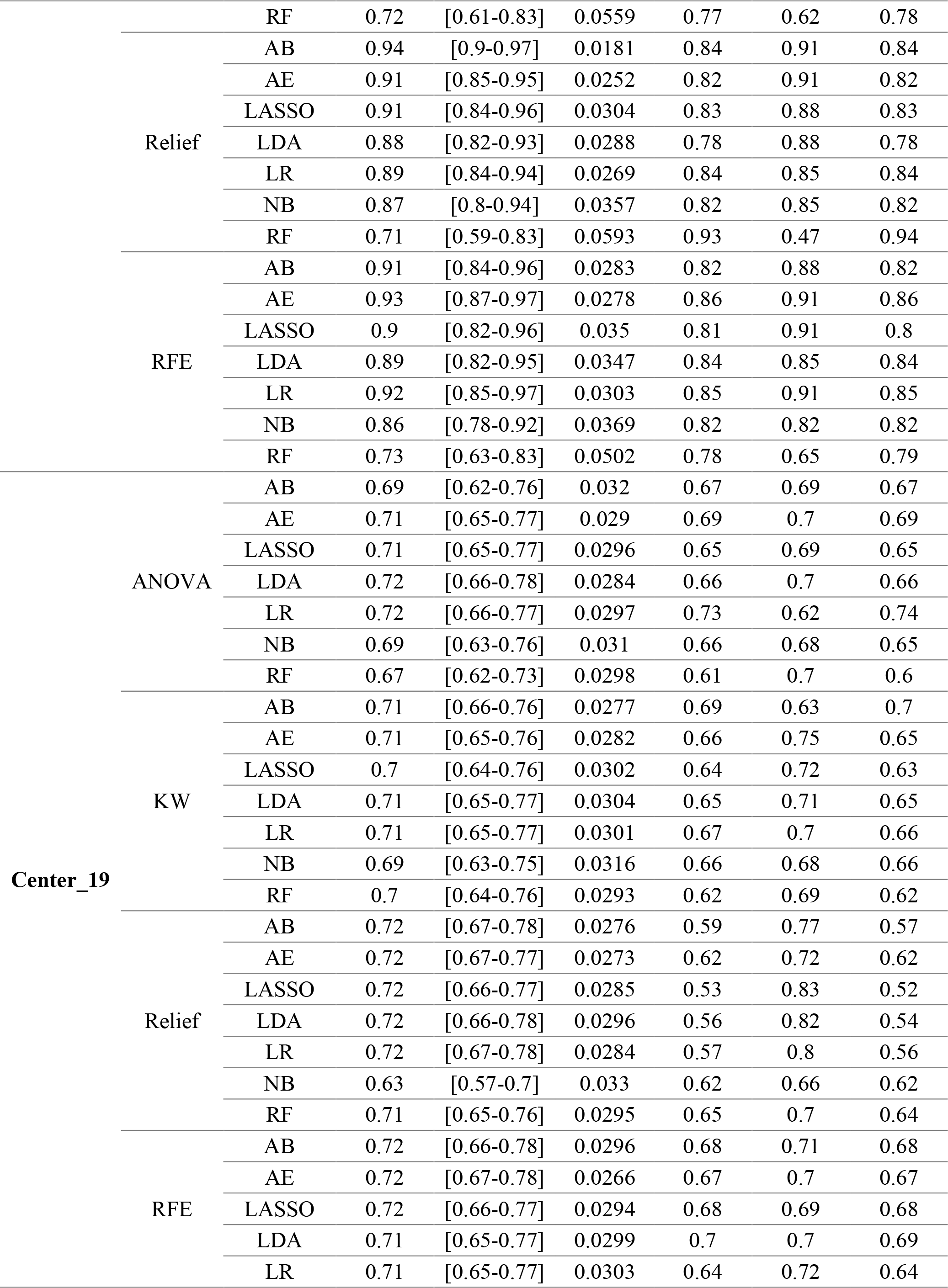
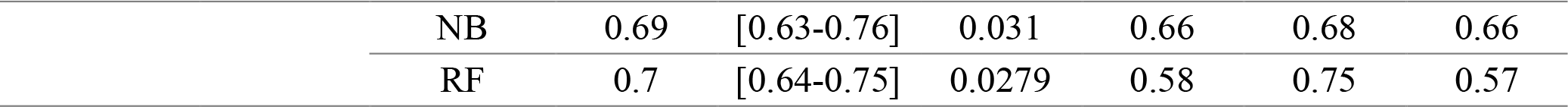
Classification performance indices of different feature selector and classifiers in Strategy 12.

**Supplemental Table 14.**
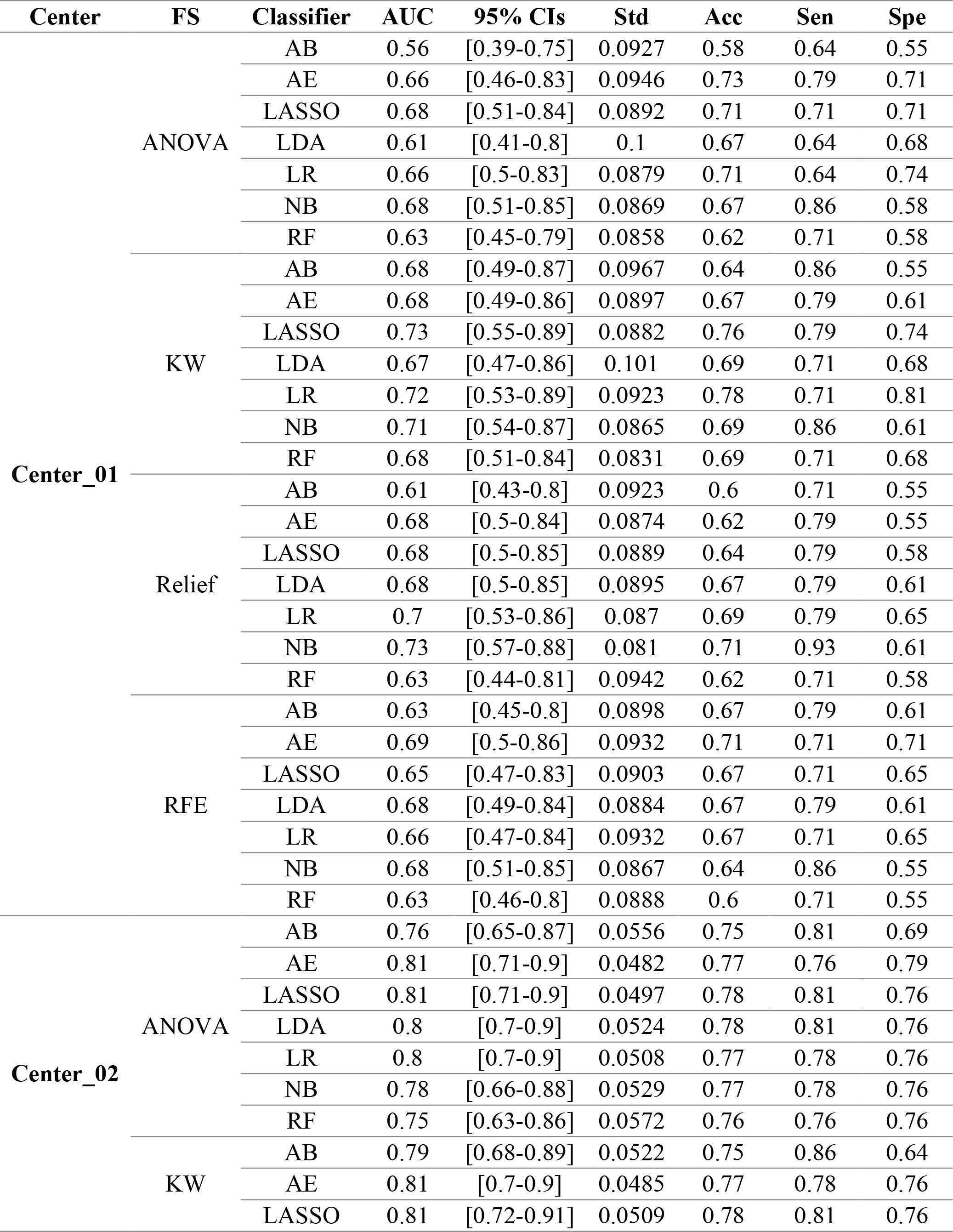
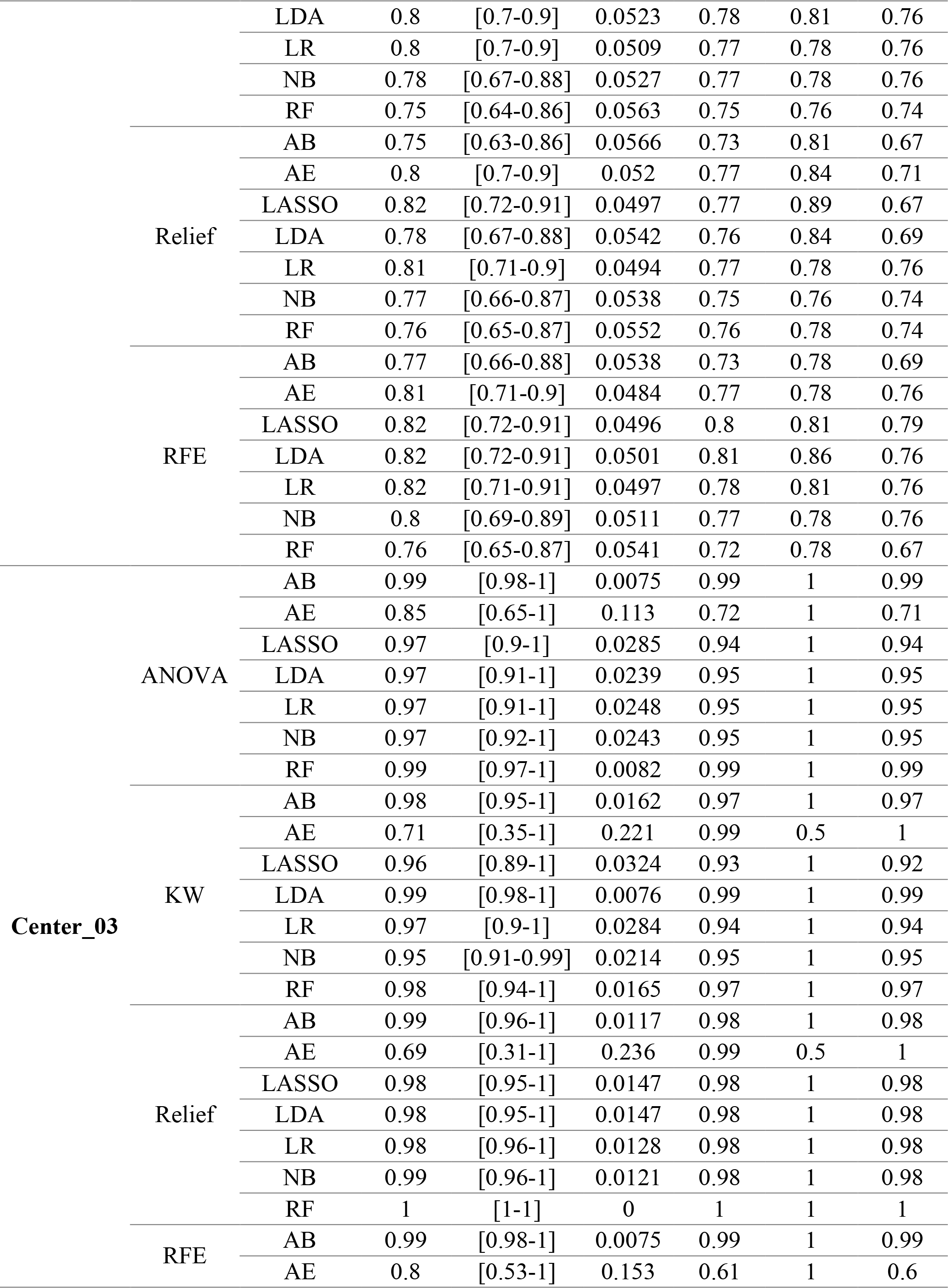
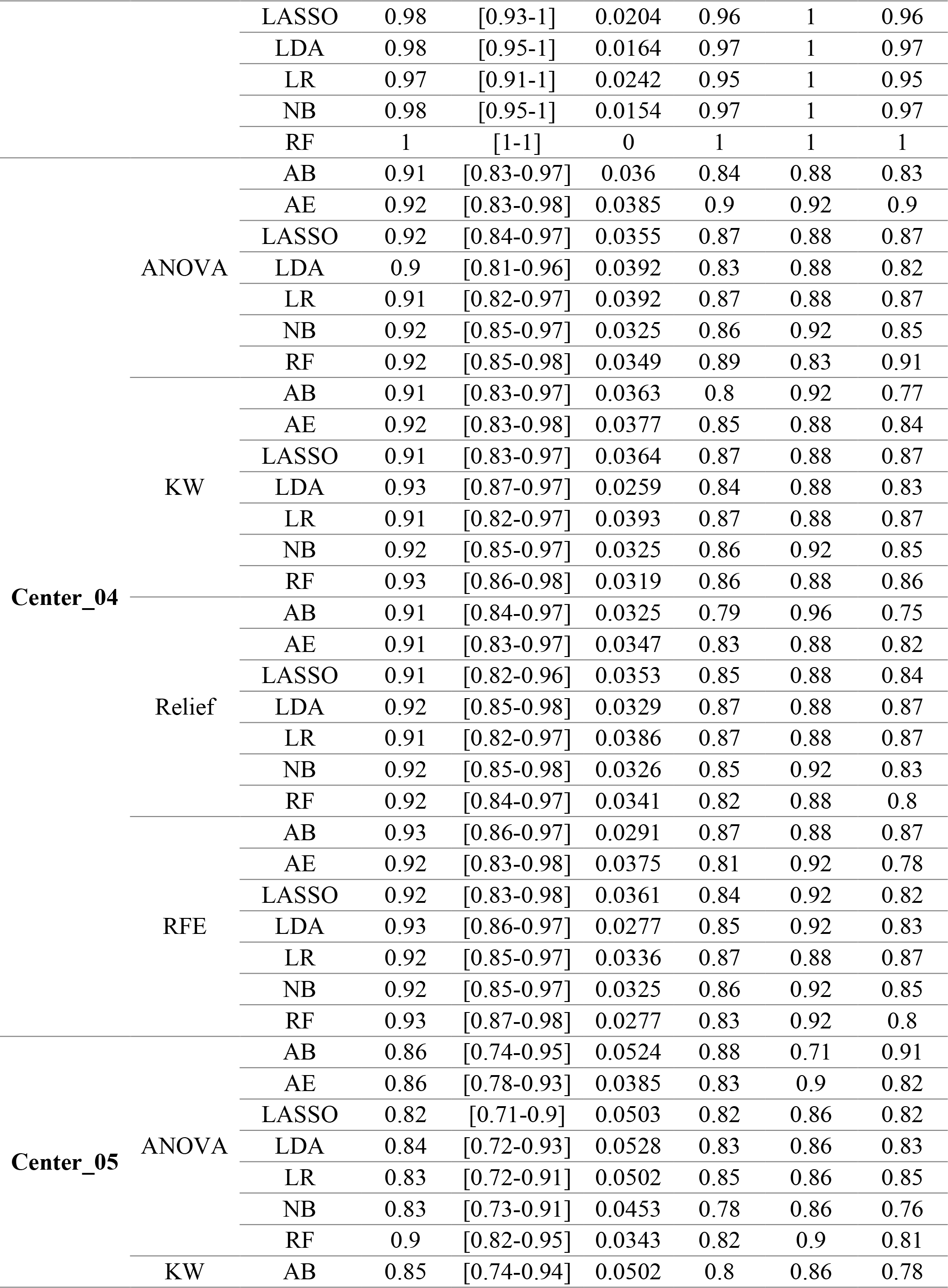
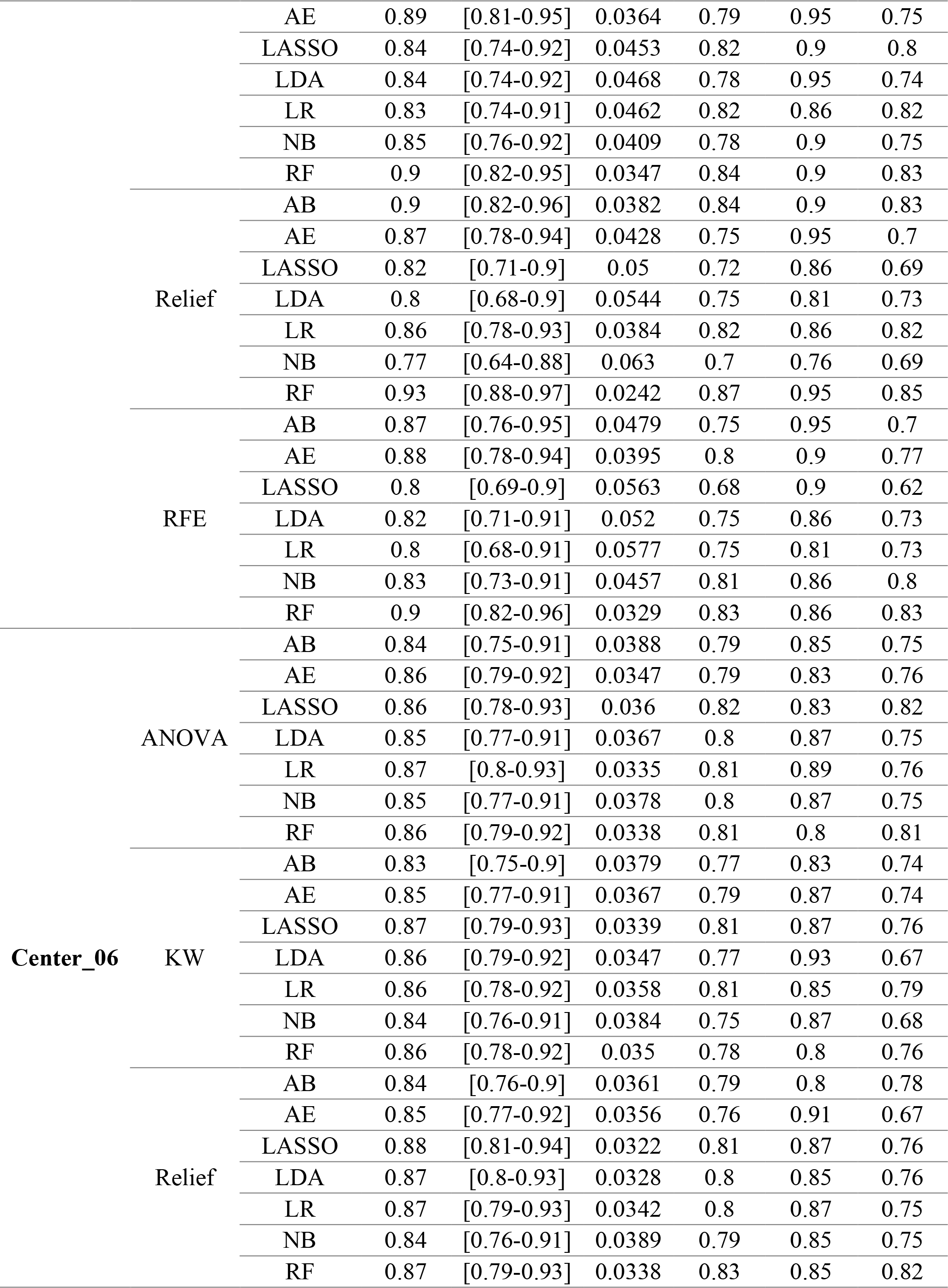
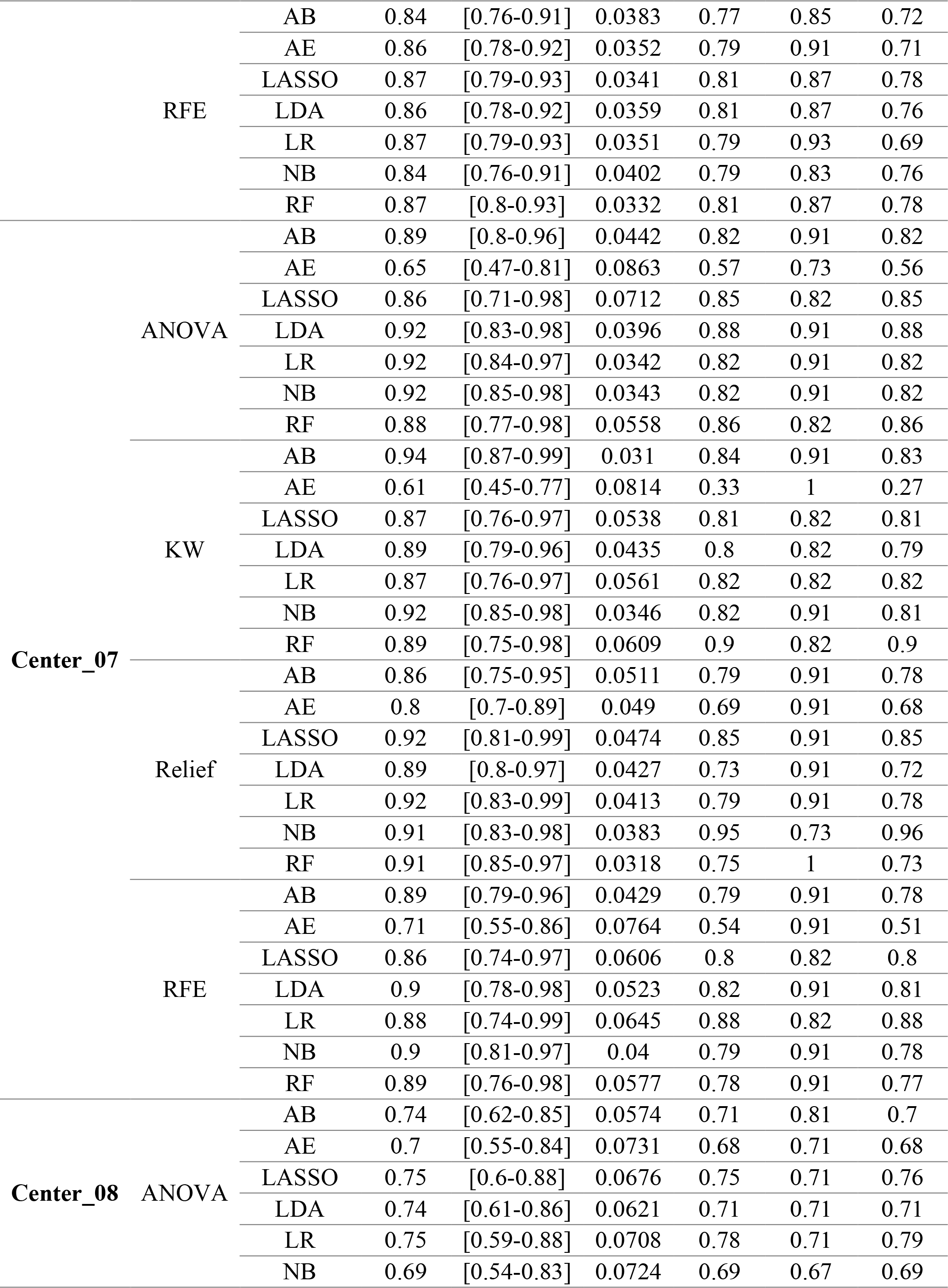
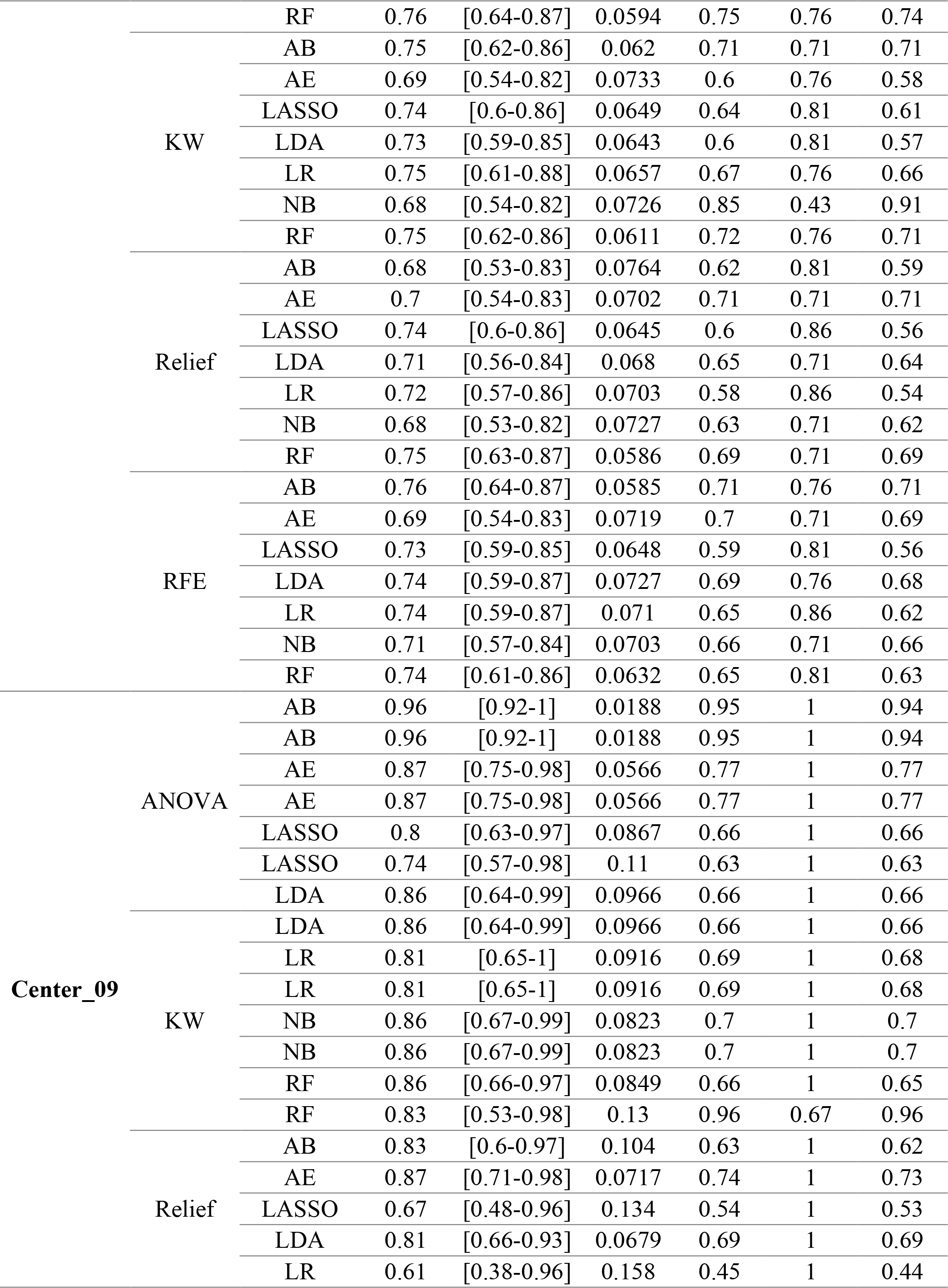
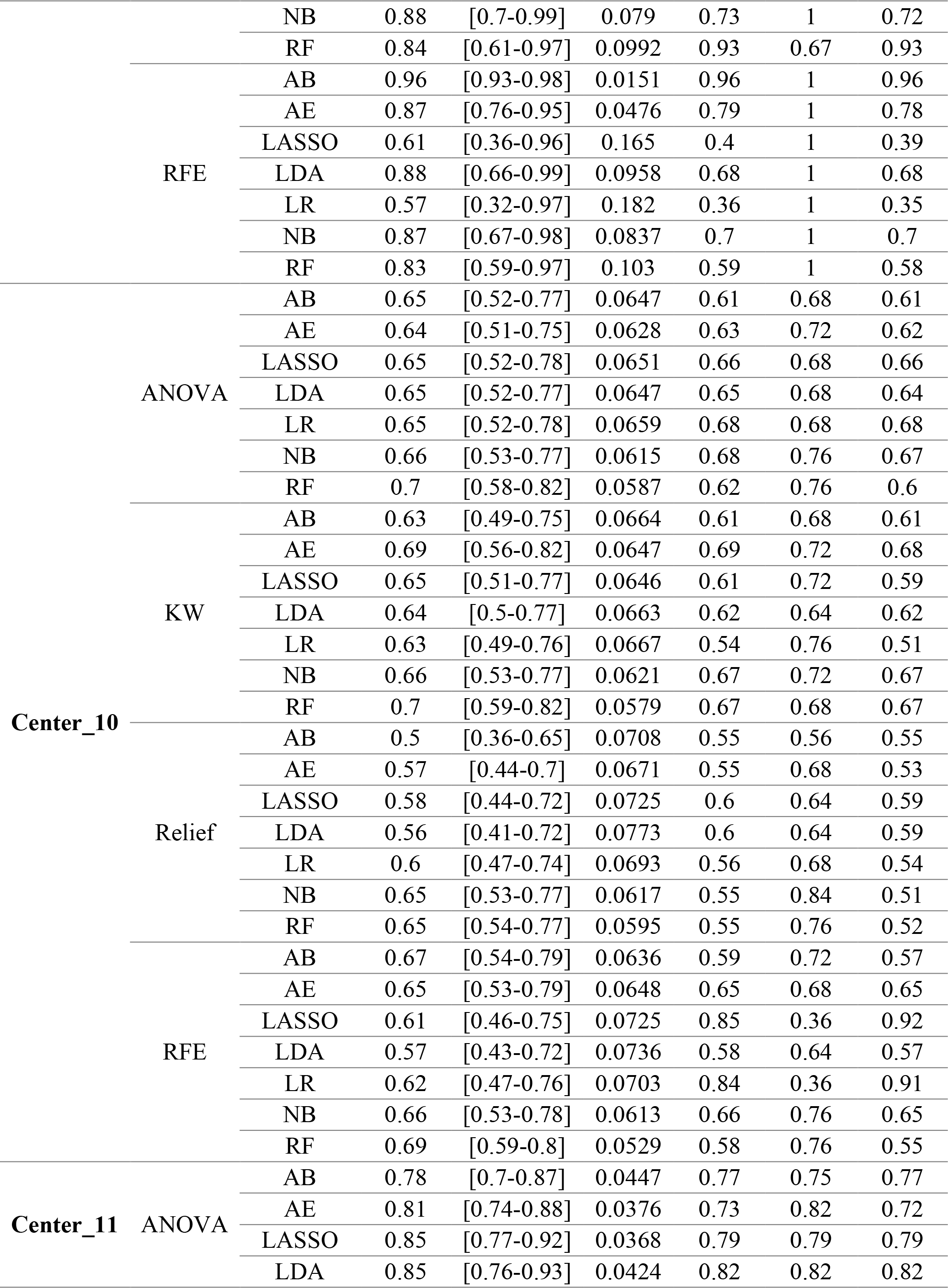
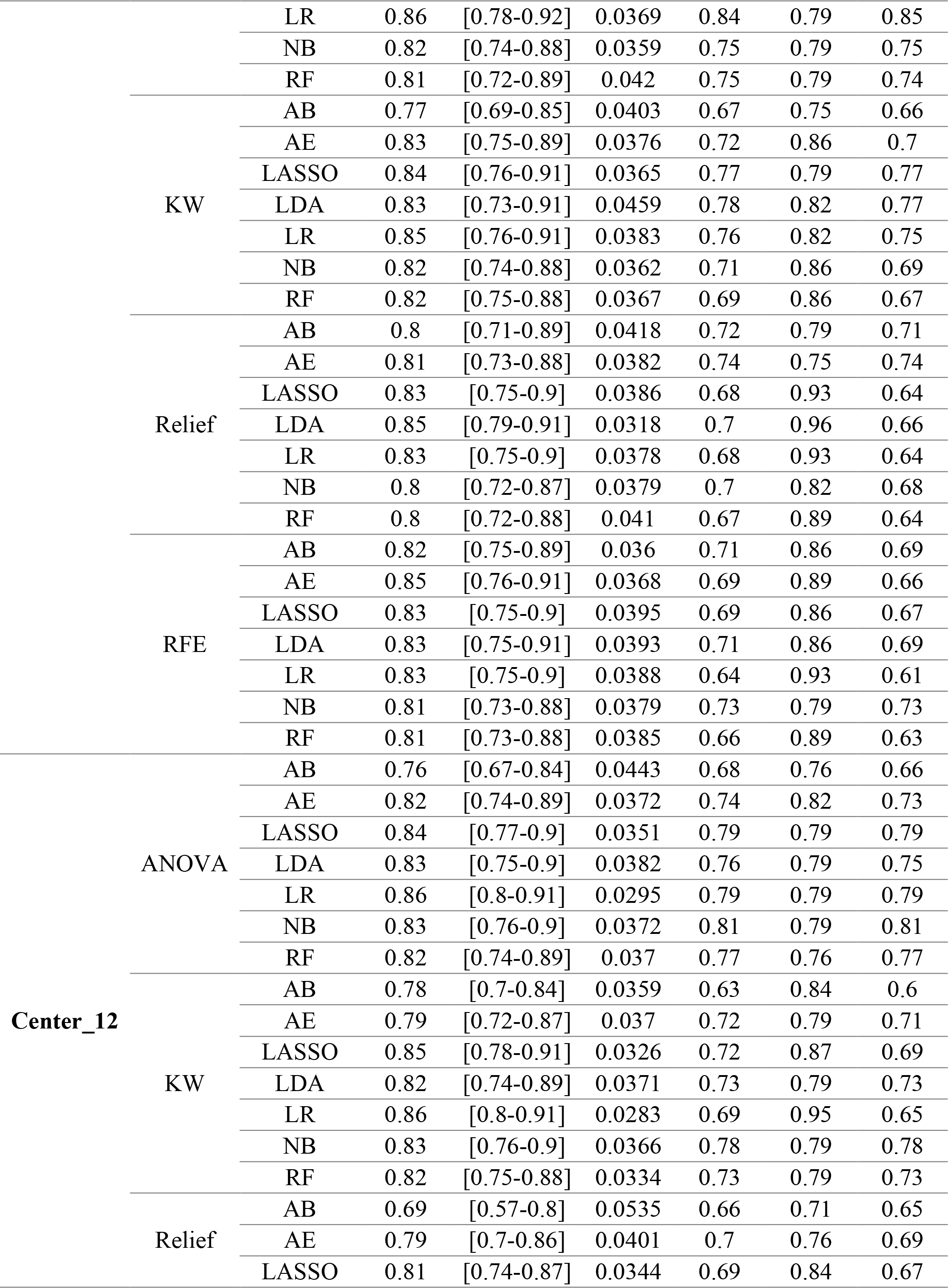
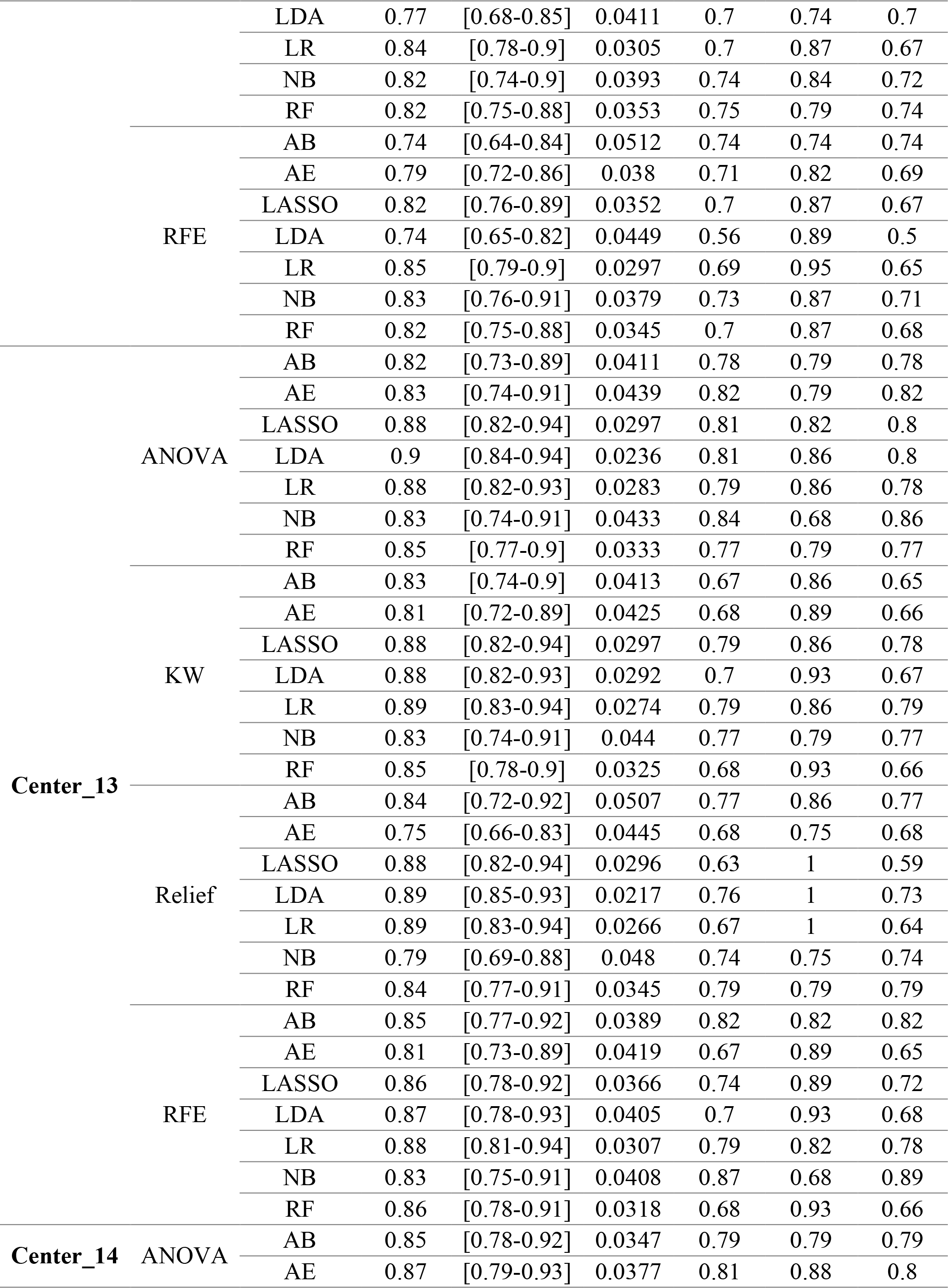
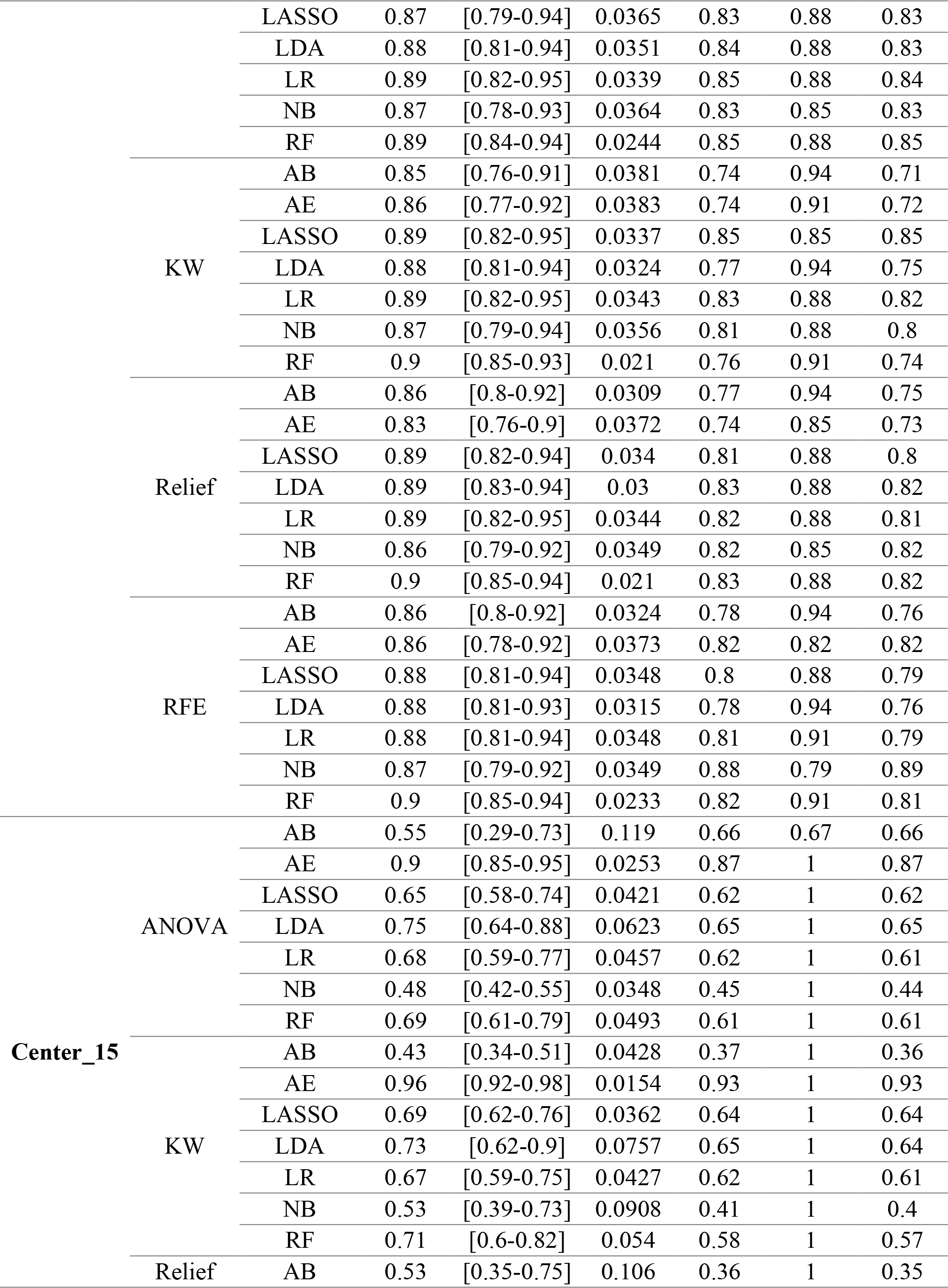
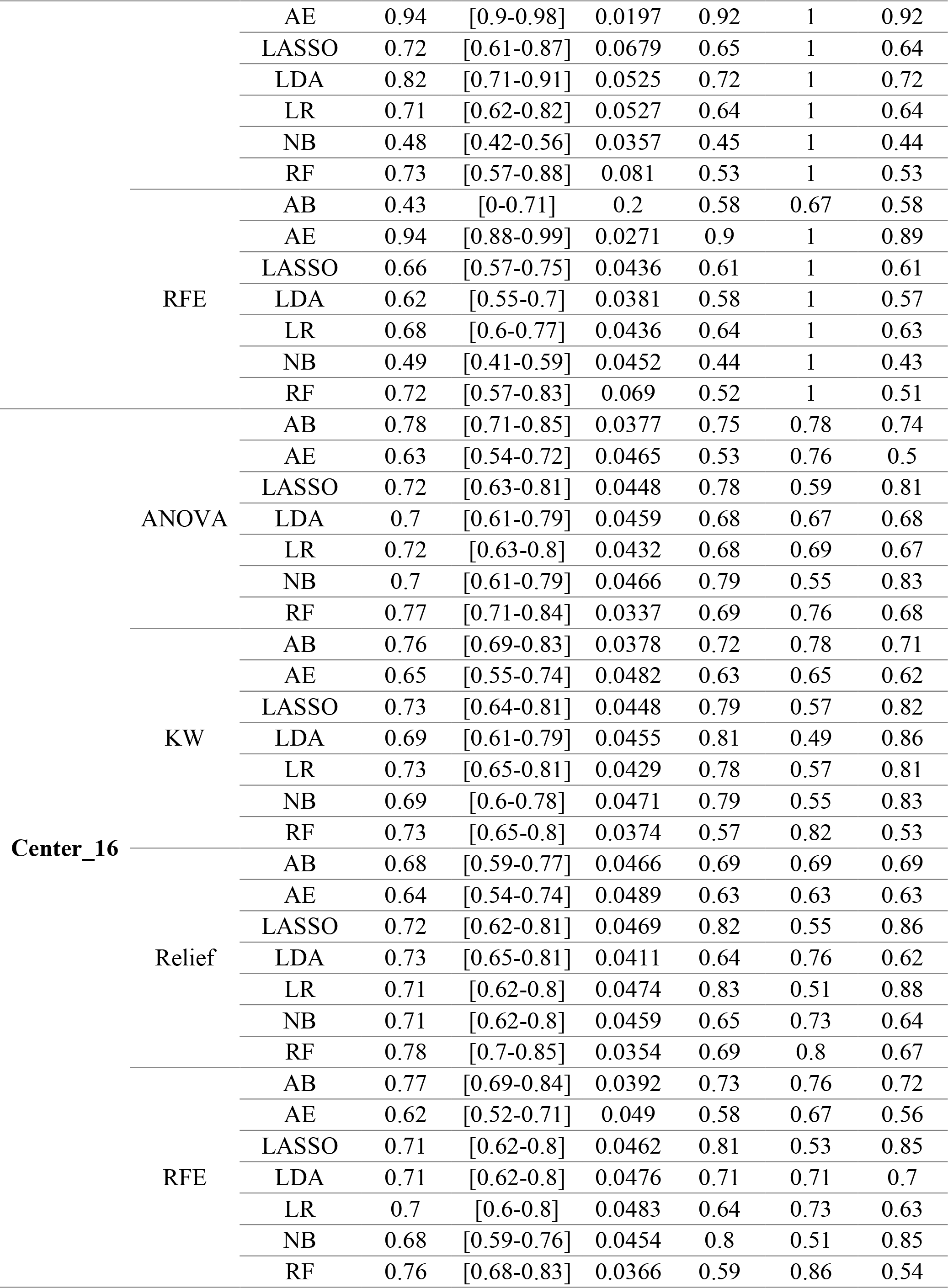
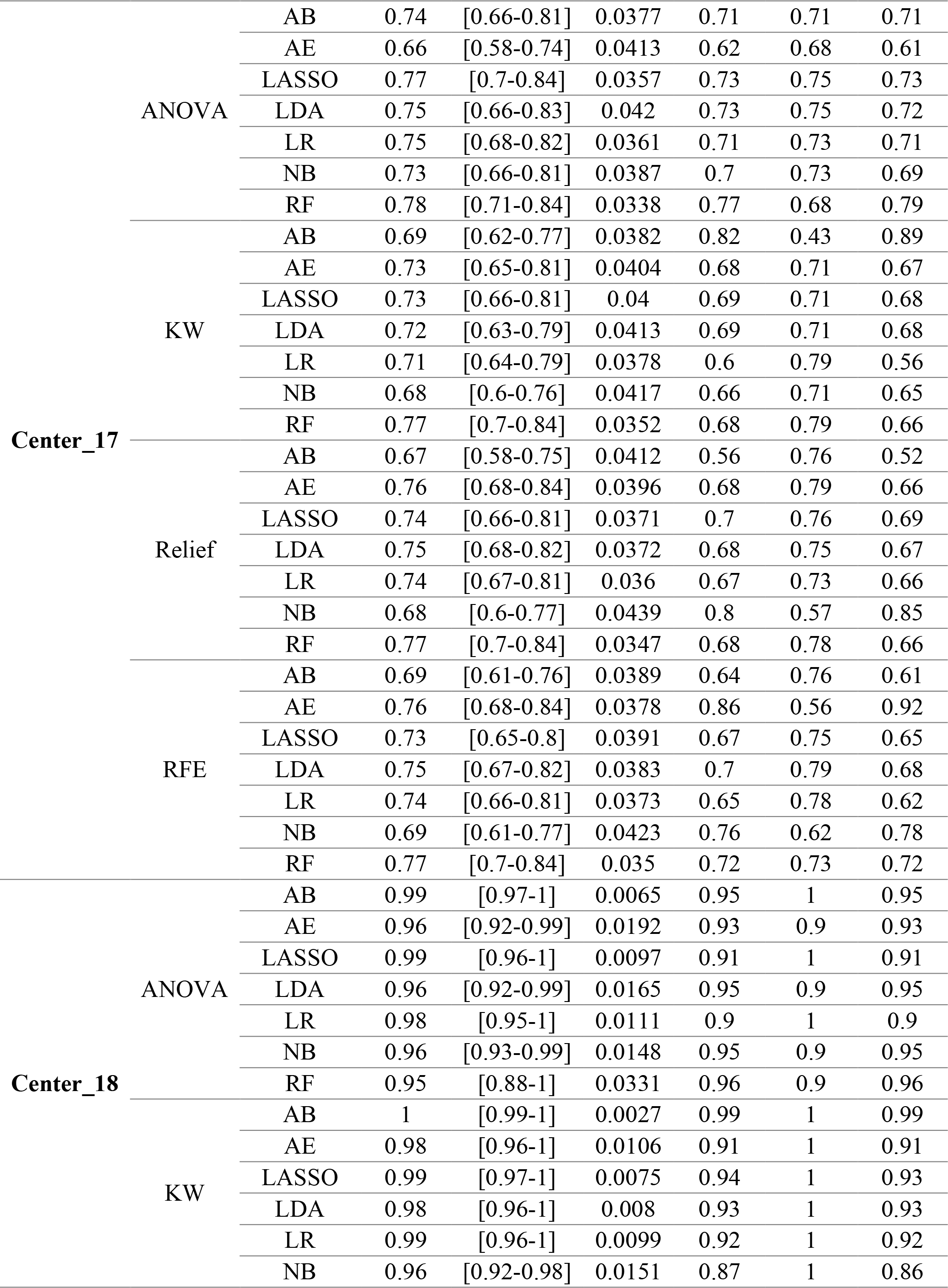
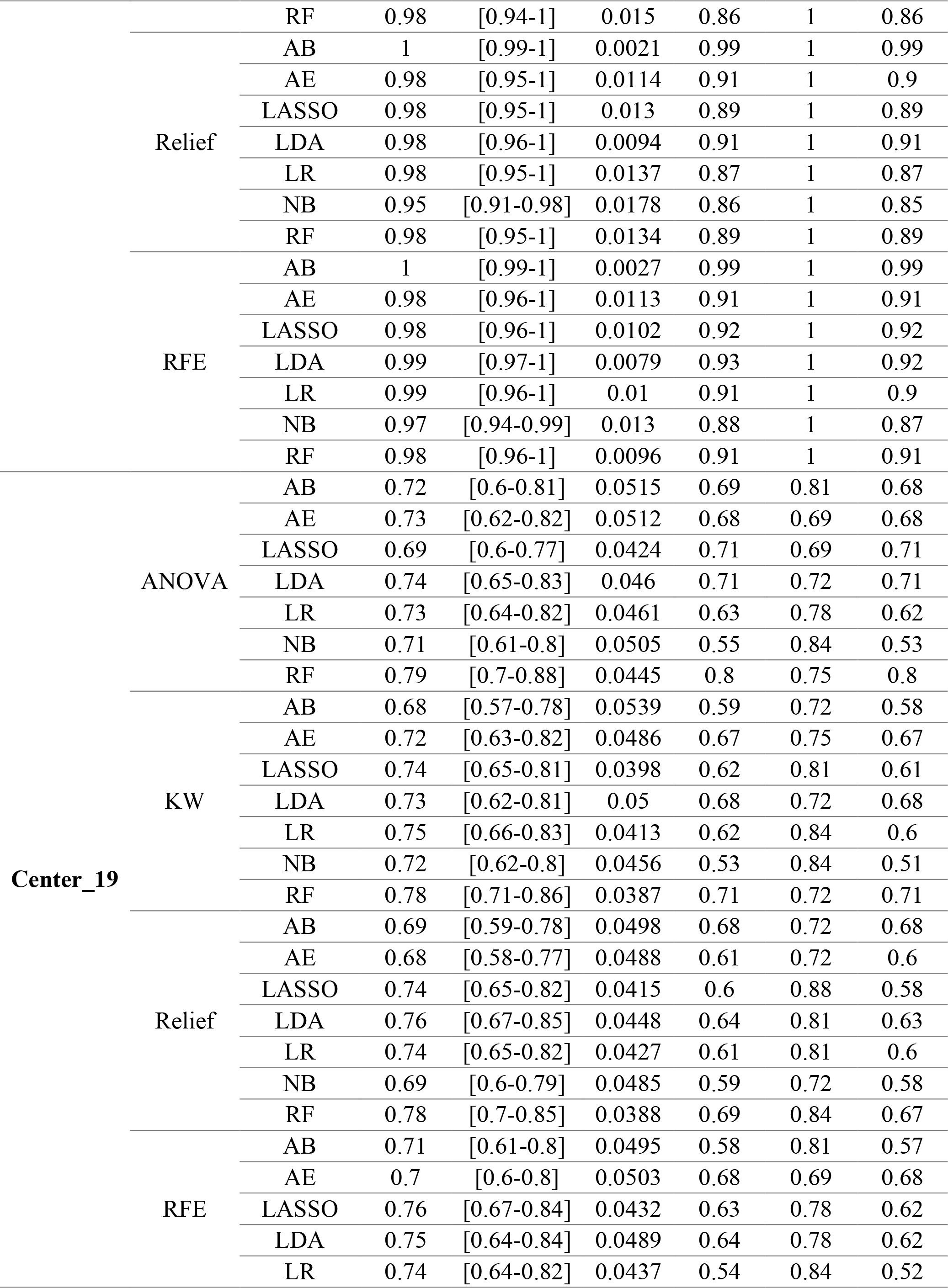
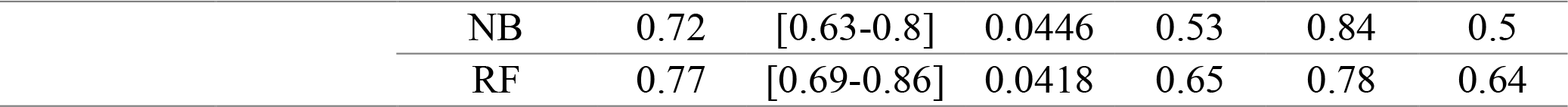
Classification performance indices of different feature selector and classifiers in Strategy 10.

